# Differential Intrahepatic Integrated HBV DNA Patterns Between HBeAg-Positive and HBeAg-Negative Chronic Hepatitis B

**DOI:** 10.1101/2025.02.28.25322668

**Authors:** Daryl T-Y Lau, Elena S Kim, Zhili Wang, Wendy C King, David E Kleiner, Marc G. Ghany, Amanda S Hinerman, Yuanjie Liu, Raymond T. Chung, Richard K Sterling, Gavin Cloherty, Selena Y. Lin, Hsin-Ni Liu, Ying-Hsiu Su, Haitao Guo

## Abstract

**Background:** HBsAg can be derived from intrahepatic cccDNA and integrated HBV DNA (iDNA). We examined the iDNA from liver tissues of 24 HBeAg(+) and 32 HBeAg(−) treatment-naive CHB participants.

**Methods:** Liver tissues were obtained from the North American Hepatitis B Research Network (HBRN). For cccDNA analysis, DNA was heat-denatured and digested by plasmid-safe ATP-dependent DNase to remove rcDNA and iDNA prior to qPCR. For iDNA detection, total DNA was subjected to HBV hybridization-targeted next generation sequencing (HBV-NGS) assay. The HBV-host junction sequences were identified by ChimericSeq. Comparison of HBV cccDNA and iDNA with serum and intrahepatic virological parameters were assessed.

**Results:** Intrahepatic cccDNA, serum HBV DNA, HBV RNA, HBcrAg and qHBsAg were higher among the HBeAg(+) participants. Among the HBeAg(+) samples, 87% had positive intrahepatic HBcAg staining compared to 13% of HBeAg(−) samples (p<0.0001). HBsAg staining, in contrast, was present in over 85% of both HBeAg(+) and (−) livers. 23 (95.8%) HBeAg(+) participants had ≤50% iDNA of total HBV DNA whereas 25 (78.1%) HBeAg(−) participants had >50% iDNA in their livers. The iDNA junction-breakpoint distributions for the HBeAg(+) group were random with 15.9% localized to the DR2-DR1 region. In contrast, 52.4% of the iDNA were clustered at DR2-DR1 region among the HBeAg(−) participants. Microhomology-mediated end joining (MMEJ) patterns of dslDNA HBV integration was more frequent in HBeAg (+) livers.

**Conclusion:** Serum RNA and HBcrAg reflect the intrahepatic cccDNA concentrations. HBeAg(−) CHB participants had high levels of intrahepatic iDNA and HBsAg despite lower cccDNA levels suggesting that iDNA is the primary source of HBsAg in HBeAg(−) CHB.

## INTRODUCTION

Hepatitis B virus (HBV) is a hepatotropic DNA virus that belongs to the *Hepadnaviridae* family.^1^ Despite the availability of an effective preventive vaccine and therapy with nucleos(t)ide analogs (NUCs), chronic hepatitis B (CHB) remains a significant public health issue and is a major cause of end-stage liver diseases including cirrhosis and hepatocellular carcinoma (HCC).^2–4^ Currently, the goal of HBV therapy is to achieve a functional cure that is defined as the suppression of hepatitis B surface antigen (HBsAg) to below the limit of detection at <0.05 IU/ml by the HBsAg assay.^5, 6^ However, only about 10% of the HBeAg(+) patients achieved a functional cure with prolonged NUC therapy over 7 years of continuous use, and functional cure rate is particularly low among patients with HBeAg(−) CHB.^3^

HBV has a partially double-stranded, relaxed circular DNA (rcDNA) genome.^7^ After entering the hepatocyte, the partially double-stranded DNA is completed by host DNA repair machinery in the nucleus to form the covalently closed circular DNA (cccDNA). The cccDNA is a stable, non-integrated DNA that interacts with histones to form a mini-chromosome that serves as the *bona fide* transcriptional template for mRNA production including the pregenomic RNA (pgRNA), which is reverse transcribed into progeny rcDNA.^8, 9^ In about 10% of cases, the reverse transcription produces a double stranded linear DNA (dslDNA) byproduct, which may randomly integrate into the host genome at sites of the host double stranded DNA breaks through non-homologous end joining (NHEJ) or microhomology-mediated end joining (MMEJ) DNA repair mechanism.^10, 11^ The dslDNA is replication defective; the pgRNA coding cassette and the encoded ORFs of HBV PreC/Core and polymerase are disconnected from the enhancer II and core promoter (EnhII/Cp) on the dslDNA form, leading to loss of their expression. In contrast, the regulatory sequence for the S gene remains intact and continues to produce HBsAg. Human and chimpanzee studies have shown that a large portion of HBsAg in HBeAg(−) patients originates from integrated HBV DNA (iDNA) rather than cccDNA.^12–14^ If the majority of HBsAg is derived from iDNA, functional cure may not be achievable. Current serum qualitative/quantitative assays cannot distinguish the sources of HBsAg.

To date, there are limited comprehensive evaluations of the intrahepatic HBV cccDNA and iDNA in CHB patients.^4, 15^ Additionally, serum HBV markers such as HBV RNA and HBcrAg have been shown to serve as surrogate measures of cccDNA and may have potentially important roles in evaluating disease states and in monitoring therapeutic responses.^15–17^ In this study, we systematically evaluated liver biopsies and the corresponding serum/plasma samples from treatment-naive CHB patients to better understand 1) the cccDNA and iDNA profiles of adults with HBeAg (+) and (−) CHB, respectively, and 2) the relationships between liver and peripheral blood biomarkers.

## MATERIALS AND METHODS

### Hepatitis B Research Network (HBRN)

The HBRN is a research network of 28 clinical sites throughout the U.S. and Canada, funded by the National Institutes of Health, initiated to study the natural history of CHB and to conduct clinical trials in both children and adults. The Adult Cohort study (NCT01263587) enrolled HBsAg(+) and (−) participants ≥18 years, between 2012 and 2017, who were not currently on antiviral therapy. Assessment of demographics and clinical features has been previously described.^18^ The HBRN study protocols were approved by the institutional review boards (Research Ethics Board in the case of the Toronto site) of each participating institution and each participant provided written, informed consent.

### Participant Selection

For this analysis, participants of the Adult Cohort study who had not initiated antiviral therapy and had available liver tissues within 24 weeks of their clinical and virological assessment were identified. Among 194 identified participants, 60 participants were initially selected to have representation of HBV DNA levels across the phenotype spectrum.

### Intrahepatic HBV cccDNA quantitation

Approximately 0.5 mm-long segments of 16-gauge needle liver biopsy samples stored in RNA*later* stabilization solution (Invitrogen, #AM7020) were transferred to β-mercaptoethanol-supplemented RLT lysis buffer (Qiagen, #79216) and broken down in PowerBead Tubes, filled with Ceramic 1.5 mm beads (Qiagen #13113-50) using Bead Mill 24 Homogenizer (Fisherbrand™, #15-340-163). Next, total DNA and RNA were isolated using AllPrep® DNA/RNA Micro Kit (Qiagen, #80284), followed by quantitative and qualitative analyses on NanoDrop One (Thermofisher Scientific). For HBV cccDNA analysis, a portion of total DNA prep was heat denatured and digested by plasmid-safe ATP-dependent DNase (PSAD) to remove non-circular DNA species, followed by cccDNA-specific qPCR quantification, as described previously.^19, 20^ Mitochondrial DNA COX3 gene qPCR was used for normalization of cccDNA, and the copy number of the host gene human hemoglobin subunit β (HBB) quantified by qPCR was used to calculate the cccDNA copy number per million cells. The qPCR primers and probes are listed in Supplemental Table 1.

### Intrahepatic HBV iDNA analyses

100 ng of total DNA isolated from each biopsy samples were subjected to previously developed iDNA analyses by using HBV hybridization (Hyb)-targeted NGS (HBV-NGS) assay (JBS Science) as recently described.^21^ The HBV-host junction (HBV-JS) sequences were identified by ChimericSeq^22^ (Supplemental Figure 1). This assay pipeline determines HBV genotype, precore (PC) mutations, basal core promoter (BCP) mutations, % HBV reads of total NGS reads, % HBV-JS of all HBV reads, coverage of HBV reads along the HBV reference sequence, HBV and host breakpoint coordinates. The output files were analyzed for HBV-host homologous 2-10 or 2-25 nt overlaps on HBV-JS (NHEJ vs. MMEJ), and estimation of iDNA fraction in total HBV DNA based on NGS reads and HBV reference sequence (Supplemental Figure 2).

### Liver histology

Liver biopsy slides were stained with hematoxylin and eosin (H&E) and Masson trichrome centrally by University of Pittsburgh Medical Center. The HBRN Pathology Committee centrally scored histological findings blinded to clinical data. Each biopsy was assessed for adequacy by the central pathology committee with a minimum of three portal tracts required to be adequate.^23^ Biopsies were scored for inflammation (histology activity index; HAI) and fibrosis using Ishak scoring system.^24^ HBcAg and HBsAg immunohistology (IH) was performed with the Roche Ventana BenchMark ULTRA System without antigen retrieval. Simultaneous positive and negative control samples were run in parallel. Biopsies were graded according to the percent of immunoreactive hepatocytes for both antigens. Biopsies with both nuclear and cytoplasmic HBcAg staining were grouped by predominant pattern. For HBsAg, IH staining patterns were also assessed: granular cytoplasmic with continuous regions, granular cytoplasmic with scattered hepatocytes, inclusion-like, and membranous staining (i.e., yes/no to each).

### Serum/plasma virological parameters

Serologies were assessed using sera from the same day as the biopsy assessment. For participants missing quantitative HBsAg (which was only measured once every 48 weeks per study protocol) on the day of the biopsy, the closest available HBsAg value within 48 weeks of the biopsy was selected.

Quantitative HBV DNA and qHBsAg were tested centrally by University of Washington. HBV DNA levels were determined using a real-time PCR assay (COBAS Ampliprep/COBAS TaqMan HBV Test, v2.0; Roche Molecular Diagnostics) with a lower limit of detection (LLOD) of 10 IU/mL and lower limit of quantification (LLOQ) of 20 IU/mL. Quantitative HBsAg was tested using the Roche Diagnostics Elecsys platform with LLOD of 0.05 IU/mL.

Quantitative HBV RNA and HBcrAg were tested centrally by Abbott Diagnostics. HBV RNA was isolated from plasma and amplified as previously described,^25^ using the m2000 system and quantified as log_10_ U/ml. Levels below quantification (<1.65 log_10_ U/mL), were randomly imputed using a uniform distribution (0.01-<1.65 log10 U/mL). Non-detected HBV RNA levels were set to 0 log_10_ U/ml. Serum HBcrAg concentrations were measured using a chemiluminescence enzyme immunoassay (Lumipulse G® HBcrAg assay by Fujirebio Europe). The assay has a linear measurement range of 3.0 log_10_ to 6.8 log_10_ U/ml, with 3 log_10_ U/ml being the detection limit. Dilution was not performed for samples with concentration >6.8 log_10_ U/ml.

### Statistical analysis

All biomarkers were converted to the log_10_ scale to accommodate the large range of values. Analyses were performed in the full sample and stratified by HBeAg status. Descriptive statistics were used to report demographic and clinical characteristics. Pearson’s correlation, with Fisher Exact for computing the 95% confidence intervals (95% CI), was used to estimate the strength of associations between 1) HBV cccDNA with HBV RNA and HBcrAg, respectively, 2) HBV DNA and HBV RNA, and 3) serum HBsAg with HBV RNA, HBV DNA and iDNA (HBV-host junctional read), respectively, and test whether associations differed from zero.

## RESULTS

### Study Participants

Of the 60 participants selected for this study, 4 had insufficient intrahepatic HBV DNA; thus, analyses was conducted among 56 participants. The demographic and clinical features of the 24 HBeAg(+) and 32 were HBeAg(−) participants are shown in **Table 1**. There were predominant Asians in both group with similar age. Eleven (45.8%) HBeAg(+) participants had HBV genotype C, and 15 (46.9%) HBeAg(−) participants had genotype B. The mean serum HBV DNA and qHBsAg were significantly higher among the HBeAg (+) participants but the ALT levels were similar. Cirrhosis was present in 1 HBeAg(+) and 3 HBeAg(−) participants. None had liver cancer.

**Table 1.** Demographical and Clinical Features of North American Adults with CHB, overall and by HBeAg status.

|  | **Overall** | **HBeAg(+)** | **HBeAg(-)** |
| --- | --- | --- | --- |
|  | **N=56** | **n=24** | **n=32** |
| Age (years), median (range) | 46 (21-68) | 44 (23-66) | 50 (21-68) |
| Biological sex, n (%) |  |  |  |
| Male | 34 (60.7%) | 12 (50.0%) | 22 (68.8%) |
| Female | 22 (39.3%) | 12 (50.0%) | 10 (31.3%) |
| Race/ethnicity, n (%) |  |  |  |
| White/Non-Hispanic | 7 (12.5%) | 4 (16.7%) | 3 (9.4%) |
| Black/Non-Hispanic | 5 (8.9%) | 2 (8.3%) | 3 (9.4%) |
| Asian/Non-Hispanic | 42 (75.0%) | 17 (70.8%) | 25 (78.1%) |
| Other | 2 (3.6%) | 1 (4.2%) | 1 (3.1%) |
| ALT (ULN), median (IQR) | 2.5 (0.8-21.0) | 3.1 (0.8-11.4) | 1.9 (0.8-21.0) |
| Serum HBV DNA ($\log^{10}$ IU/mL), median (IQR) | 5.8 (2.6-9.2) | 7.9 (3.2-9.2) | 4.5 (2.6-7.7) |
| Serum HBsAg ($\log^{10}$ IU/mL), median (range) | 3.5 (-0.3-5.4) | 4.3 (1.9-5.4) | 3.0 (-0.3-4.3) |
| Serum HBcrAg ($\log^{10}$ U/mL), median (IQR) | 5.3 (0.2-10.3) | 8.2 (5.8-10.3) | 3.8 (0.2-10.3) |
| Plasma HBV RNA ($\log^{10}$ U/mL), median (IQR) | 4.4 (0.1-8.0) | 7.0 (3.1-8.0) | 2.9 (0.1-6.5) |
| HBV Genotype, n (%) |  |  |  |
| A | 10 (18.2%) | 5 (21.7%) | 5 (15.6%) |
| B | 19 (34.5%) | 4 (17.4%) | 15 (46.9%) |
| C | 18 (32.7%) | 11 (47.8%) | 7 (21.9%) |
| D | 8 (14.5%) | 3 (13.0%) | 5 (15.6%) |

The majority (87%) of HBeAg(+) biopsies stained positive for HBcAg compared to 13% of HBeAg(−) samples (p<0.0001). In contrast, 96% of HBeAg(+) and 88% of HBeAg(−) livers stained positive for HBsAg (p=0.28). Granular cytoplasmic HBsAg staining was noted in 57% and 64% of HBeAg(+) and (−) livers respectively (p=0.75). in contrast, membranous HBsAg stain pattern was noted in only 14% of HBeAg(−) compared to 78% of HBeAg(+) samples (p<0.0001). The detailed clinical characteristics of each of the enrolled 56 patients are listed in Supplemental Table 2.

### Correlations between intrahepatic and serum/plasma virological markers

The mean HBV cccDNA level for HBeAg(+) was significantly higher compared to that of the HBeAg(−) participants [4.97 vs. 3.86 log_10_ copies/1 million cells, p<0.0001]. For the entire study cohort, there were positive correlations between HBV cccDNA and plasma HBV RNA [r=0.76, p<0.001] and serum HBcrAg [r=0.68, p<0.001] (**Figure 1**). With subgroup analysis, the correlations between HBV cccDNA and plasma HBV RNA were lower, but remain significant for HBeAg(+) [r=0.47, p<0.02] and HBeAg(−) [r=0.49, p<0.004] participants. On the contary, the correlations between cccDNA and serum HBcrAg became insignificant in the separate HBeAg(−) group [r=0.33, p=0.06]. Correlatin was not reported in the separate HBeAg(+) group for HBcrAg concentration was above the limit of quantification in 92% of the participants and dilution was not performed in those samples.^26, 27^ For both HBeAg(+) and (−) groups, the serum HBV DNA and plasma HBV RNA had strong correlations [HBeAg(+) r=0.93, p<0.001; HBeAg(−) r=0.87,p<0.001]. There were significant correlations between serum qHBsAg levels and plasma HBV RNA [r=0.44, p=0.03] and serum HBV DNA [r=0.54, p=0.004] for HBeAg(+) but not (−) participants. The correlation between serum qHBsAg and HBV-host junctional read (or iDNA) was only significant among the HBeAg (−) participants [r=0.40, p=0.022].

**Figure 1.**
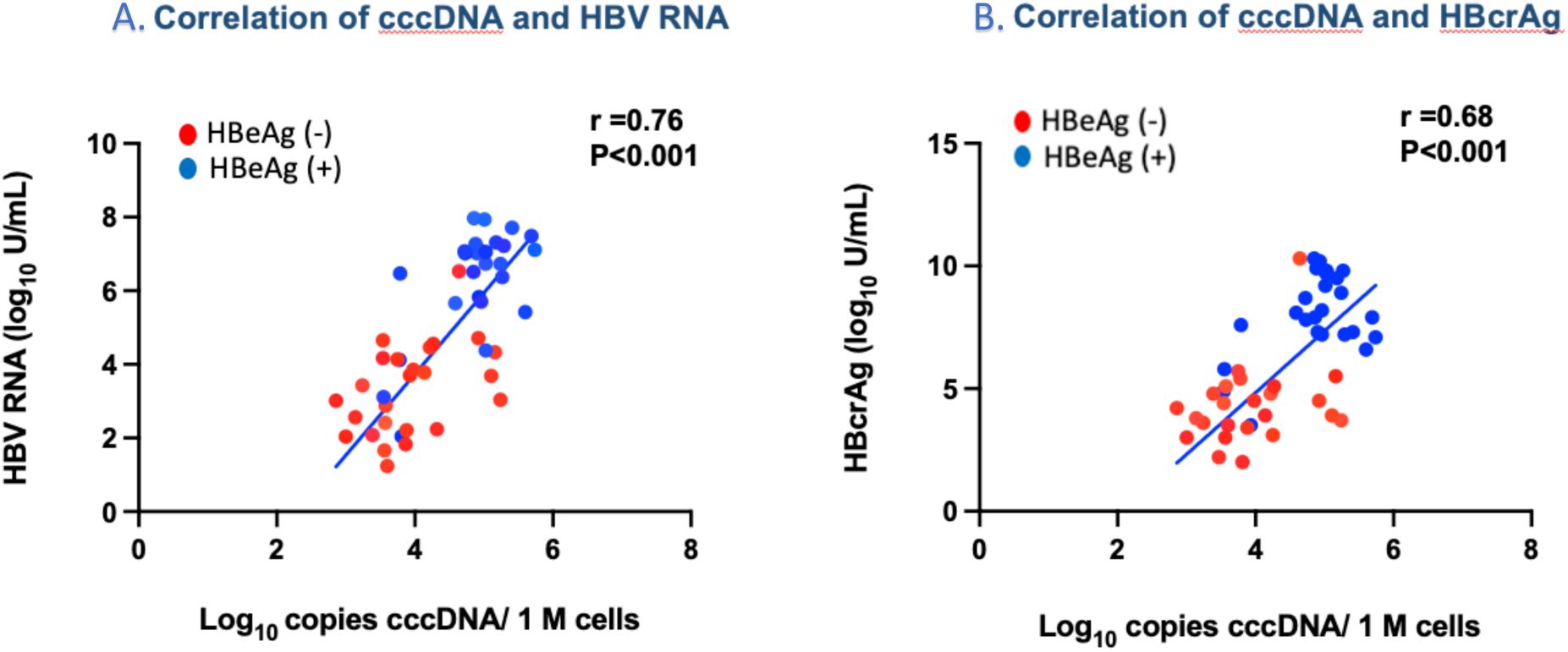
Correlations between cccDNA with (A) plasma HBV RNA, [r=0.76 (95%CI: 0.63, 0.85); p<.001and (B) HBcrAg [r=0.68 (95%CI: 0.51, 0.80); p<.001] in the entire Cohort.

### Comparison of iDNA between HBeAg(+) and (−) CHB

Based on the ChimericSeq results, the distributions and coverage depths of the HBV-host junction sequences (HBV-JS) breakpoints were mapped systematically across the HBV genome for the entire groups of the HBeAg(+) and (−) participants (**Figure 2A-B**). We specifically focused on the read distribution at the Direct Repeat (DR) 2 and DR1 loci (nucleotide (nt) 1600-1900 of the HBV genome) as this region is known to be the “hot spot” for HBV integration as previously characterized in HCC.^10, 28–30^ The junctional sequences for the HBeAg(+) group were distributed randomly across the HBV genome with only 15.9% concentrated in the corresponding DR2-DR1 region (**Figure 2A**). For the HBeAg(−) group, in contrast, 52.4% of the junctional sequences were clustered at the DR2-DR1 region (**Figure 2B**).

**Figure 2.**
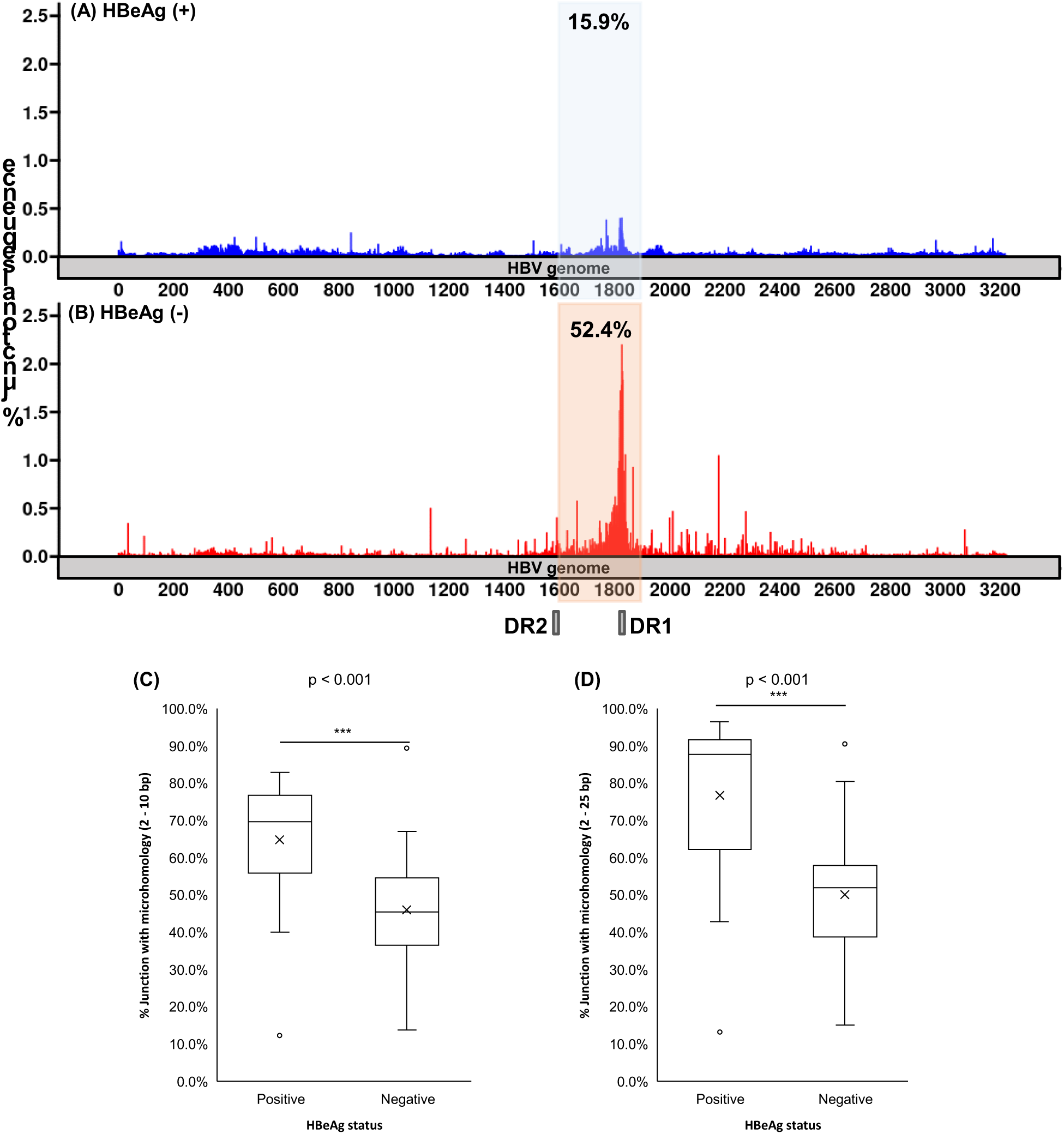
Comparison of the distribution of junction breakpoints (A & B) in the HBV genome and host and viral overlapping sequence homology (C & D) of iDNA between 24 HBeAg(+) and 32 HBeAg(−) CHB. (A, B) X-axis indicates the HBV genome with the nt positions for the DR1 and DR2 noted. Y-axis is the frequency of junction breakpoints of the total junctions. The HBV hotspot breakpoint region of nt 1600-1900 is highlighted: Blue:HBeAg (+) and red: HBeAg (−). Percent of Junction with (C) 2-10 nt or (D) 2-25 nt viral host homologous overlapping sequences. P-value was calculated by Wilcoxon Rank Sum Test (one-tail).

Previous studies indicate that HBV integrations are mainly mediated by the NHEJ mechanism using a full-length dslDNA as the precursor, resulting in breakpoints clustered at the DR2-DR1 region.^28, 29^ We also investigated whether other DNA repair mechanisms such as MMEJ or homologous recombination (HR) are responsible for over 80% of random breakpoints identified especially in the HBeAg(+) participants. An overlapping sequence homology analysis of junction sequences was performed and summarized in **Figure 2C-D**. Interestingly, a significant portion of overlapping junction sequences containing 2-10 or 2-25 nt between viral and host were observed, and such homology is significantly more frequent in the HBeAg(+) group. Moreover, in the HBeAg(+) group, the 2-25 nt overlapping homology frequencies (mean 76.7%) were similar in the breakpoints both within and outside of the DR2-DR1 regions (mean 84.1% vs. 76.6%). In contrast, HBeAg(−) participants had the iDNA breakpoints clustered in the DR2-DR1 region which was known to be associated with HCC (**Figure 2B**). We next examined the frequencies of basal core promoter (BCP) 1762T/1764A mutations and the precore (PC) G1896A stop codon mutation, both are known to lead to reduced and abolished HBeAg expression. BCP mutations were also implicated to increase HCC risk.^31^ (**Figure 3**). As expected, the prevalence of the G1896A mutation was significantly higher in the HBeAg(−) compared to the HBeAg(+) group (34.4% vs. 4.2%, respectively). The prevalence of BCP mutations was slightly higher in the HBeAg(+) group but was not statistically different. There was no HCC identified in this study cohort after 10 years of the follow-up.

**Figure 3.**
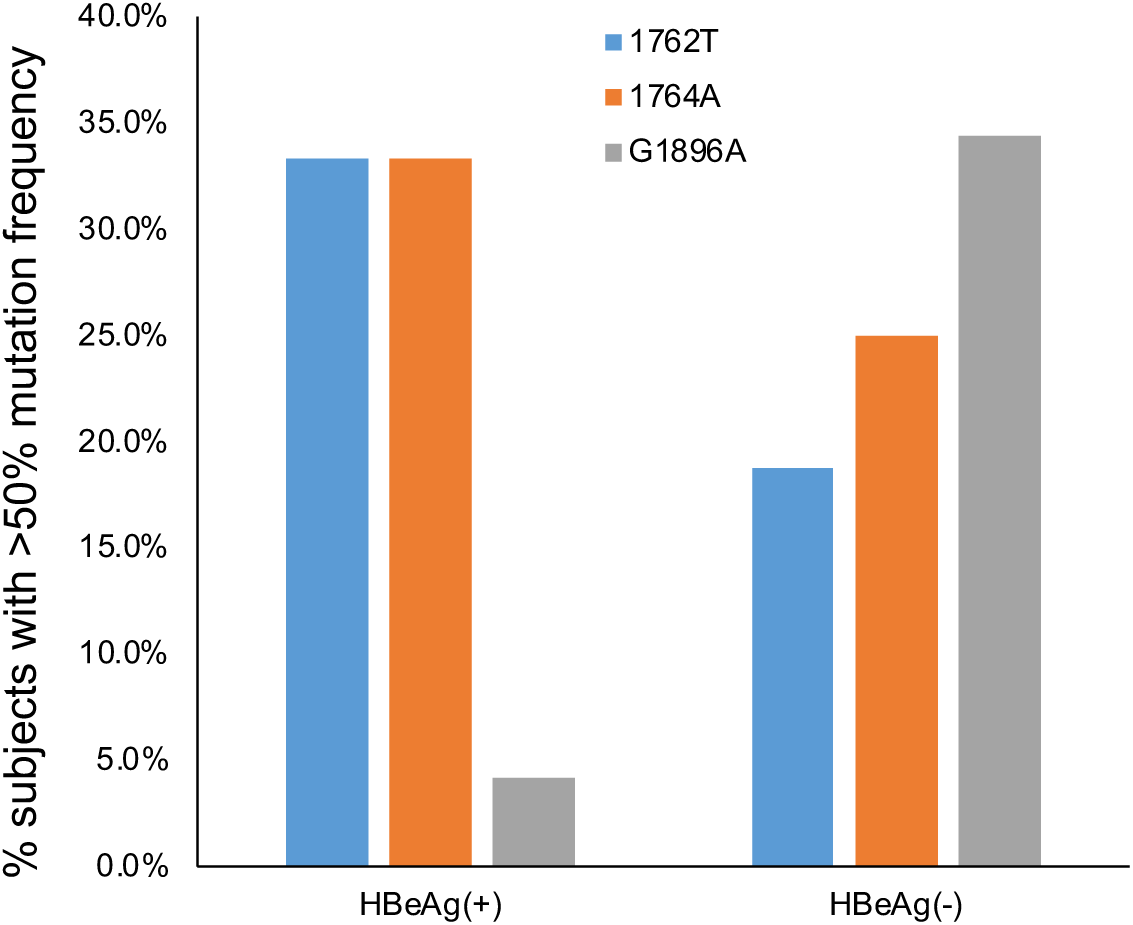
Comparison of the basal core promoter (nt 1762T/1764A) and precore G1896A mutations detected by HBV-targeted NGS assay between HBeAg(+) and HBeAg(−) CHB. Y-axis is the percent of patients contained mutation detected in >50% of NGS reads.

The proportion of the intrahepatic iDNA from the total HBV DNA was calculated as described in Supplemental Figure 1 and 2 in details. By applying this analysis, all but 1 (95.8%) HBeAg(+) participants had ≤50% iDNA in their livers. In contrast, among the HBeAg(−) participants, 25 (78.1%) had >50% iDNA; 2 (6.3%) had 30-50% and 5 (15.7%) had <30% iDNA (**Figure 4**). For both the HBeAg(+) and (−) groups, there was no positive correlation between the levels of iDNA and serum HBV DNA. HBeAg(−) participants with inactive and immune active phases of CHB had similar iDNA levels. HBeAg(−) participants generally had higher % iDNA compared to HBeAg(+) ones regardless of their HBV DNA levels (**Figure 5**).

**Figure 4.**
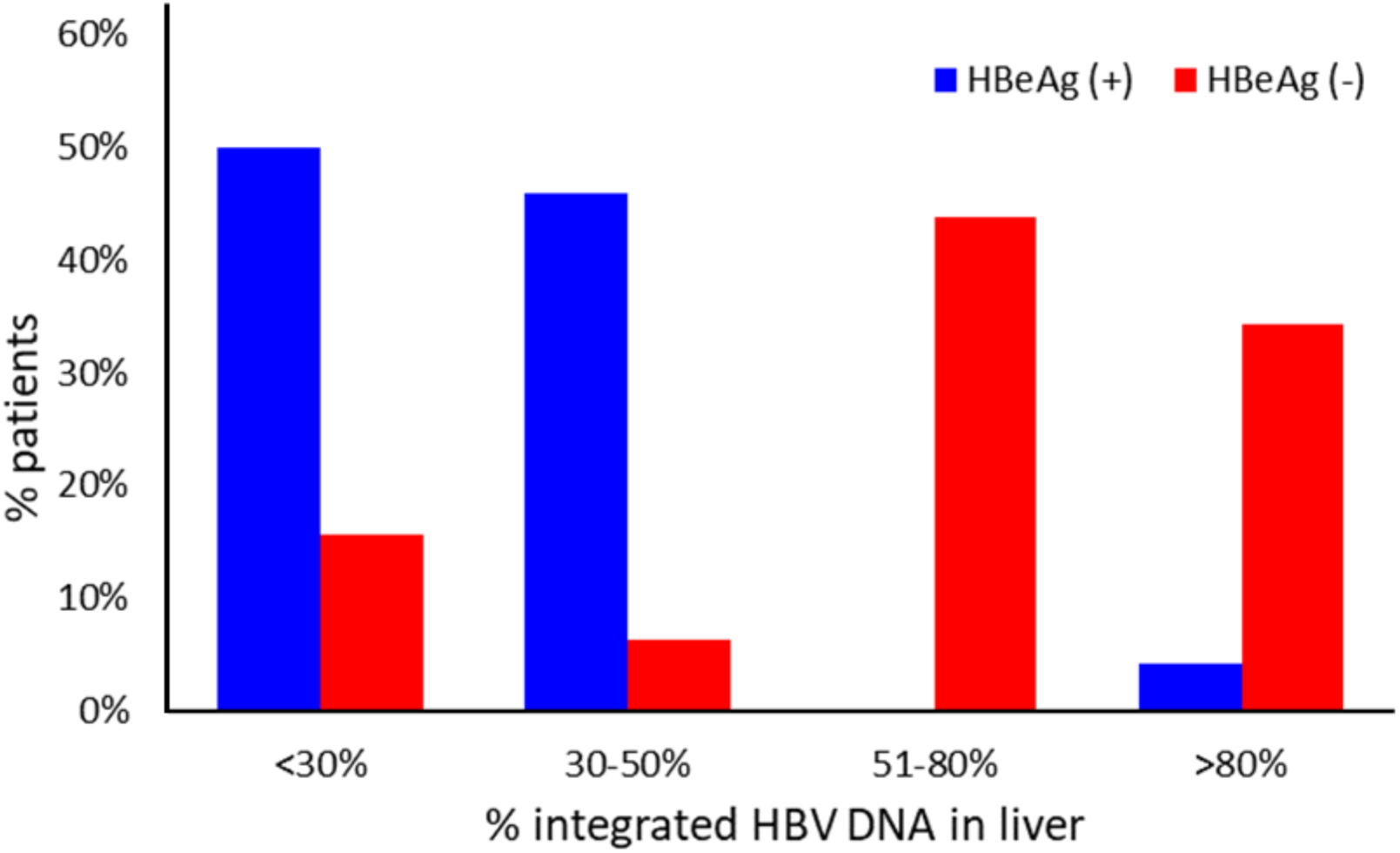
Intrahepatic integrated HBV DNA Levels. Over 95.8 % of HBeAg(+) livers had less than 50% integrated DNA. In contrast, 78.1% of HBeAg(−) livers had >50% integrated DNA.

**Figure 5.**
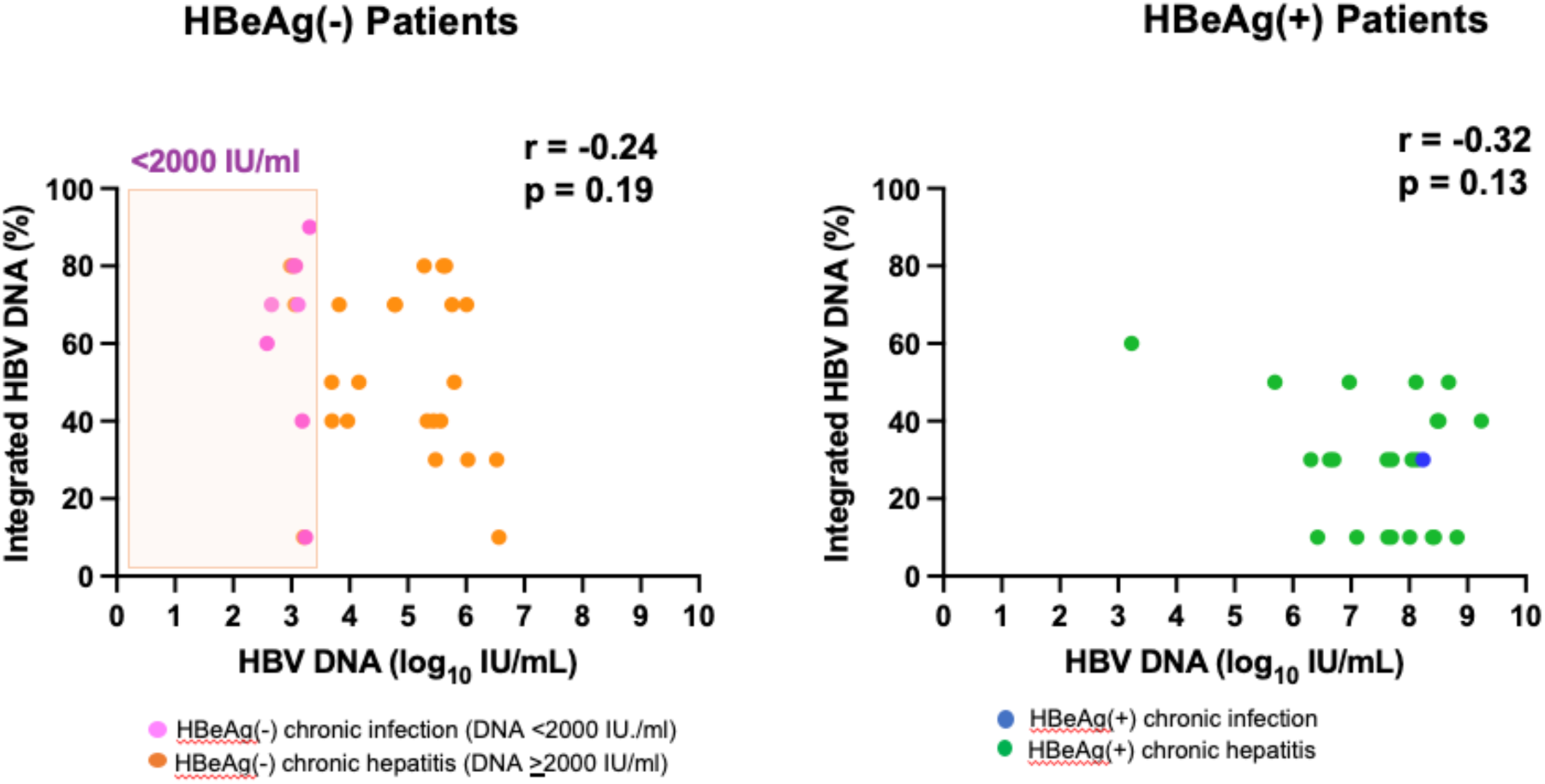
No correlation between integrated HBV DNA in livers and serum HBV DNA levels in either HBeAg(+) or (−) participants.

## DISCUSSION

HBV integration is a universal hallmark of HBV infection and represents another persistent form of viral DNA besides the cccDNA despite being dispensable in the HBV life cycle.^32^ In vitro infection models suggest that HBV integration into the host genome tends occur early following infection, within 3 days after acute hepatitis B and the site appears to be random within the host genome.^33^ During chronic infection, the hepatocytes with iDNA undergo clonal expansion in response to the persistent inflammation.^34, 35^ HBV integration is believed to play a significant role in HCC development.^29, 36–40^ Another important role of iDNA is to maintain viral persistence by providing an additional stable source of immunosuppressive HBsAg protein production.^13^ The current quantitative serum HBsAg assay, however, cannot distinguish between intrahepatic cccDNA and iDNA as the source of HBsAg.

In this predominantly Asian born patient antiviral treatment naïve study cohort with HBV genotypes B and C, the HBeAg(+) participants, as expected, had significantly higher intrahepatic cccDNA, plasma HBV RNA, serum HBV DNA, HBcrAg and qHBsAg compared to the HBeAg(−) ones. Similarly, the majority (88%) of the HBeAg(+) participants had detectable intrahepatic HBcAg staining compared to only 13% among the HBeAg(−) participants. This HBcAg stain pattern likely was the result of higher HBV replication in HBeAg(+) CHB.^41^ While positive cytoplasmic HBsAg staining was noted in similar frequency in HBeAg(+) and HBeAg(−) livers, a membranous HBsAg staining pattern was observed in only 14% of HBeAg(−) compared to 78% of HBeAg(+) samples. Our findings were consistent with prior observations that the cytoplasmic HBsAg stain was common in both HBeAg(+) and (−) CHB regardless of the HBV replication status whereas membranous staining of HBsAg correlated with active HBV replication and high HBV DNA levels.^41^ These intrahepatic findings provided clues that there were differential sources of HBsAg between HBeAg(+) and (−) disease states. The lack of correlation between serum qHBsAg levels and plasma HBV RNA and serum HBV DNA but a significant correlation between qHBsAg and iDNA (HBV-host junctional reads) among HBeAg(−) participants supported the premise that the HBsAg was mostly generated by iDNA in HBeAg(−) CHB.

For the detection of intrahepatic cccDNA, Southern blot remains the “gold standard” for cccDNA visualization; however, it lacks sensitivity and is less quantitative.^42^ While qPCR eliminates these limitations, the presence of HBV rcDNA in total DNA or Hirt DNA extraction can introduce contamination compromising its specificity and accuracy.^43^ Previous studies have employed PSAD to remove rcDNA, but we have noticed that rcDNA is resistant to PSAD due to its circular form.^19^ Alternatively, exonucleases, such as T5 and ExoI/III, etc., can digest rcDNA with open ends.^44, 45^ In a recent effort to standardize the cccDNA qPCR assay within the International Coalition to Eliminate HBV (ICE-HBV) consortium, it was observed that while exonucleases efficiently remove rcDNA, they may also overdigest cccDNA, whereas PSAD was less prone to induce cccDNA overdigestion.^46^ To address these concerns, we introduced a pre-heating step to denature rcDNA into single-stranded DNA, which can then be efficiently degraded by PSAD, leaving cccDNA as the sole viral DNA template for qPCR detection.^19, 20, 47^ Since a standard cccDNA qPCR assay is not universally available, researchers may choose the rcDNA-eliminating nuclease and method at their discretion to clean up DNA samples for cccDNA qPCR. However, maintaining consistency in the choice of enzyme and protocol is crucial to ensure the reliability of relative quantification of cccDNA throughout the study.

Multiple methods have been developed to detect iDNA.^10^ The major advantages of the NGS technology include its capacity to sequence millions of reads in a single run and its independence from prior knowledge regarding the HBV or host gene sequences. This minimizes potential selection biases. For iDNA quantitation using the ChimericSeq output,^22^ we determined the relationship between HBV-JS NGS reads and HBV-JS specific qPCR using stringent controls to eliminate false positive background reads.^21^ By using this method, we were able to compare the percentages of iDNA across the liver samples. In agreement with the previous publication,^12^ HBeAg(−) CHB samples had significantly higher fraction of iDNA compared to HBeAg(+) ones (**Figure 4**). In fact, over 70% of the HBeAg(−) participants had >50% iDNA among total HBV DNA; in contrast, 96% of HBeAg(+) participants had ≤50% iDNA in their livers. Similar to the qHBsAg profile, there was no correlation between iDNA and serum HBV DNA levels (**Figure 5**). It is worth noting that patients with HBeAg(−) CHB generally had longer disease duration with lower viral replication compared to those with HBeAg(+) CHB. While the HBV DNA integration rates might graduately diminish in HBeAg(−) CHB, the fraction of iDNA likely increased due to its survival with the selective infected hepatocytes over time.

Most strikingly, for the HBeAg(−) group, 52.4% of the junctional sequence breakpoints clustered at nucleotide (nt) positions 1600 −1900 around the DR2-DR1 region; however, the distribution of junctional breakpoints for the HBeAg(+) group was more random across the HBV genome with only 15.9% concentrated in the corresponding DR2-DR1 region (**Figure 2A**). The significant differences in the breakpoint distribution of iDNA suggest that the HBeAg status may influence iDNA stability, mobility, or HCC pathogenesis. For HBeAg(+) group, active viral replication facilitated integrations through promoting host cell double strand DNA breaks and providing more viral DNA integration precursors, thus the iDNA breakpoints represented more random integration breakpoints, as demonstrated by their high frequency of MMEJ, without selection of iDNA survival (**Figure 2B**). An illustration of the interpretation for the distinct iDNA patterns between the HBeAg(+) and HBeAg(−) CHB is presented in **Figure 6**. Given an overall high level of short-patch homology of HBV genome to the host especially among the HBeAg (+) participants, MMEJ or even homologous recombination may play a role in HBV integration in addition to the well-known NHEJ mechanism.^29, 32, 36^ In contrast, in HBeAg(−) CHB, the identified iDNAs were likely selected during the early stage of active viral replication which survived the elimination of infected cells by the host immune response and underwent clonal expansion during the course of chronic infection. Thus, their breakpoints clustered in the DR2-DR1 hotspots likely resulted from both targeted selection over the prolonged disease course, preferential integration and expansion. The integration breakpoints located in the DR2-DR1 region contain HBV enhancer II and core promoter(EnhII/Cp) which likely favor the selection of those hepatocytes in which HBV integrations have conferred a selective advantage; namely, those have escaped from cytotoxic immune response by activating specific host genes with relevant functions that may favor clonal expansion, a hallmark of carcinogenesis.^28^ The observation of integration breakpoints clustered in the DR2-DR1 hotspots was originally discovered in HBV-induced HCC.^48, 49^

**Figure 6.**
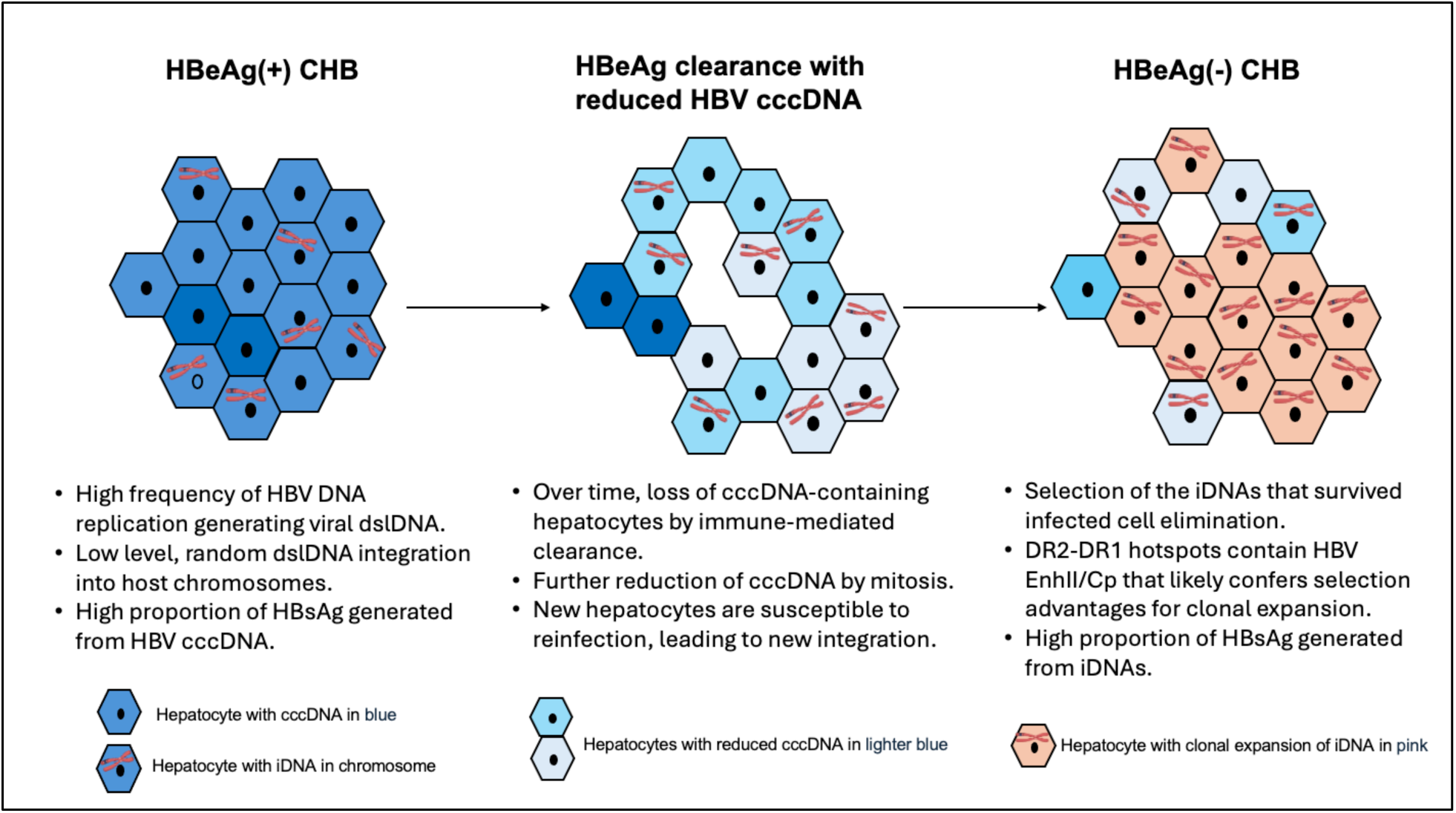
A potential model of the differential iDNA patterns from HBeAg(+) to HBeAg(−) phase of chronic hepatitis B

With prolonged nucleos(t)ide therapy, a number of studies reported reduction in HBV cccDNA and iDNA levels.^50, 51^ In functional cure or HBsAg clearance, the intrahepatic cccDNA is expected to be in a transcriptionally inactive state even if cccDNA persists. On the other hand, detectable HBsAg from transcriptionally active iDNA can persist despite cccDNA inactivation or elimination. In a recent publication, Gao et al^51^ analyzed liver biopsy samples from 47 participants who achieved functional cure after pegylated interferon (Peg-IFN) therapy and reported that none of these participants had positive HBcAg stain in liver tissues. However, 25.5% of the participants still had positive HBsAg stain; it did not mention whether the HBsAg was predominantly cytoplasmic or membranous in distribution. Intrahepatic HBV cccDNA and RNA were detectable in 68.1% and 72.2% respectively. Similar to our findings in untreated HBeAg(−) CHB, the peak junctional reads were detectable in the DR1 region. More in depth analyses would be necessary to determine whether the RNA transcripts were derived from cccDNA and if the detectable cccDNA was transcriptionally active. Relapse or reactivation of hepatitis B would be likely if the cccDNA was still transcriptionally active despite undetectable HBsAg in serum. The discrepancy in the detectability of HBsAg in livers and blood could be due to the sensitivity of the assay in detecting low levels of HBsAg in serum, or the HBsAg could be bound to the anti-HBs to form the immune complexes in blood.

Detecting changes in cccDNA concentration and transcriptional activities without the need for repeat liver biopsies is desirable to monitor treatment response. Serum HBV RNA and HBcrAg have been applied as surrogate cccDNA markers. These markers, however, have not been fully standardized; moreover, their sensitivities and specificities need to be optimized.^15, 16, 26, 52^ Another limitation of the current HBcrAg assay is its inability to distinguish between HBcAg, HBeAg and precore proteins.^15, 53^

Novel antivirals currently in development aim to eliminate cccDNA and iDNA.^4, 32, 54, 55^ Some patients might have no expression of HBsAg from cccDNA after prolonged antiviral therapy but continue to have detectable HBsAg derived from iDNA. It is highly desirable to have a liquid biopsy test that can detect iDNA to monitor the efficacy of therapy and to identify patients who have achieved or likely will achieve functional cure.

## Data Availability

Study materials will be made available to other researchers. Data will be open-access and available publicly after patent filing and publication.

## ACKNOWLEGEMENT

DEK: This study was supported, in part, by the Intramural Research Program of the National Institutes of Health, National Cancer Institute.

