## Supplemental Table 1 for "Differential Intrahepatic Integrated HBV DNA Patterns Between HBeAg-Positive and HBeAg-Negative Chronic Hepatitis B"

**Supplemental Table 1.** **List of oligonucleotides used in the study.**

| **Target** | **Primer and probe sequences (5’→3’)** | |
| --- | --- | --- |
| HBV cccDNA | Forward | GTCTGTGCCTTCTCATCTGC (nt 1553-1572) |
|  | Reverse | AGTAACTCCACAGWAGCTCCAAATT (nt1949-1925) |
|  | Probe | FAM-TTCAAGCCTCCAAGCTGTGCCTTGGGTGGC-TAMRA (nt 1865-1894) |
| Cytochrome c oxidase subunit III (COX3, mitochondrial) | Forward | CCCTCTCGGCCCTCCTAATAACCT**GC** |
|  | **Reverse** | GCCTTCTCGTATAACATCGCGTCA |
| Hemoglobin subunit β (HBB) | TaqMan^™^ Gene Expression Assay (FAM), with exact sequences of the primers and probe undisclosed (ThermoFisher Scientific, Cat # 4351370, Hs00758889_s1) | |
