## Supplemental Figure 1 for "Differential Intrahepatic Integrated HBV DNA Patterns Between HBeAg-Positive and HBeAg-Negative Chronic Hepatitis B"

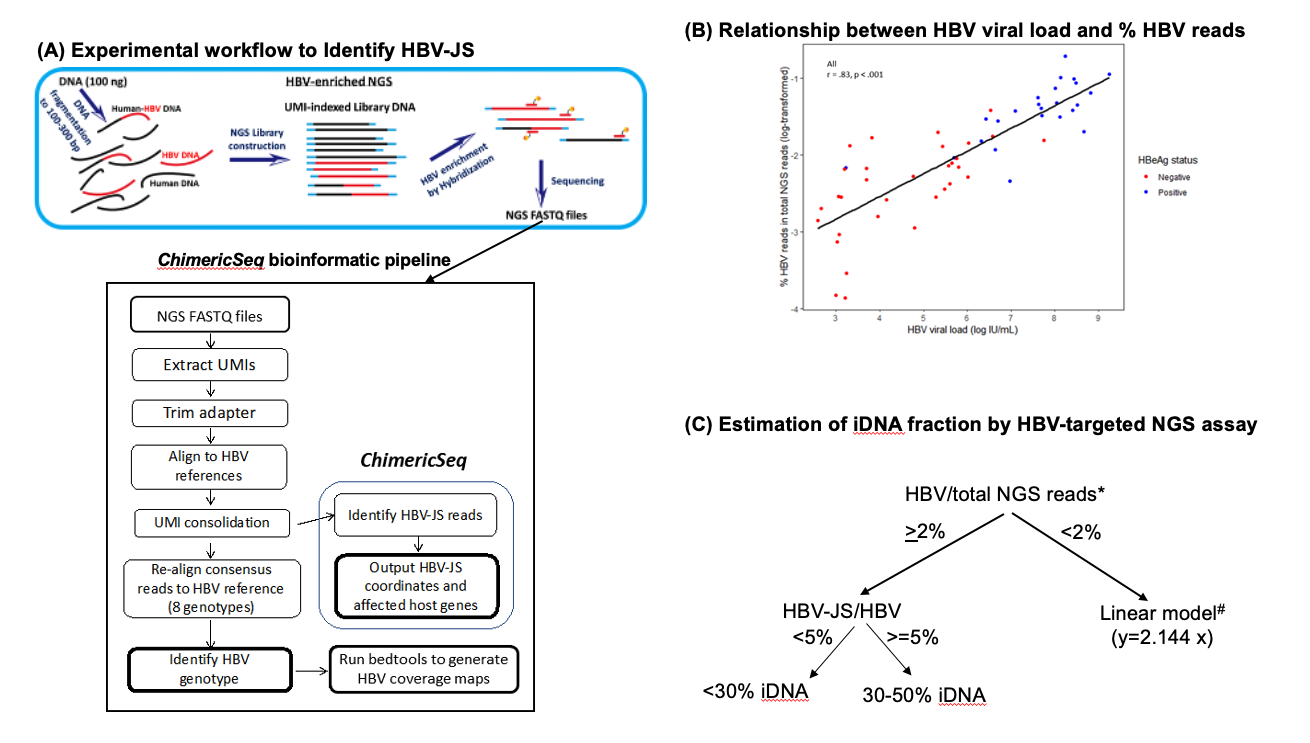

**Intrahepatic iDNA characterization and quantity estimation.** iDNA fraction was estimated and categorized into 4 categories, <30%, 30-50%, 51-80%, and >80%, as illustrated. **(A)** Experimental and bioinformatic workflow to identify HBV-JS. **(B)** Relationship between HBV viral load (HBV DNA level in log IU/mL) and percentage of HBV reads in total NGS reads (log-transformed). The HBV viral load positively correlates to the percentage of HBV reads in total NGS reads among all samples (Pearson’s correlation coefficient = 0.83, p < 0.001). HBeAg(+) samples are plotted in blue, and HBeAg(-) in red. **(C)** A flowchart illustration of the process of estimating the integrated DNA (iDNA) fraction in tissue biopsy samples using a previously developed linear model (MedRxiv) for patients with serum viral loads < 4 log IU/mL. Among 56 samples analyzed with the HBV-NGS assay, all patients with serum viral loads below 4 log IU/mL had HBV reads comprising <2% of total NGS reads, thus qualifying these samples for iDNA fraction estimation using the linear model. For samples with ≥2% HBV/total NGS reads, iDNA was estimated to be under 50% fraction. A threshold of 5% HBV-JS/total HBV reads was used to categorize iDNA fractions as either <30% or 30–50%. The 5% as the threshold is based on the followings. If the entire HBV genome is integrated, with an average NGS read length of ~145 bp, one iDNA molecule would generate 22 NGS reads. In an ideal situation, among these, 2 will be detected as HBV-JS reads, equating to ~10% HBV-JS. This percentage was halved to 5%, determined by the experimental biases, including the removal of junction reads with <20 nt of HBV sequences by the bioinformatics pipeline and reduced hybridization affinity of host-sequence-containing junction fragments to the HBV probes (data not shown) and accommodating the presence of various forms of HBV DNA in unknown proportions, which arise in samples with ongoing viral replication. **(D)** The HBV DNA and JS breakpoint coverage maps generated from tissue DNA of 56 patients. For each coverage map, top panel is the HBV DNA coverage of the HBV NGS read distributions and depth (number of reads) across the entire HBV genome (in black) as to the coverage map reference generated from a monomeric full-length HBV DNA of respected genome type (in red). The median of the coverage depth of each sample with the lowest and the highest HBV DNA coverages are noted in blue. The bottom panel is the HBV-JS breakpoint distribution and coverage map. The total UMI-consolidated HBV read numbers are noted on the top left coner of each chart.
