## Supplemental Figure 2 for "Differential Intrahepatic Integrated HBV DNA Patterns Between HBeAg-Positive and HBeAg-Negative Chronic Hepatitis B"

### Supplemental Figure 2 file. iDNA determination for each of the 56 subjects

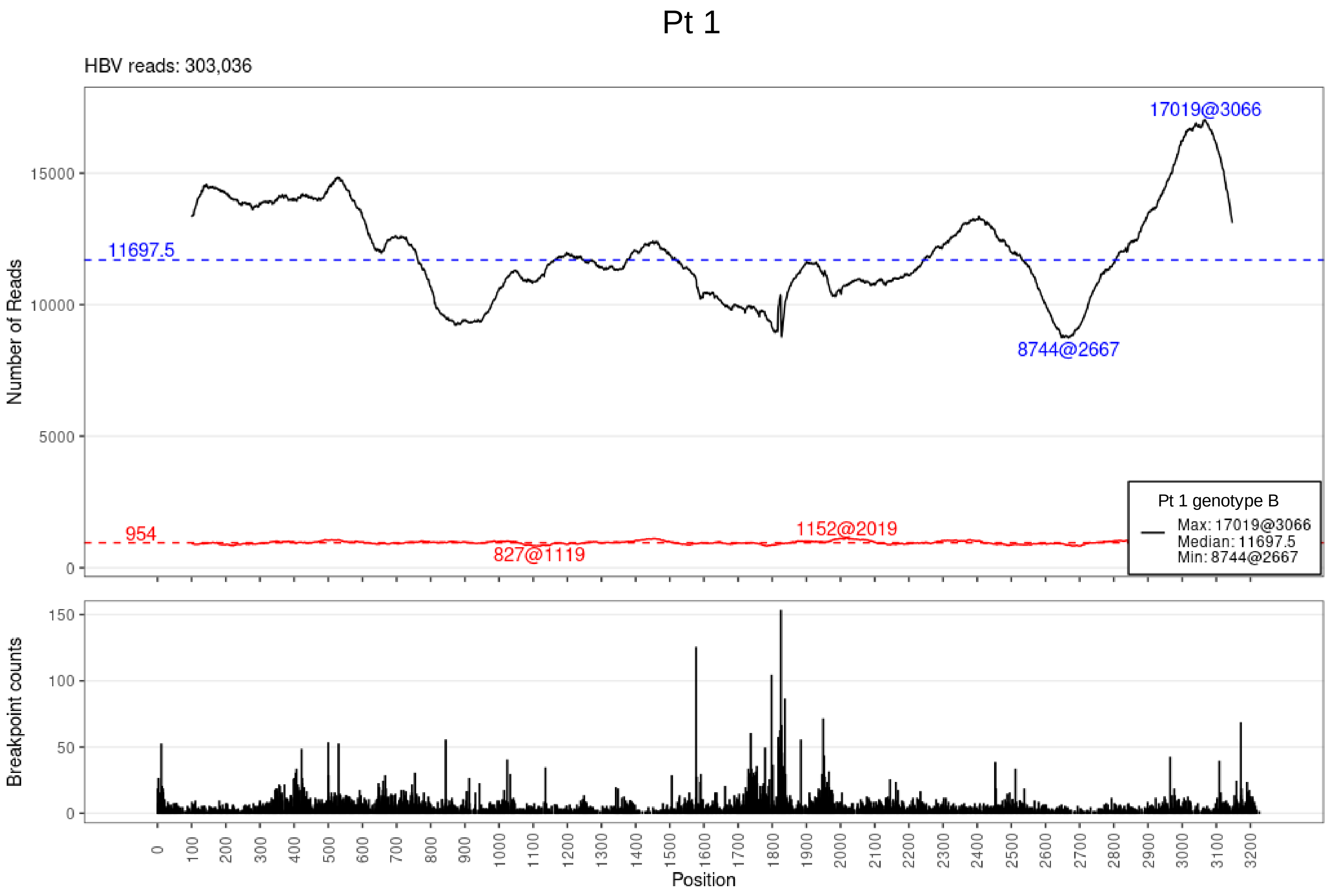

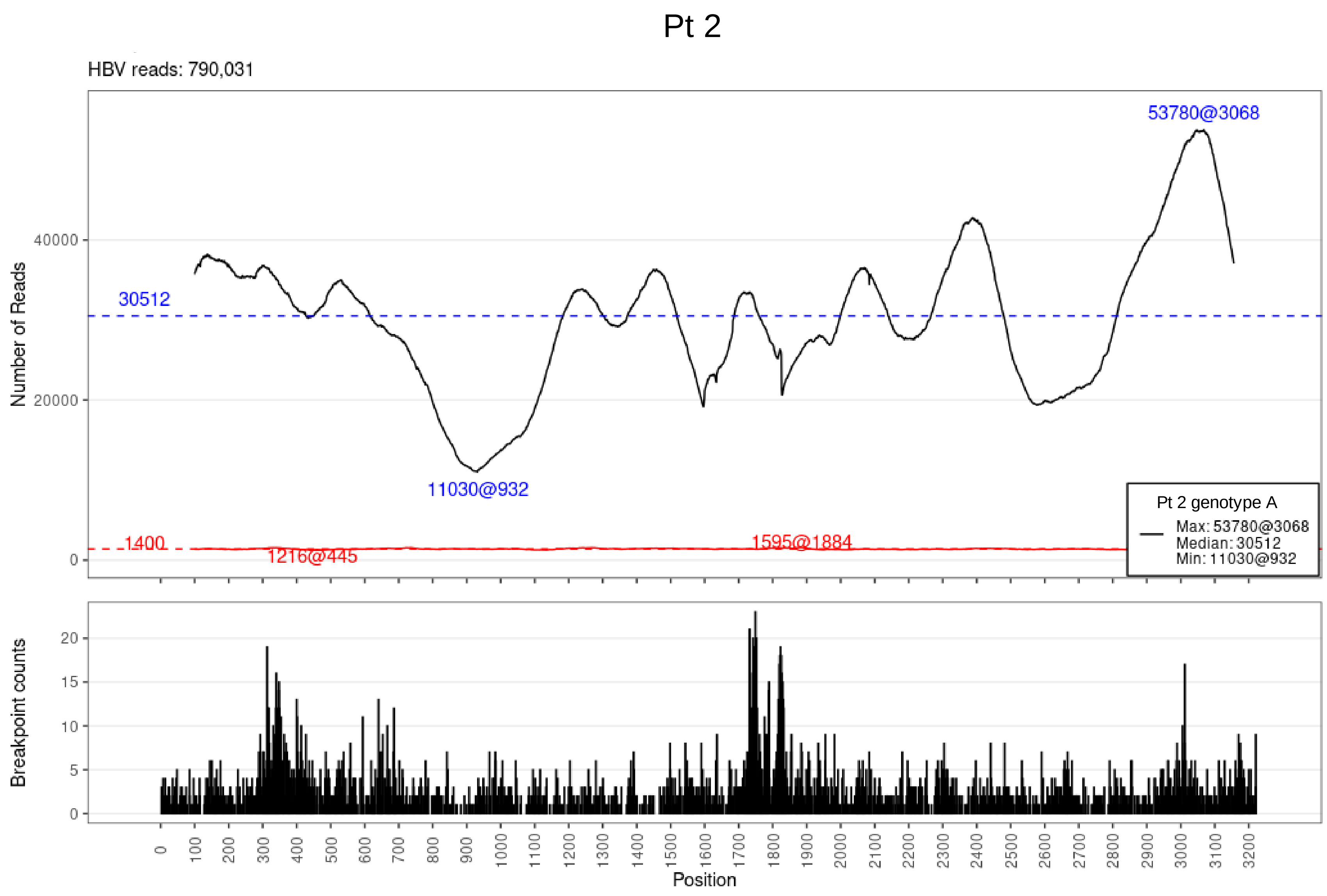
Pt 2
HBV reads: 790,031
53780@3068
30512
11030@932
1400
1216@445
1595@1884
Pt 2 genotype A
Max: 53780@3068
Median: 30512
Min: 11030@932
Number of Reads
Breakpoint counts
Position

## Pt 3

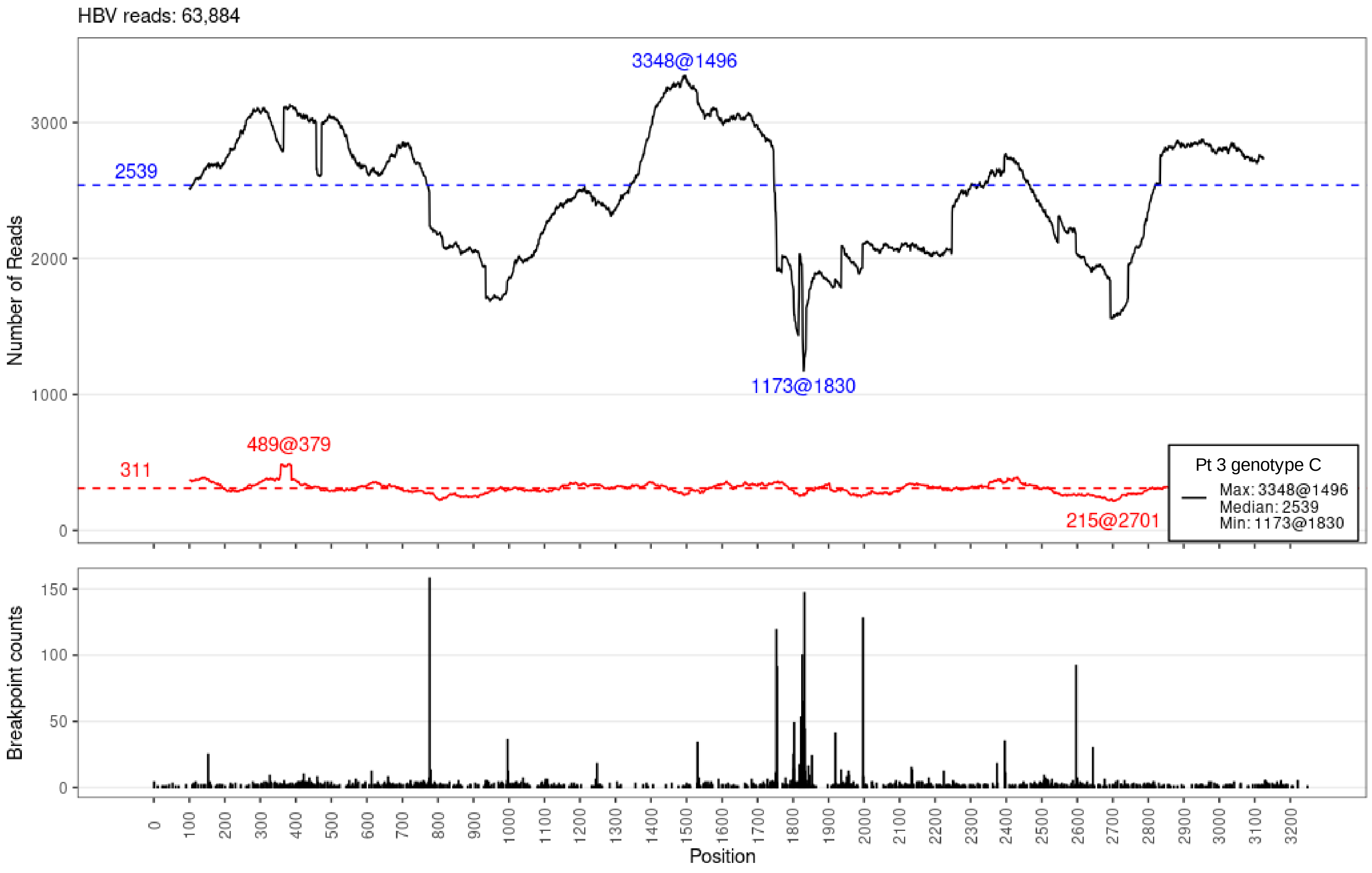

HBV reads: 63,884
3348@1496
2539
1173@1830
489@379
311
215@2701
Pt 3 genotype C
Max: 3348@1496
Median: 2539
Min: 1173@1830
Number of Reads
Breakpoint counts
Position

# Pt 4

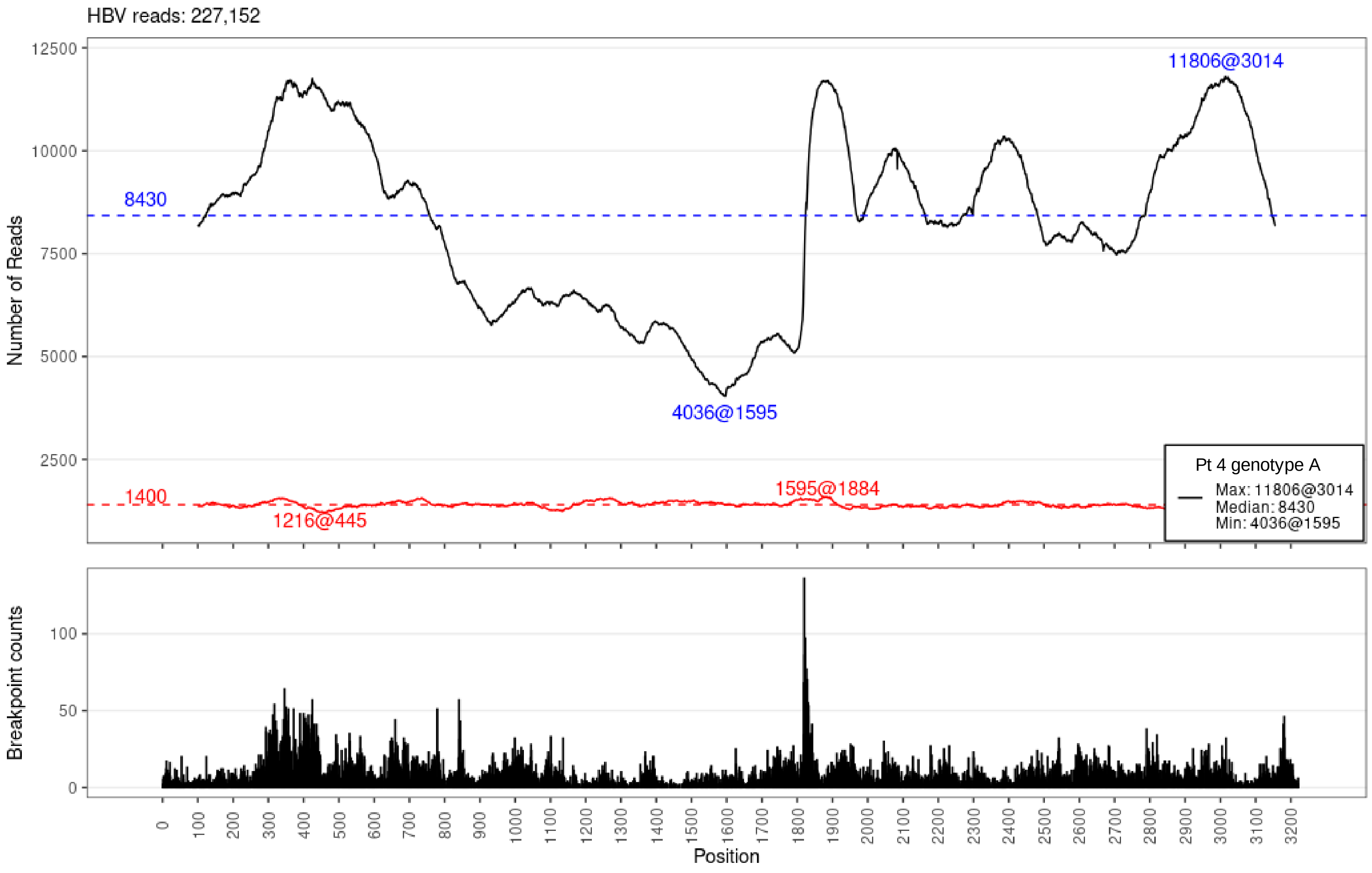

HBV reads: 227,152
Number of Reads
12500
10000
7500
5000
2500
8430
11806@3014
4036@1595
1400
1595@1884
1216@445
Pt 4 genotype A
Max: 11806@3014
Median: 8430
Min: 4036@1595
Breakpoint counts
100
50
0
0
100
200
300
400
500
600
700
800
900
1000
1100
1200
1300
1400
1500
1600
1700
1800
1900
2000
2100
2200
2300
2400
2500
2600
2700
2800
2900
3000
3100
3200
Position

# Pt 5

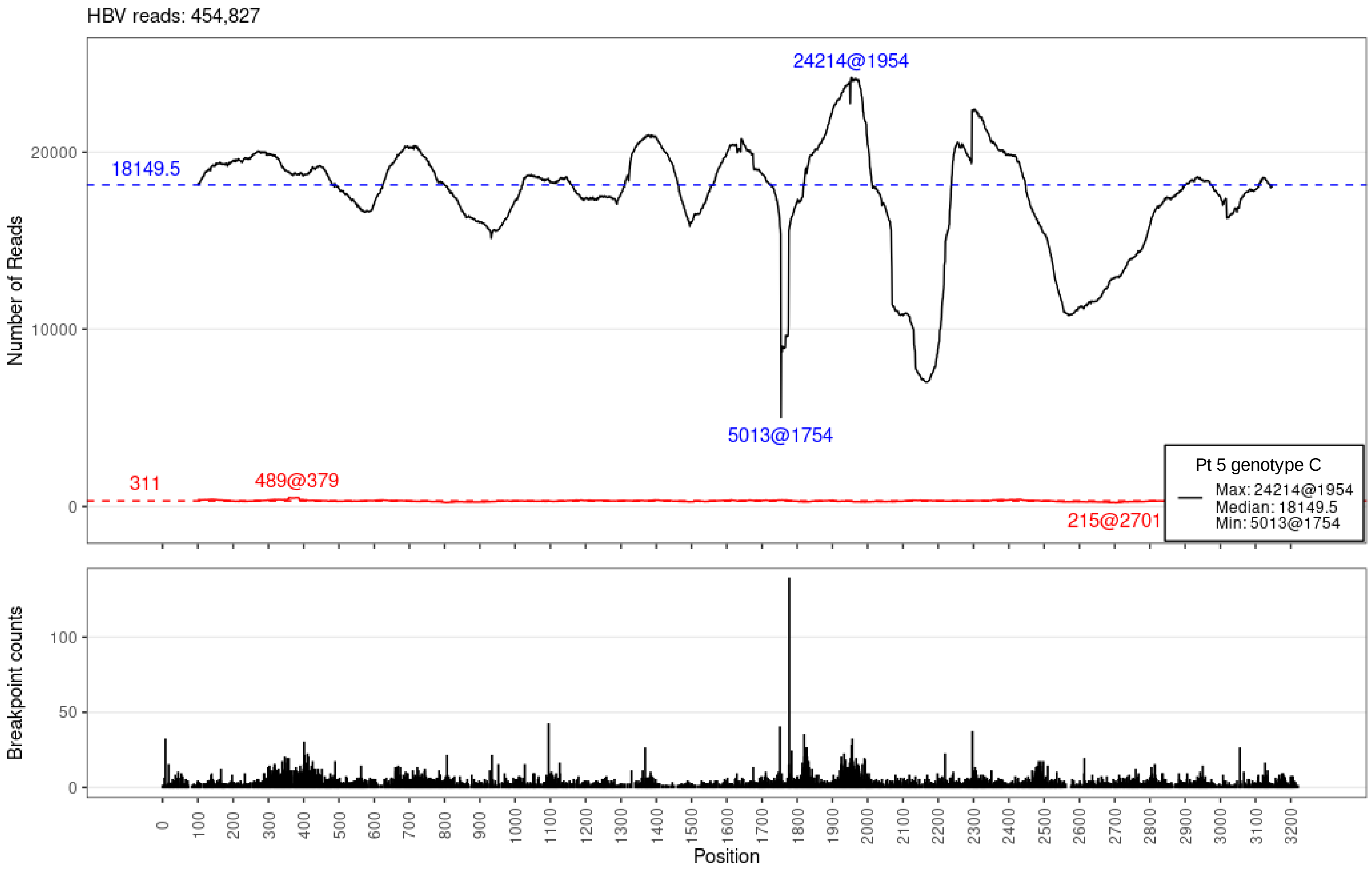
HBV reads: 454,827
24214@1954
18149.5
5013@1754
311
489@379
215@2701
Pt 5 genotype C
Max: 24214@1954
Median: 18149.5
Min: 5013@1754
Number of Reads
Breakpoint counts
Position

Pt 6

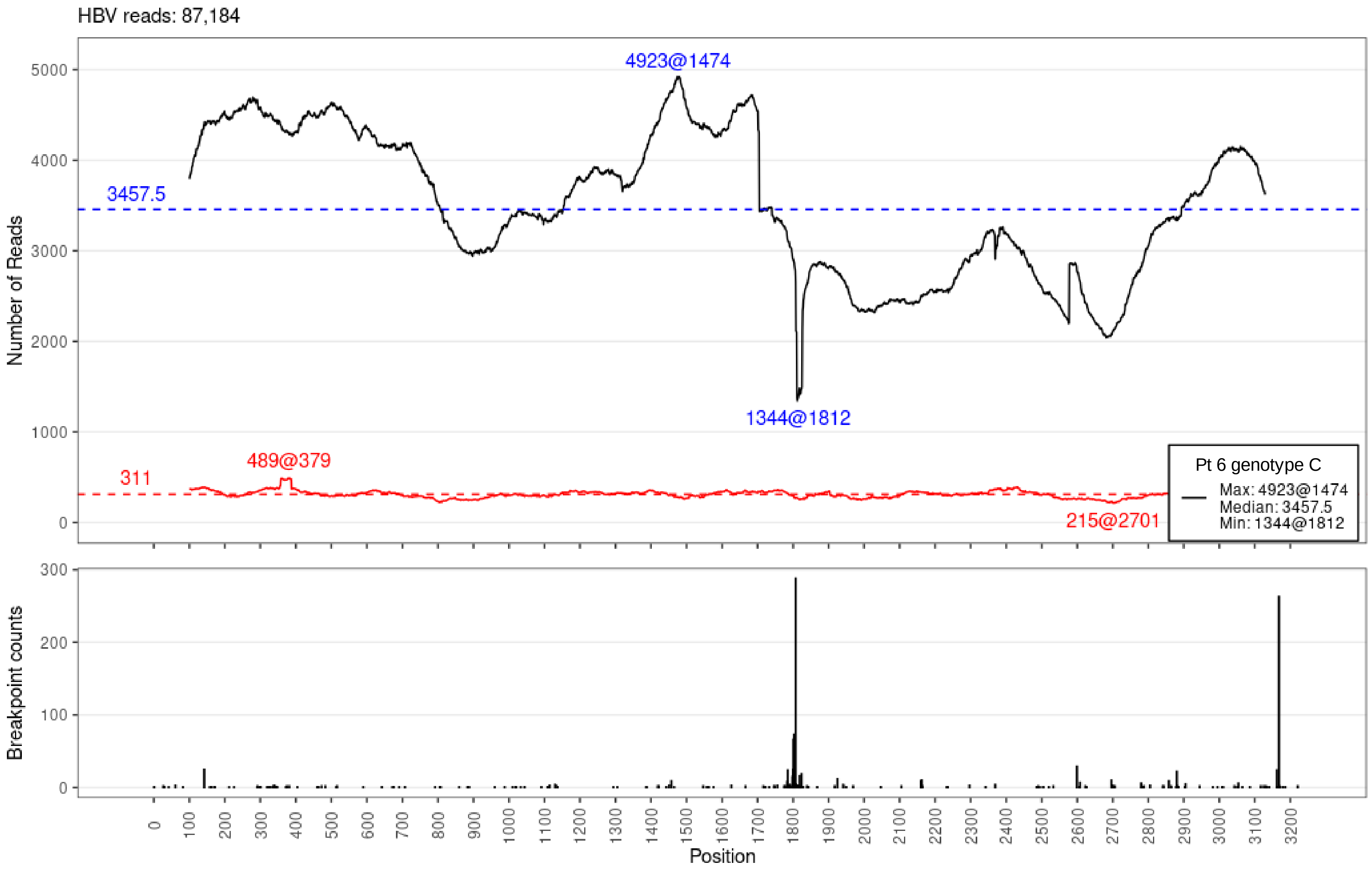
HBV reads: 87,184
4923@1474
3457.5
1344@1812
489@379
311
215@2701
Pt 6 genotype C
Max: 4923@1474
Median: 3457.5
Min: 1344@1812
Number of Reads
Breakpoint counts
Position

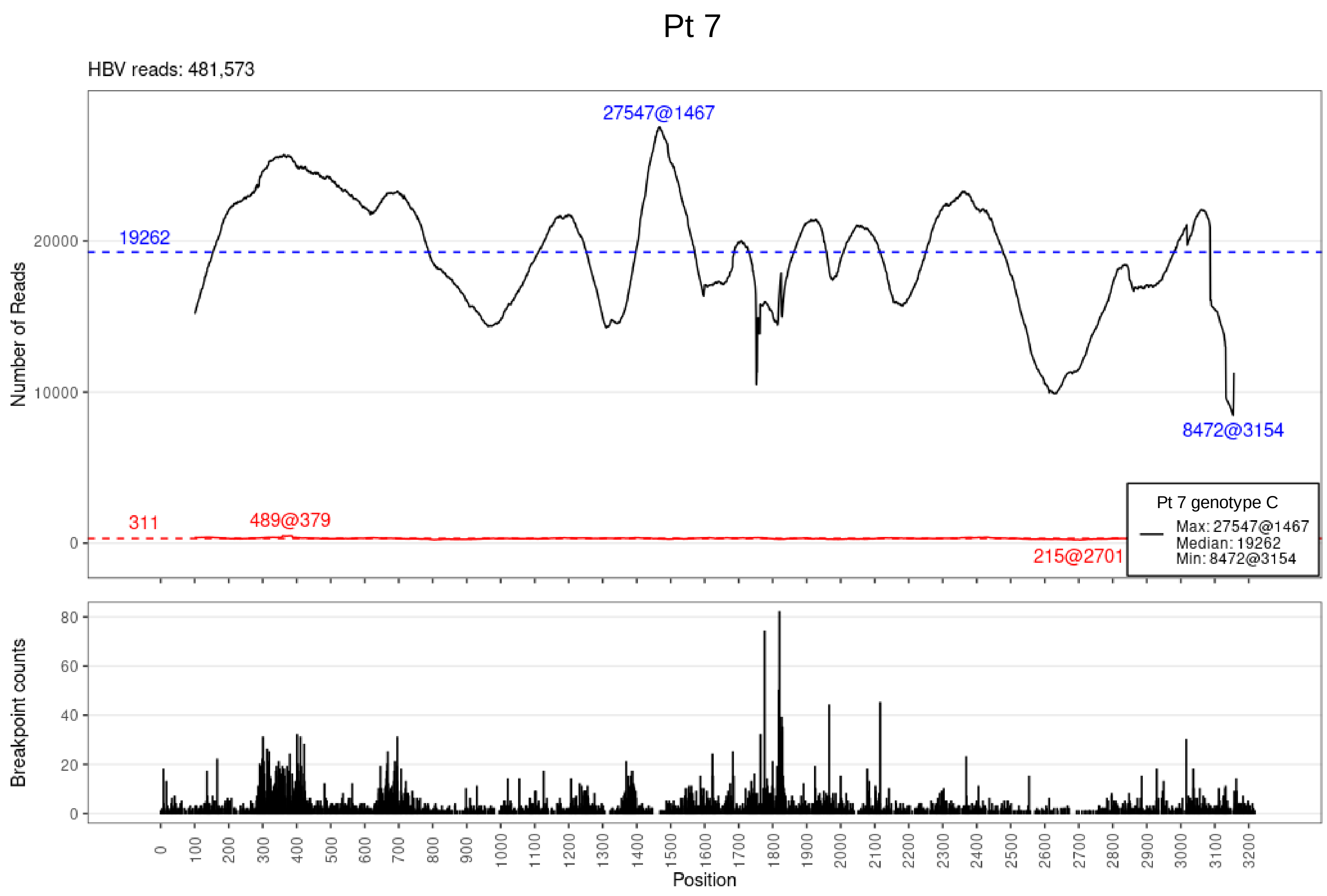

Pt 7
HBV reads: 481,573
27547@1467
19262
8472@3154
311
489@379
215@2701
Pt 7 genotype C
Max: 27547@1467
Median: 19262
Min: 8472@3154
Number of Reads
Breakpoint counts
Position

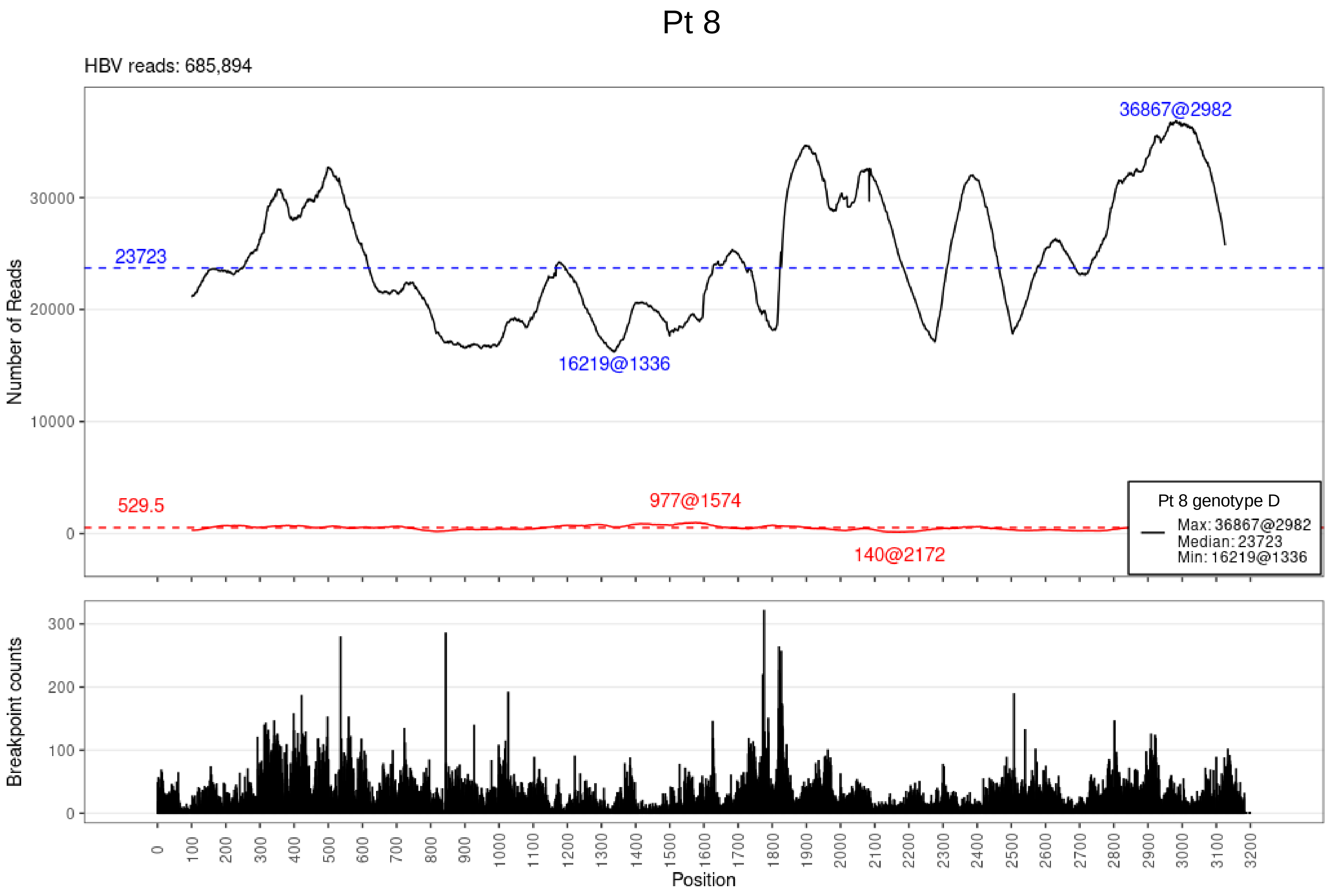

Pt 8
HBV reads: 685,894
36867@2982
23723
16219@1336
529.5
977@1574
140@2172
Pt 8 genotype D
Max: 36867@2982
Median: 23723
Min: 16219@1336
Number of Reads
Breakpoint counts
Position

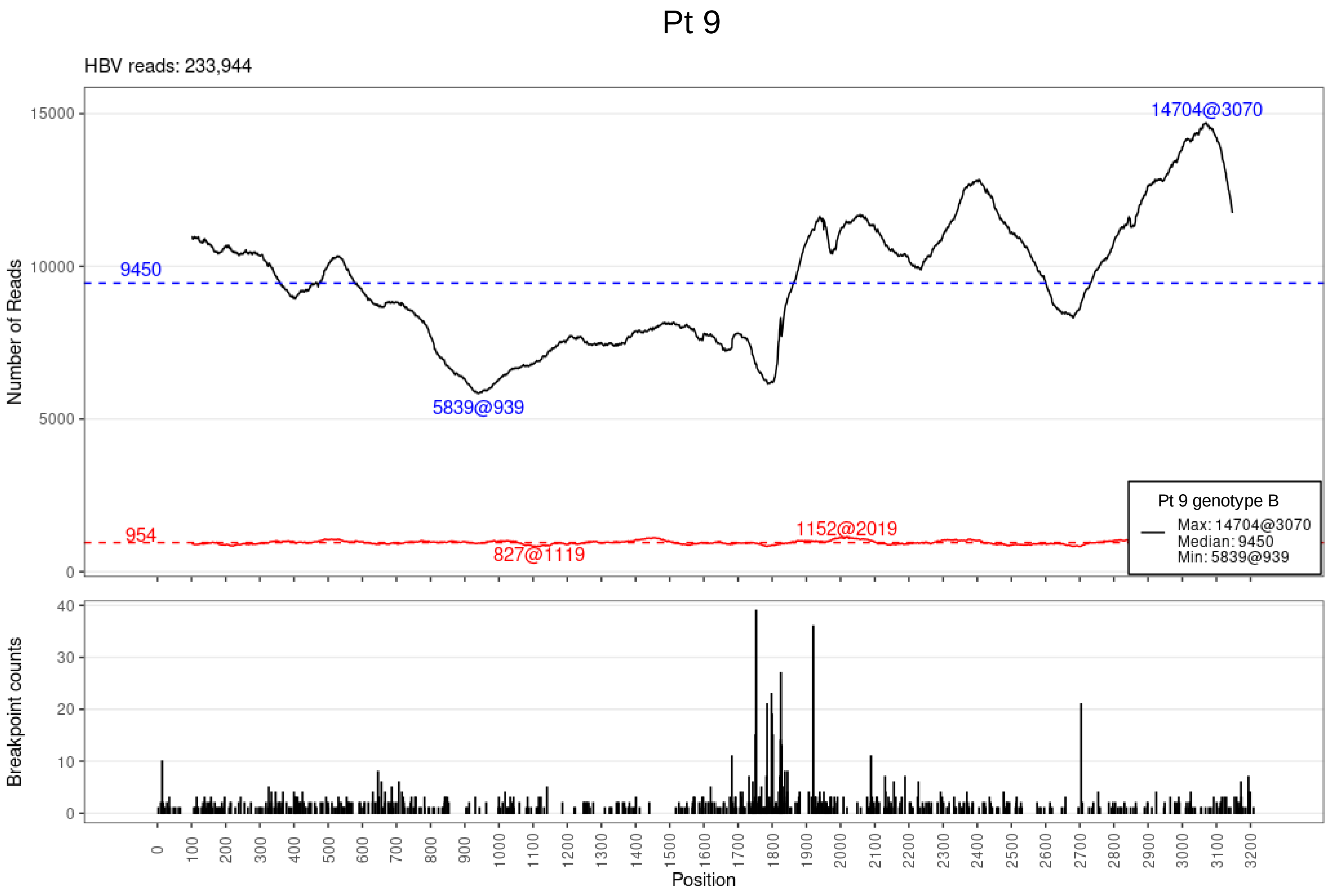

Pt 9
HBV reads: 233,944
Number of Reads
15000
10000
5000
0
9450
954
14704@3070
5839@939
1152@2019
827@1119
Pt 9 genotype B
Max: 14704@3070
Median: 9450
Min: 5839@939
Breakpoint counts
40
30
20
10
0
0 100 200 300 400 500 600 700 800 900 1000 1100 1200 1300 1400 1500 1600 1700 1800 1900 2000 2100 2200 2300 2400 2500 2600 2700 2800 2900 3000 3100 3200
Position

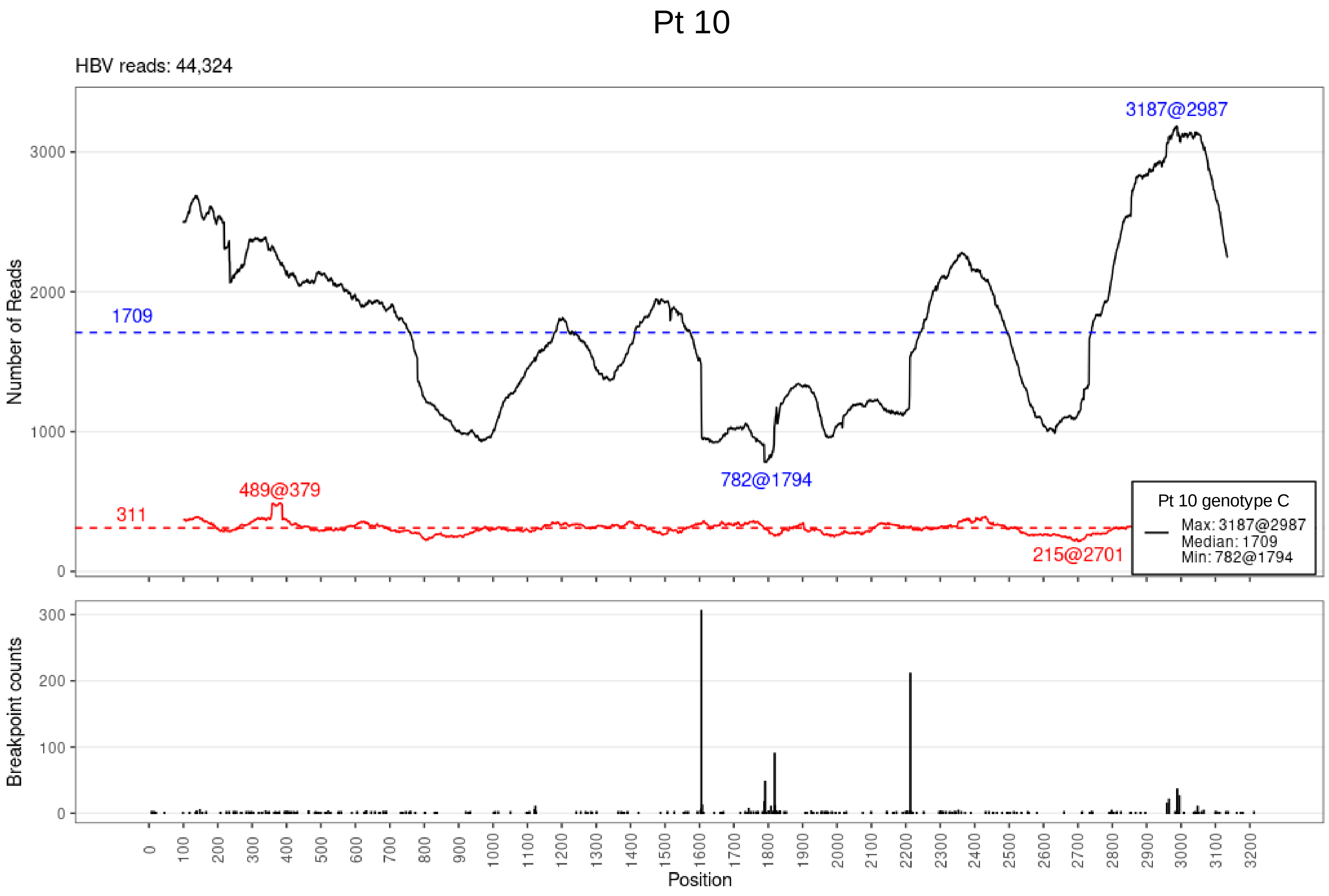

Pt 10
HBV reads: 44,324
3187@2987
1709
782@1794
489@379
311
215@2701
Pt 10 genotype C
Max: 3187@2987
Median: 1709
Min: 782@1794
Number of Reads
Breakpoint counts
Position

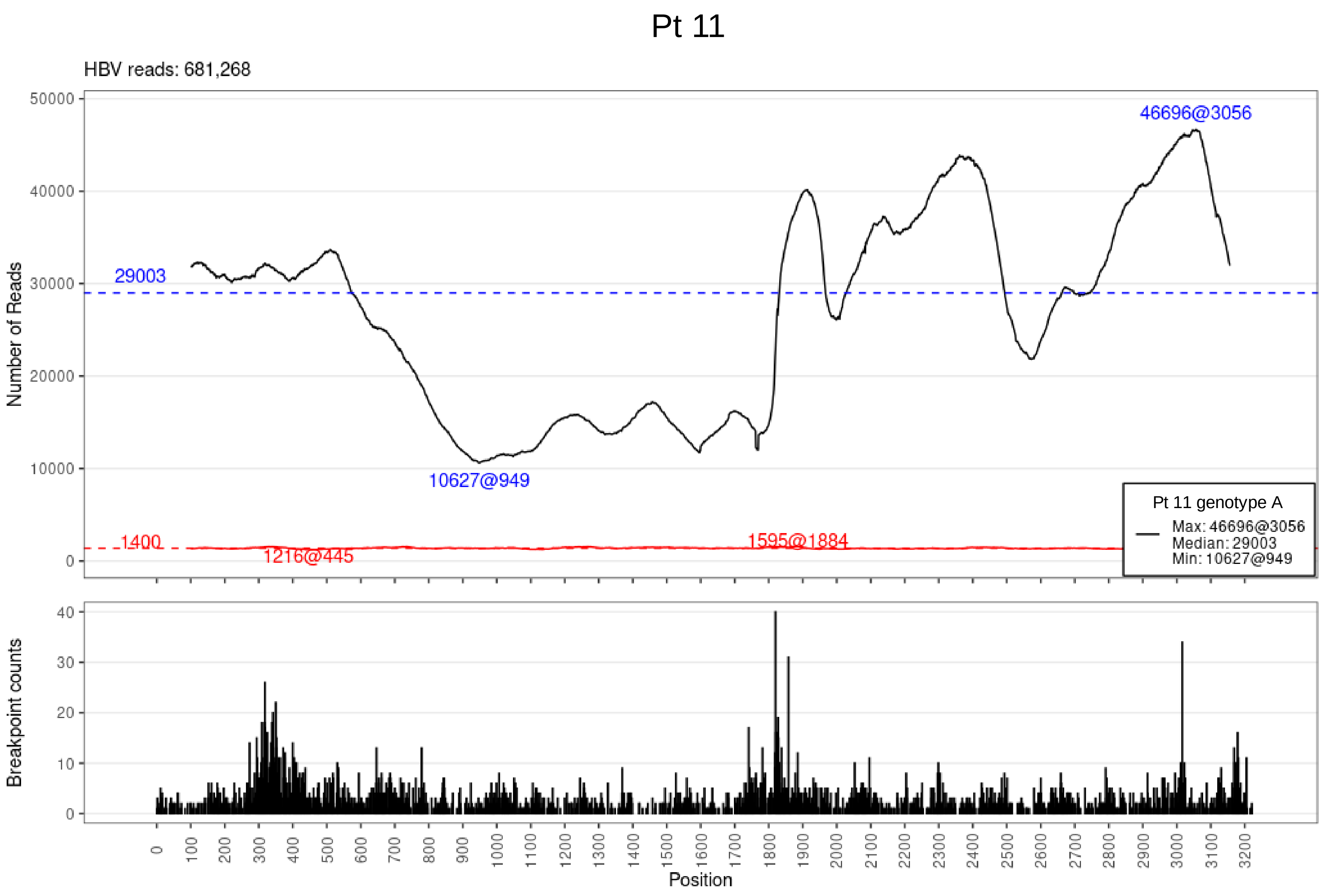

Pt 11
HBV reads: 681,268
Number of Reads
46696@3056
29003
10627@949
1400
1216@445
1595@1884
Pt 11 genotype A
Max: 46696@3056
Median: 29003
Min: 10627@949
Breakpoint counts
Position

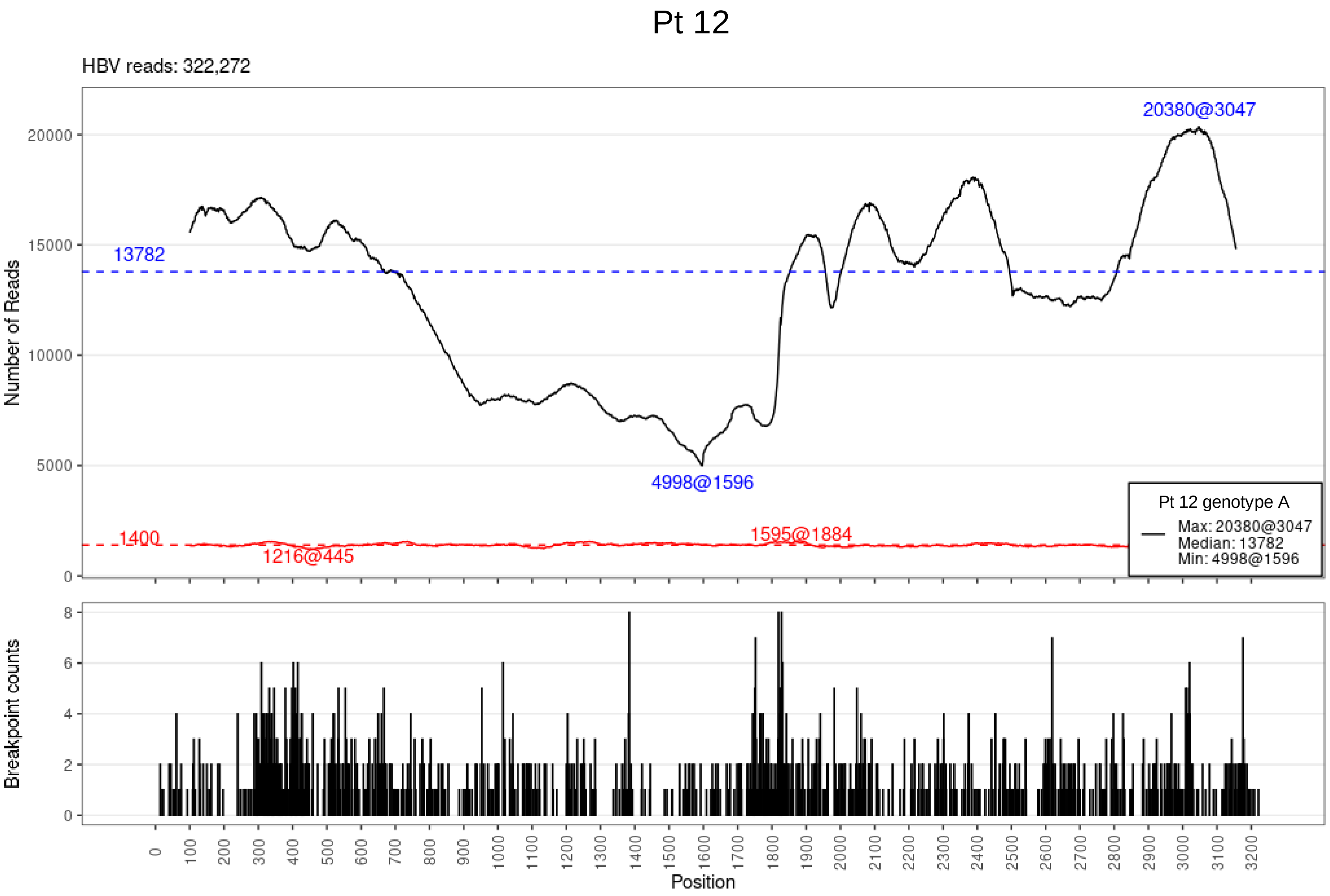

Pt 12
HBV reads: 322,272
Number of Reads
20000
15000
10000
5000
0
13782
1400
20380@3047
4998@1596
1216@445
1595@1884
Pt 12 genotype A
Max: 20380@3047
Median: 13782
Min: 4998@1596
Breakpoint counts
8
6
4
2
0
0 100 200 300 400 500 600 700 800 900 1000 1100 1200 1300 1400 1500 1600 1700 1800 1900 2000 2100 2200 2300 2400 2500 2600 2700 2800 2900 3000 3100 3200
Position

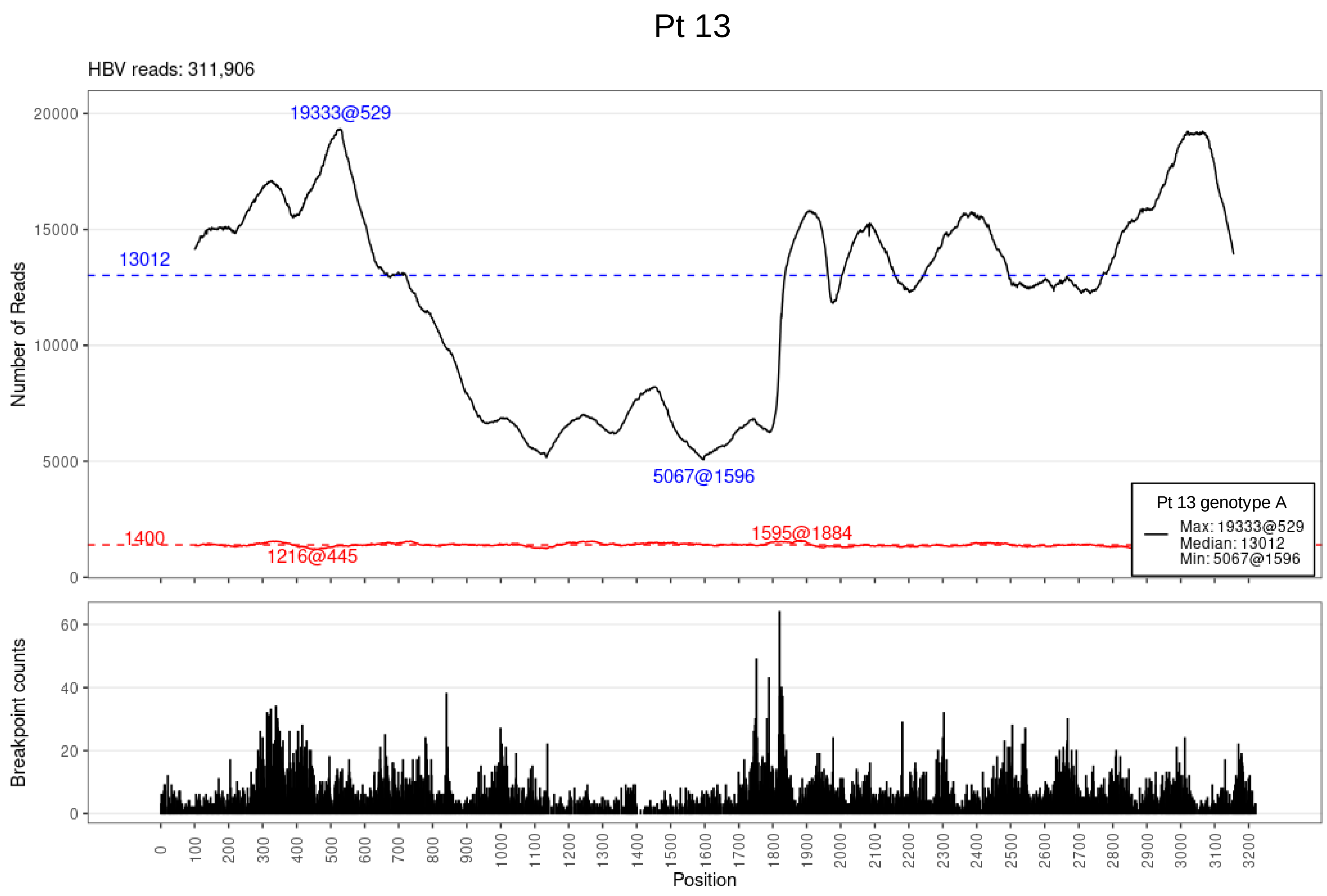

Pt 13
HBV reads: 311,906
Number of Reads
19333@529
13012
5067@1596
1400
1216@445
1595@1884
Pt 13 genotype A
Max: 19333@529
Median: 13012
Min: 5067@1596
Breakpoint counts
Position

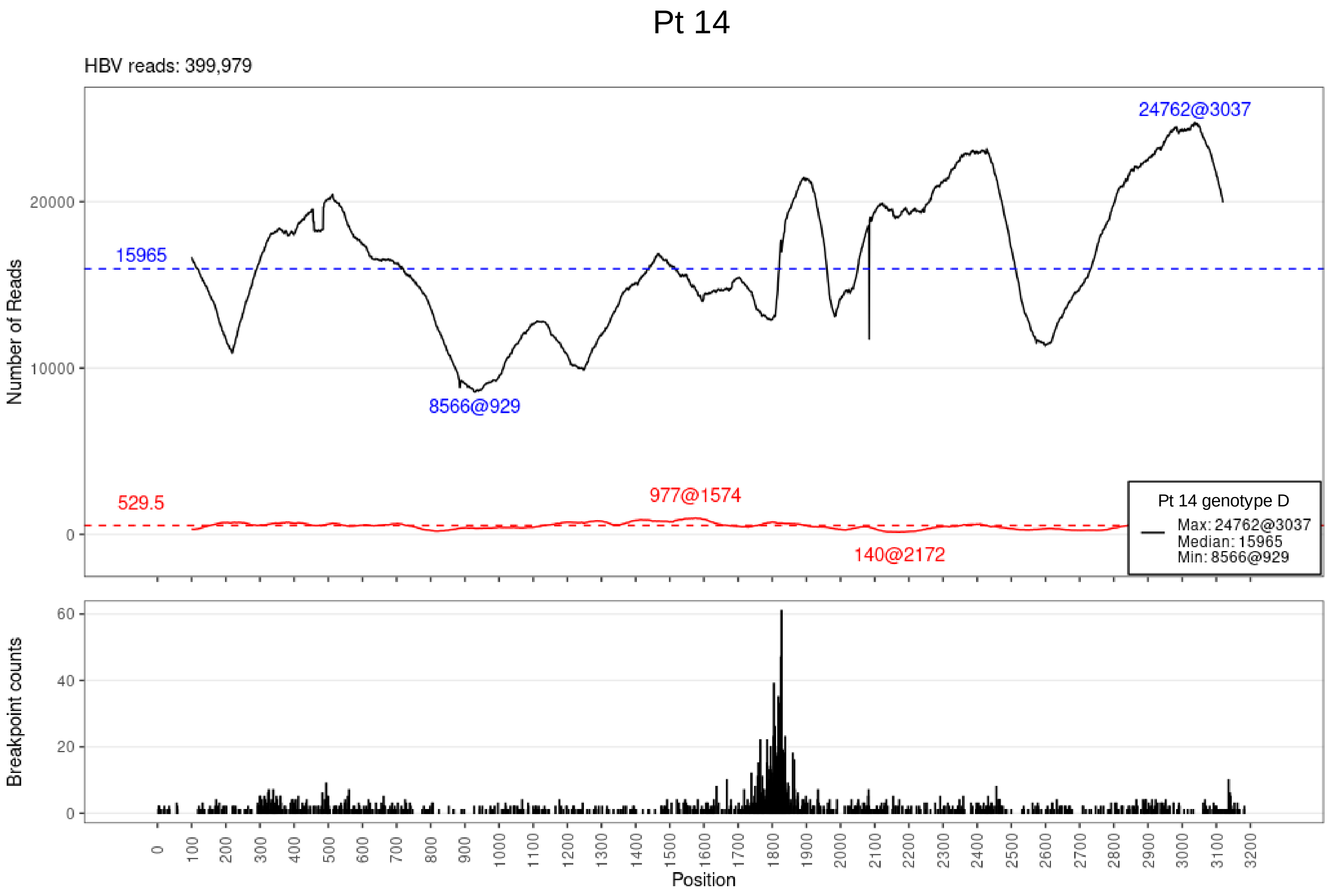

Pt 14
HBV reads: 399,979
24762@3037
15965
8566@929
529.5
977@1574
140@2172
Pt 14 genotype D
Max: 24762@3037
Median: 15965
Min: 8566@929
Number of Reads
Breakpoint counts
Position

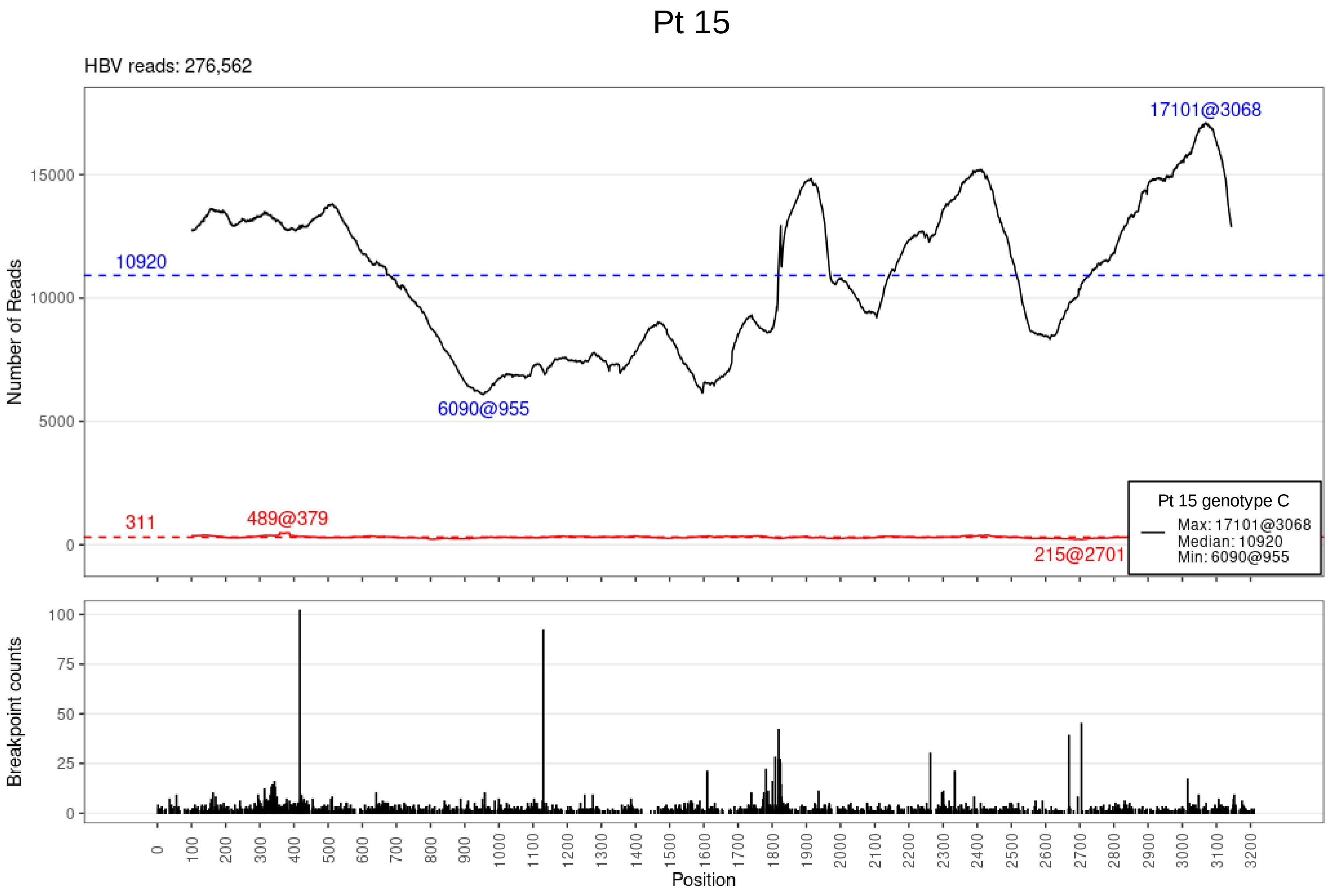

Pt 15
HBV reads: 276,562
Number of Reads
15000
10000
5000
0
17101@3068
10920
6090@955
311
489@379
215@2701
Pt 15 genotype C
Max: 17101@3068
Median: 10920
Min: 6090@955
Breakpoint counts
100
75
50
25
0
0 100 200 300 400 500 600 700 800 900 1000 1100 1200 1300 1400 1500 1600 1700 1800 1900 2000 2100 2200 2300 2400 2500 2600 2700 2800 2900 3000 3100 3200
Position

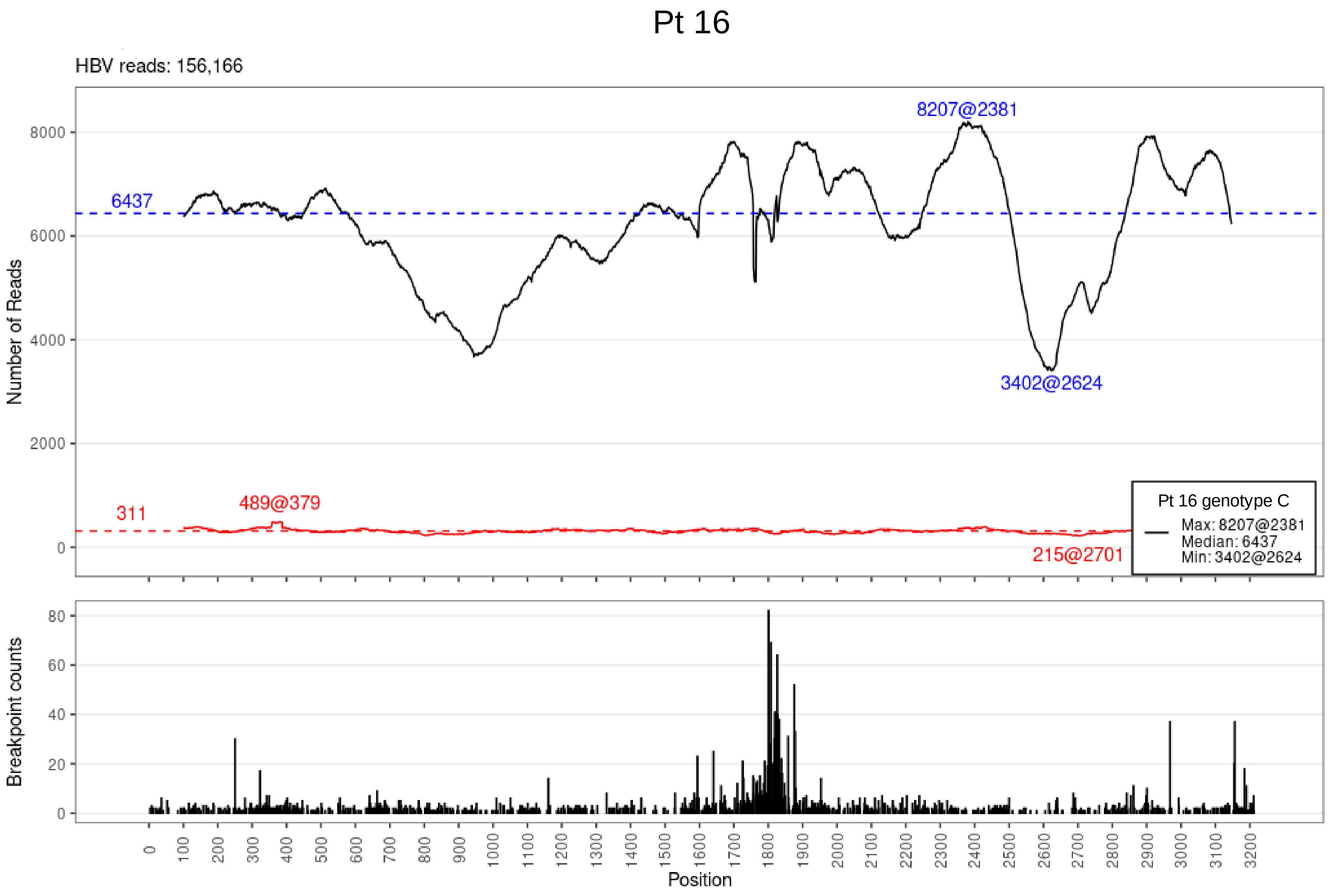

Pt 16
HBV reads: 156,166
Number of Reads
8000
6000
4000
2000
0
6437
8207@2381
3402@2624
311
489@379
215@2701
Pt 16 genotype C
Max: 8207@2381
Median: 6437
Min: 3402@2624
Breakpoint counts
80
60
40
20
0
0
100
200
300
400
500
600
700
800
900
1000
1100
1200
1300
1400
1500
1600
1700
1800
1900
2000
2100
2200
2300
2400
2500
2600
2700
2800
2900
3000
3100
3200
Position

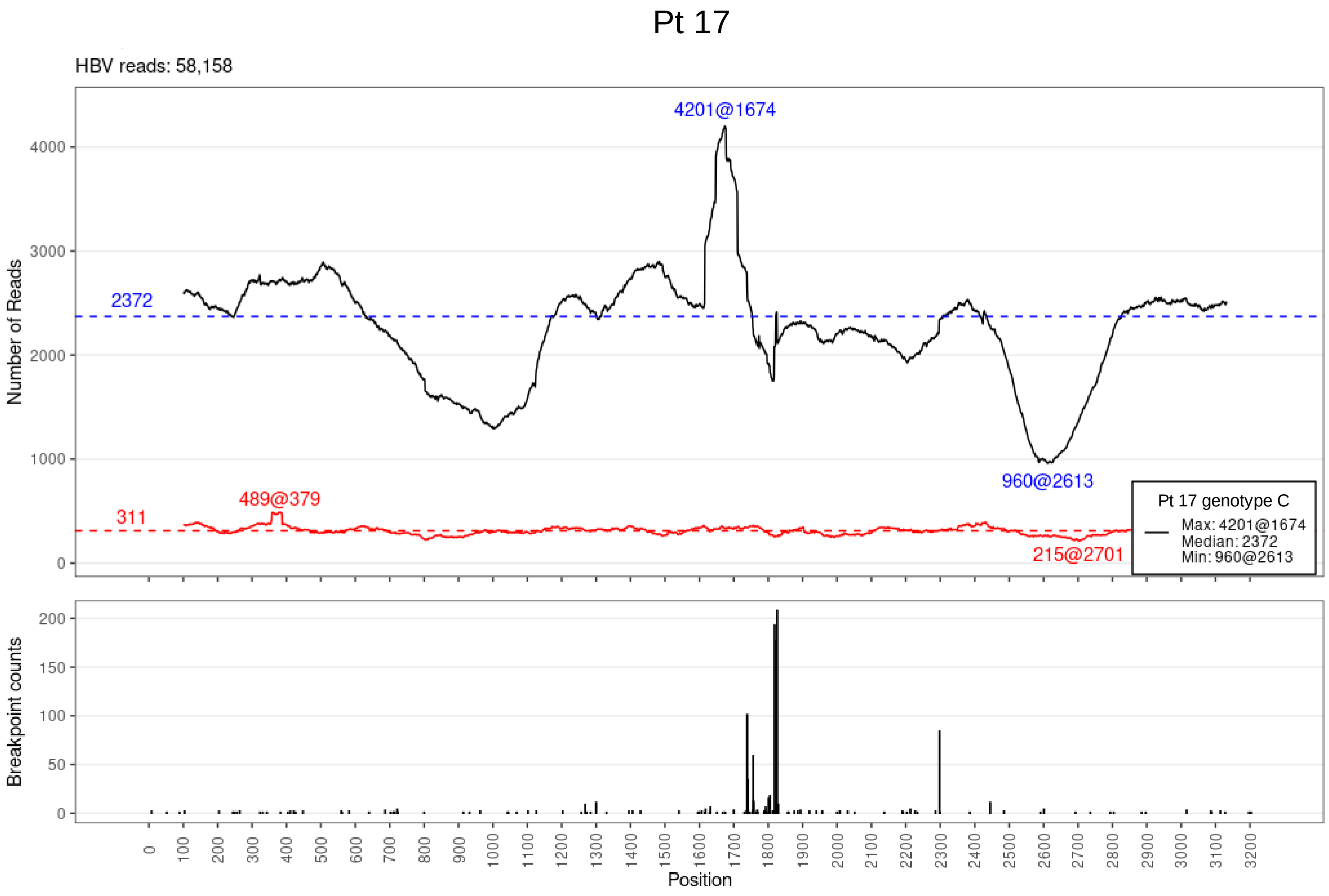

Pt 17
HBV reads: 58,158
4201@1674
2372
960@2613
489@379
311
215@2701
Pt 17 genotype C
Max: 4201@1674
Median: 2372
Min: 960@2613
Number of Reads
4000
3000
2000
1000
0
Breakpoint counts
200
150
100
50
0
0
100
200
300
400
500
600
700
800
900
1000
1100
1200
1300
1400
1500
1600
1700
1800
1900
2000
2100
2200
2300
2400
2500
2600
2700
2800
2900
3000
3100
3200
Position

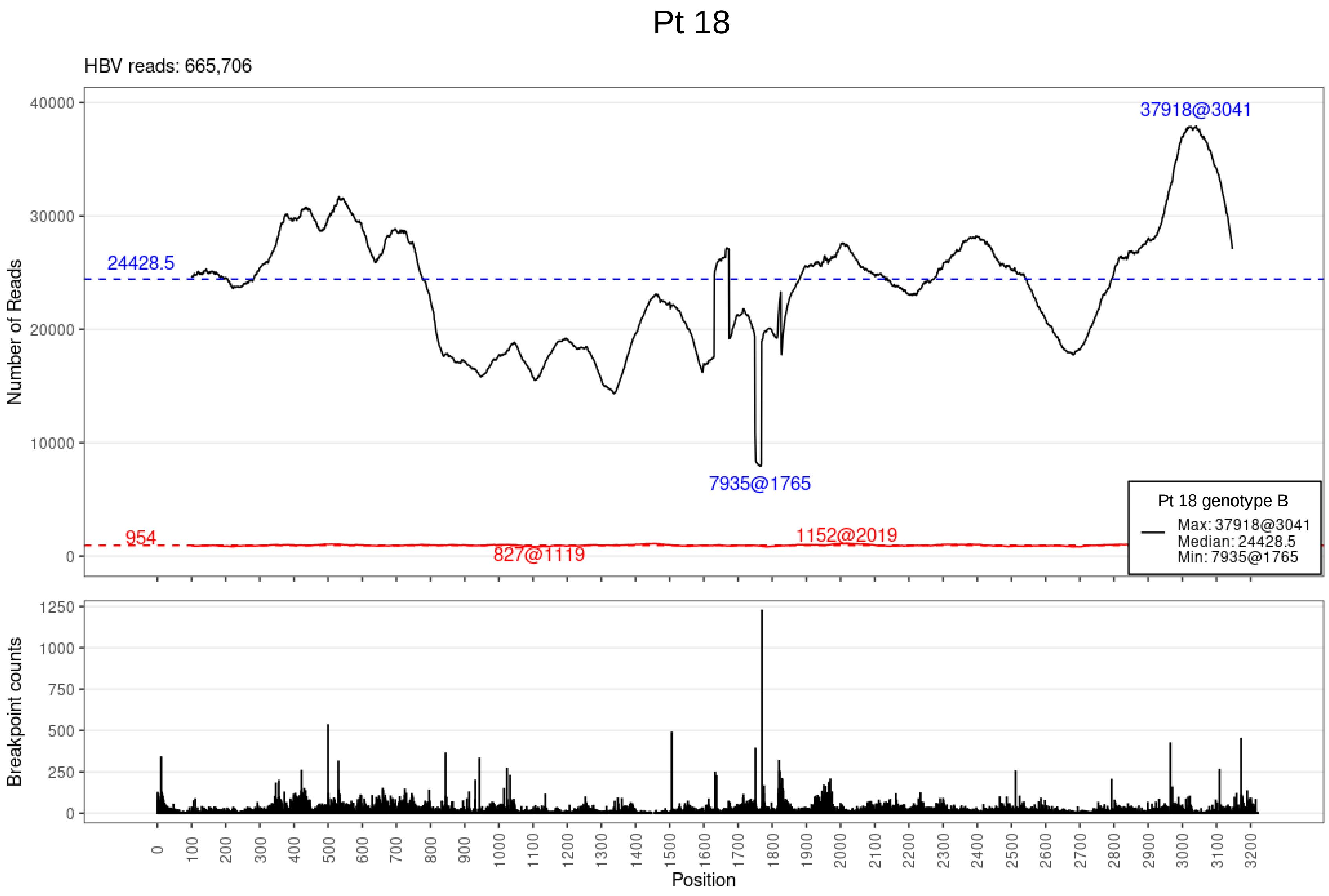
Pt 18
HBV reads: 665,706
Number of Reads
37918@3041
24428.5
7935@1765
954
827@1119
1152@2019
Pt 18 genotype B
Max: 37918@3041
Median: 24428.5
Min: 7935@1765
Breakpoint counts
Position

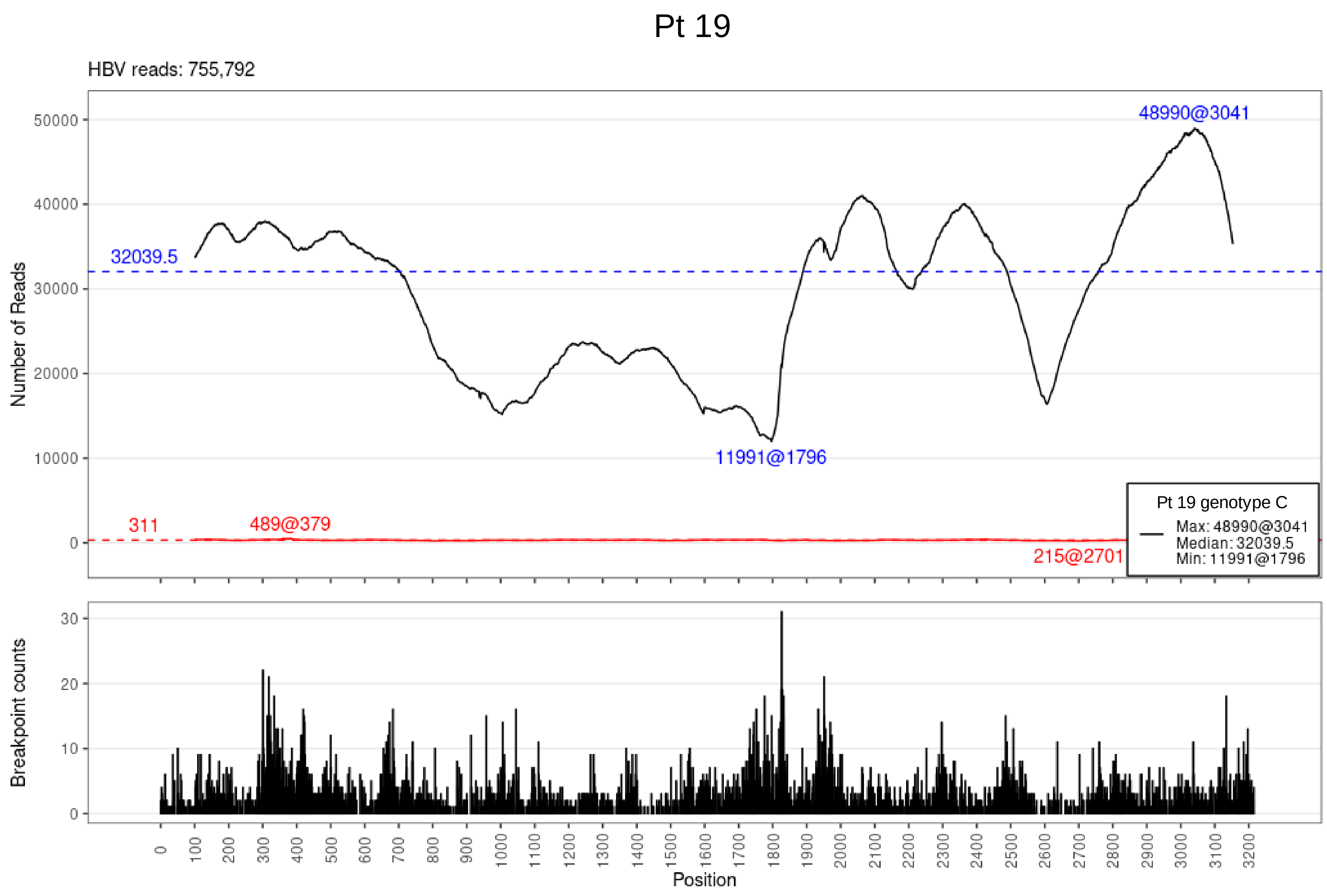

Pt 19
HBV reads: 755,792
48990@3041
32039.5
11991@1796
311
489@379
215@2701
Pt 19 genotype C
Max: 48990@3041
Median: 32039.5
Min: 11991@1796
Number of Reads
Breakpoint counts
Position

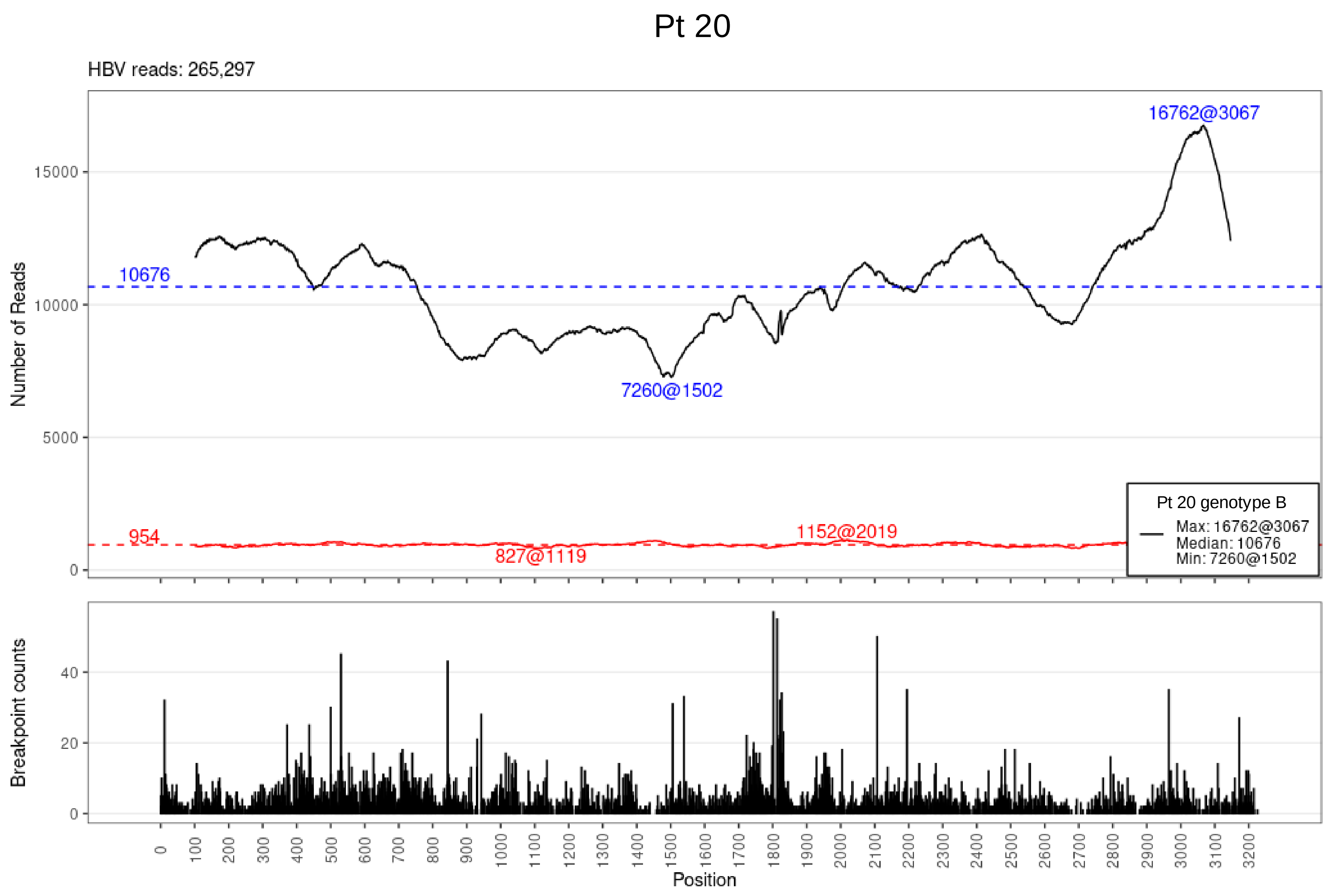

Pt 20
HBV reads: 265,297
Number of Reads
16762@3067
10676
7260@1502
954
1152@2019
827@1119
Pt 20 genotype B
Max: 16762@3067
Median: 10676
Min: 7260@1502
Breakpoint counts
Position

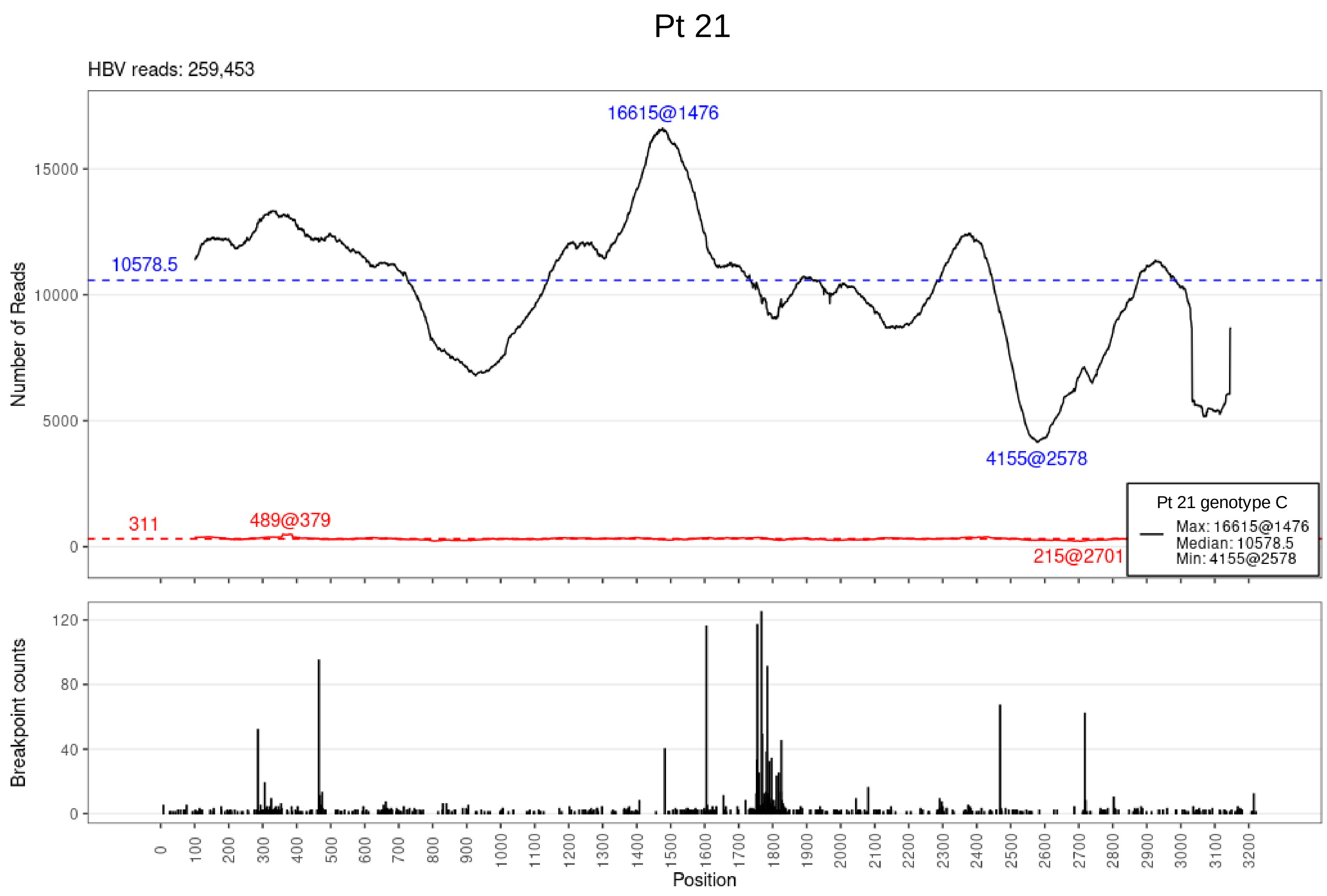

Pt 21
HBV reads: 259,453
Number of Reads
15000
10000
5000
0
16615@1476
10578.5
4155@2578
311
489@379
215@2701
Pt 21 genotype C
Max: 16615@1476
Median: 10578.5
Min: 4155@2578
Breakpoint counts
120
80
40
0
0
100
200
300
400
500
600
700
800
900
1000
1100
1200
1300
1400
1500
1600
1700
1800
1900
2000
2100
2200
2300
2400
2500
2600
2700
2800
2900
3000
3100
3200
Position

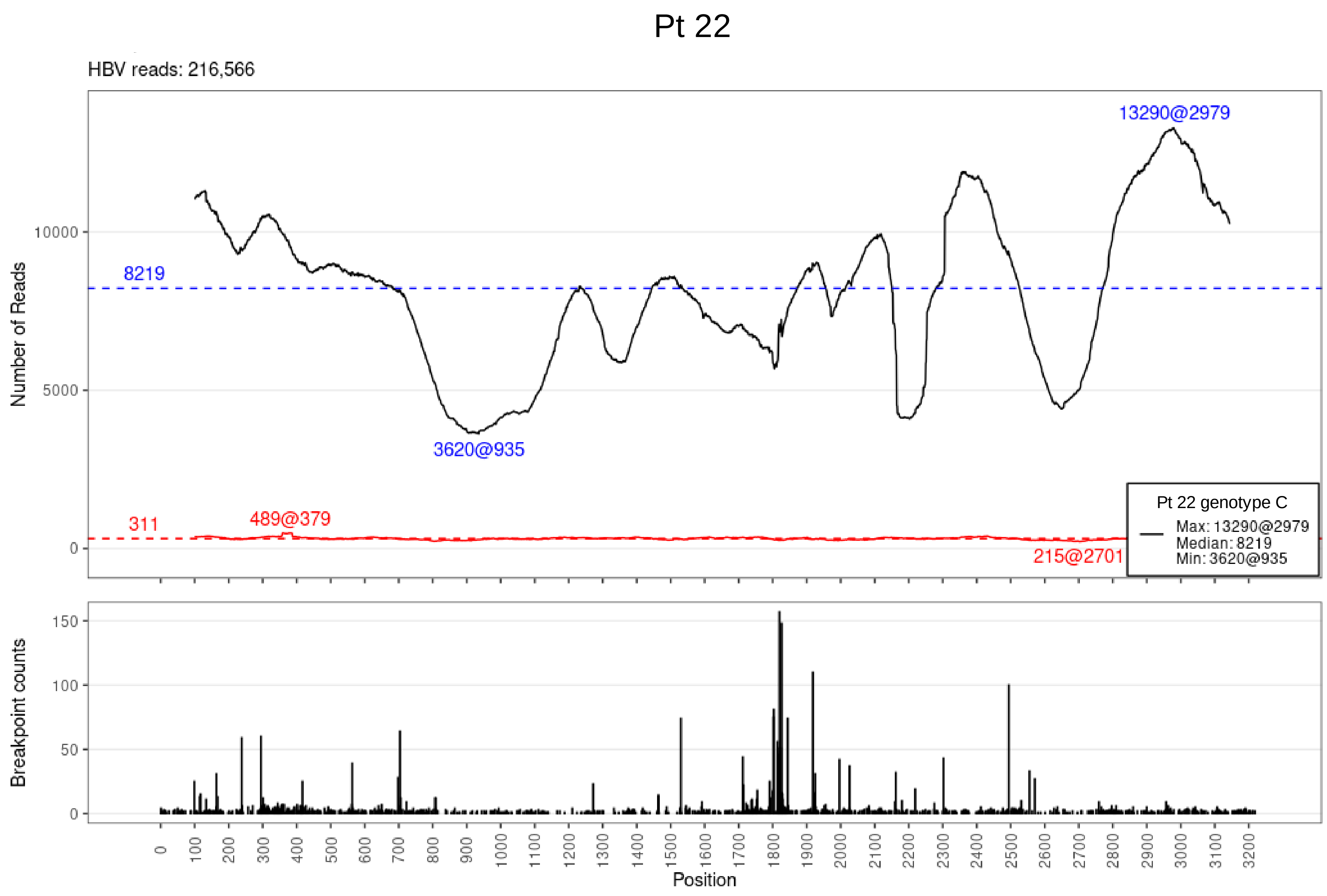

Pt 22
HBV reads: 216,566
Number of Reads
13290@2979
8219
3620@935
311
489@379
215@2701
Pt 22 genotype C
Max: 13290@2979
Median: 8219
Min: 3620@935
Breakpoint counts
Position

# Pt 23

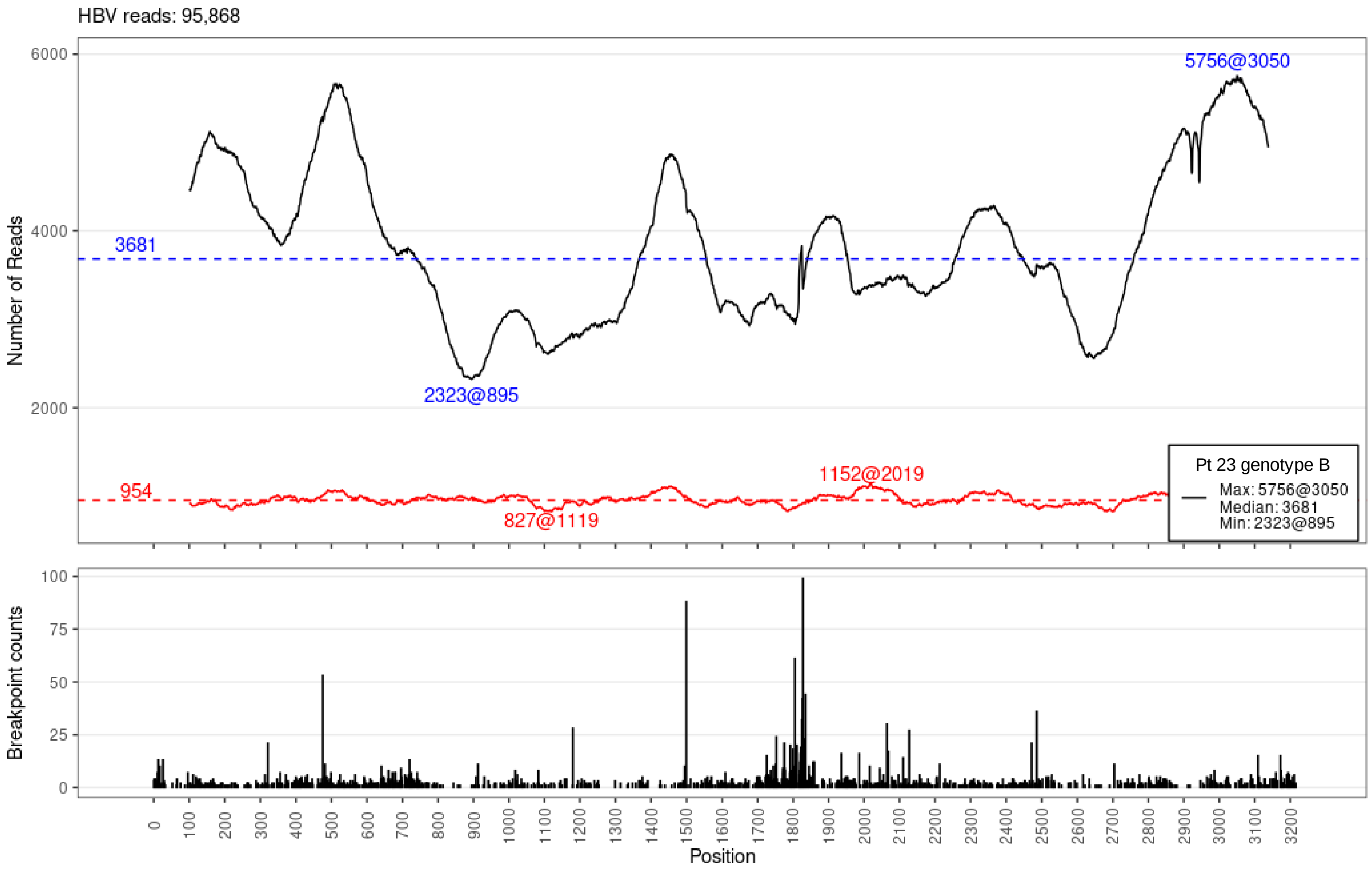

HBV reads: 95,868
Number of Reads
6000
4000
2000
3681
5756@3050
2323@895
954
1152@2019
827@1119
Pt 23 genotype B
Max: 5756@3050
Median: 3681
Min: 2323@895
Breakpoint counts
100
75
50
25
0
0
100
200
300
400
500
600
700
800
900
1000
1100
1200
1300
1400
1500
1600
1700
1800
1900
2000
2100
2200
2300
2400
2500
2600
2700
2800
2900
3000
3100
3200
Position

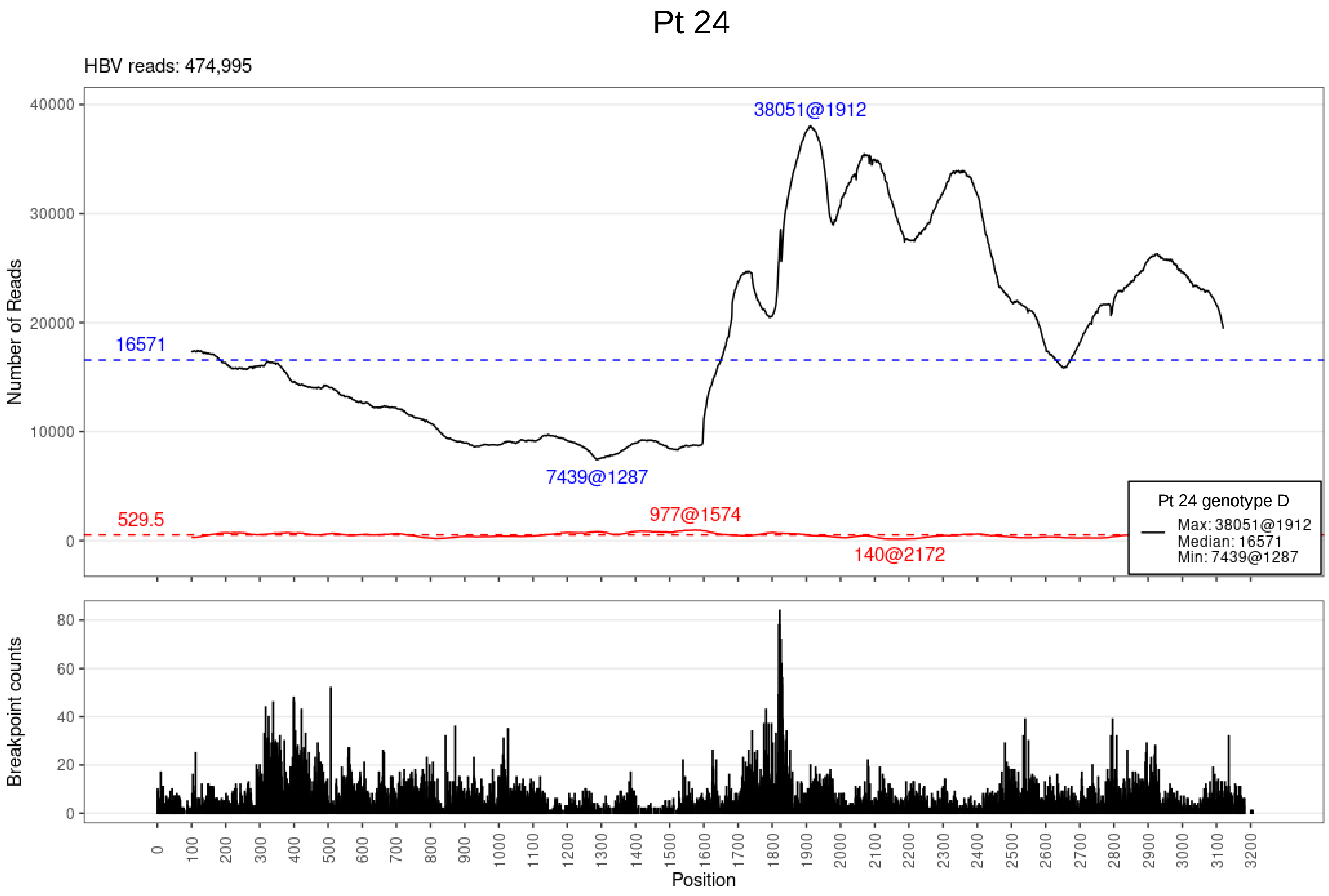

Pt 24
HBV reads: 474,995
Number of Reads
40000
30000
20000
10000
0
38051@1912
16571
7439@1287
529.5
977@1574
140@2172
Pt 24 genotype D
Max: 38051@1912
Median: 16571
Min: 7439@1287
Breakpoint counts
80
60
40
20
0
0 100 200 300 400 500 600 700 800 900 1000 1100 1200 1300 1400 1500 1600 1700 1800 1900 2000 2100 2200 2300 2400 2500 2600 2700 2800 2900 3000 3100 3200
Position

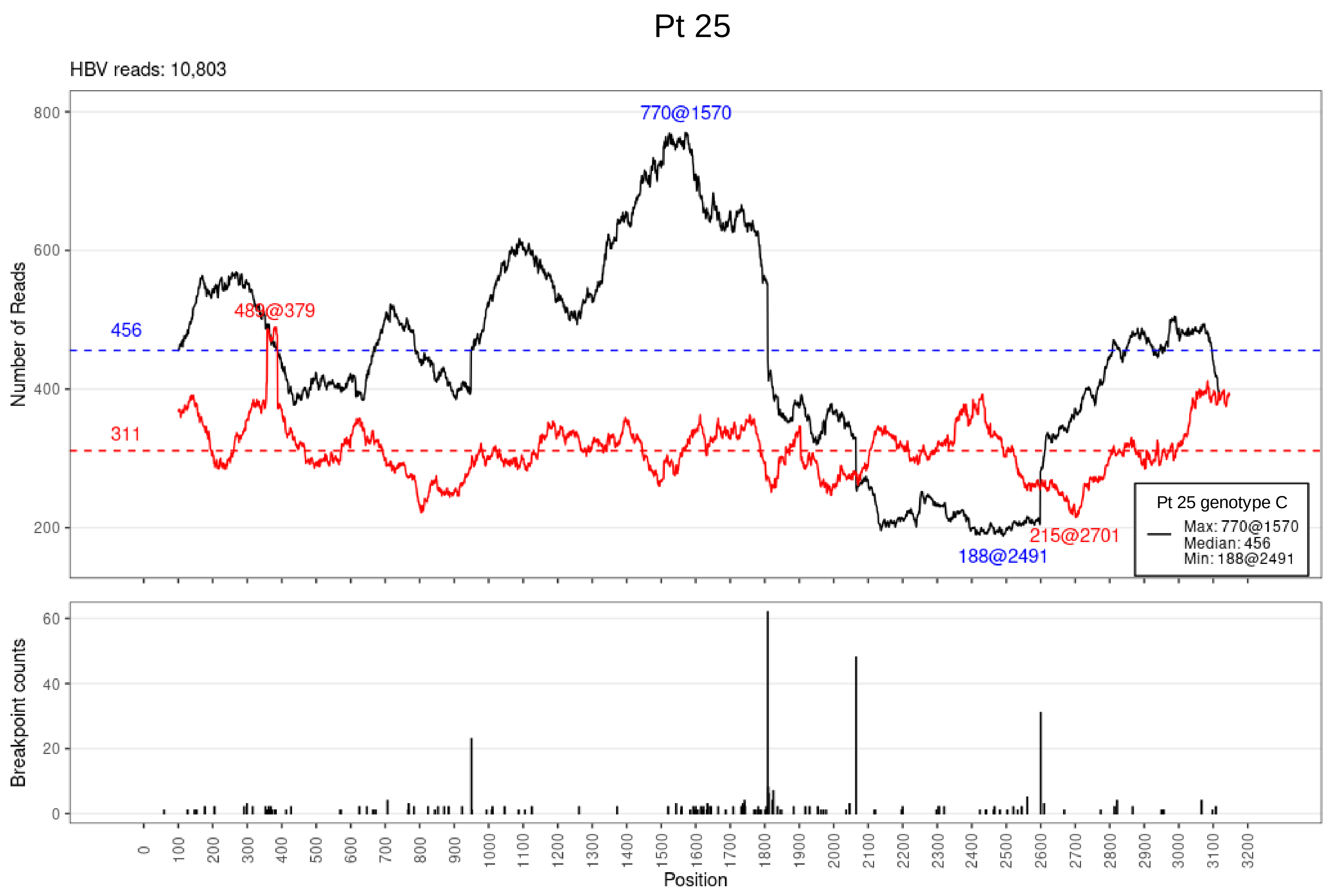
Pt 25
HBV reads: 10,803
Number of Reads
770@1570
489@379
456
311
215@2701
188@2491
Pt 25 genotype C
Max: 770@1570
Median: 456
Min: 188@2491
Breakpoint counts
Position

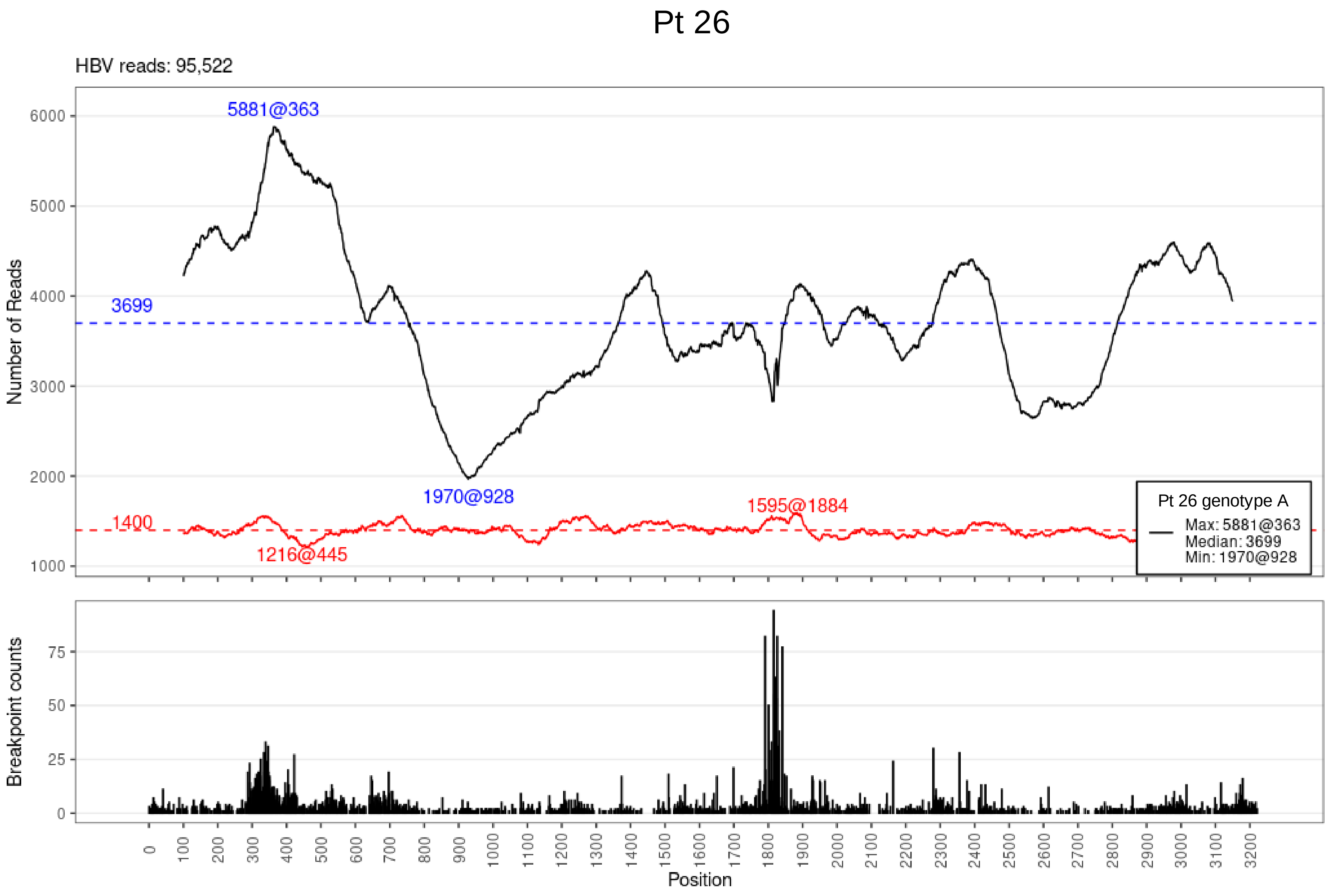

Pt 26
HBV reads: 95,522
Number of Reads
6000
5000
4000
3000
2000
1000
5881@363
3699
1970@928
1400
1216@445
1595@1884
Pt 26 genotype A
Max: 5881@363
Median: 3699
Min: 1970@928
Breakpoint counts
75
50
25
0
0 100 200 300 400 500 600 700 800 900 1000 1100 1200 1300 1400 1500 1600 1700 1800 1900 2000 2100 2200 2300 2400 2500 2600 2700 2800 2900 3000 3100 3200
Position

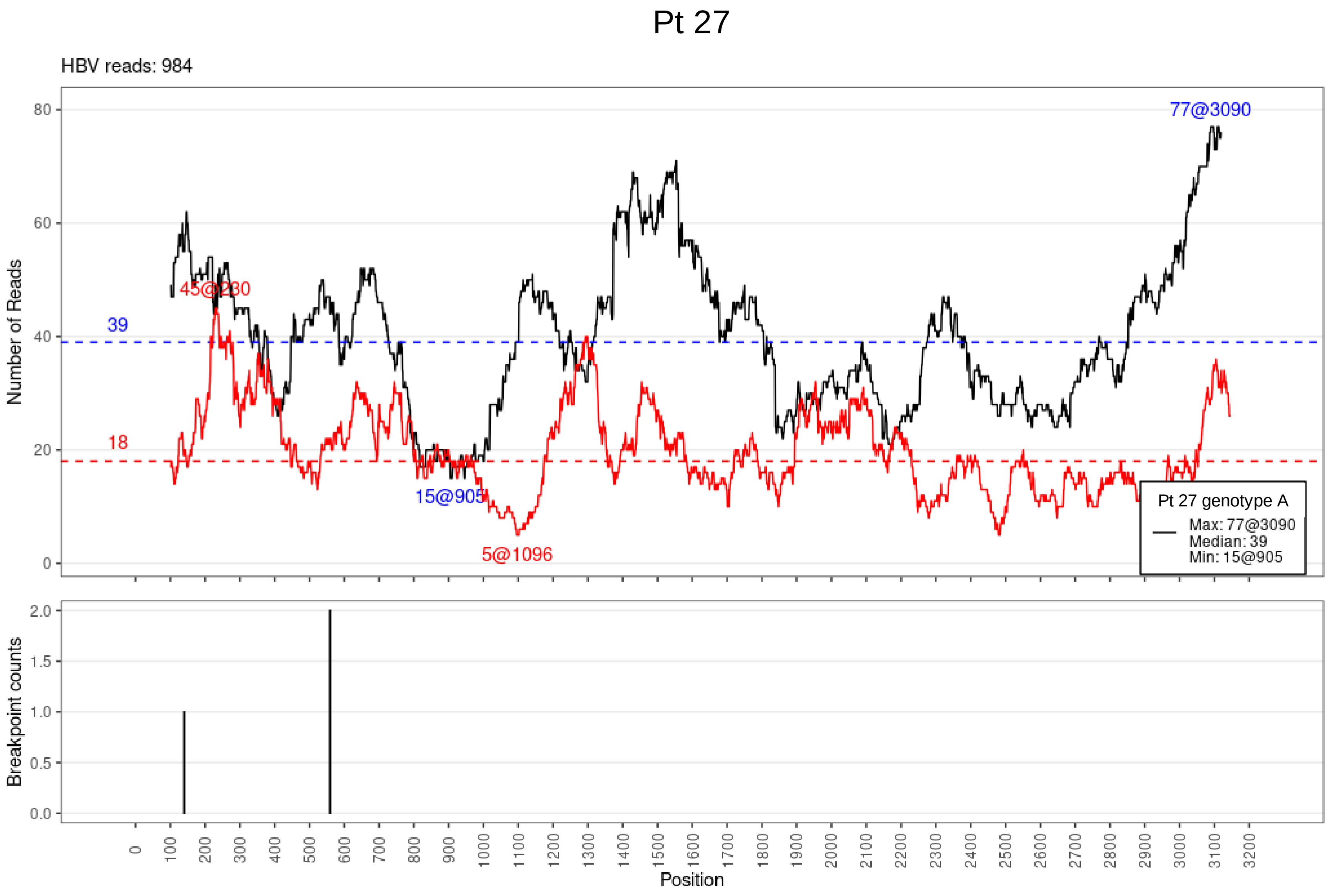

Pt 27
HBV reads: 984
Number of Reads
77@3090
45@230
39
18
15@905
5@1096
Pt 27 genotype A
Max: 77@3090
Median: 39
Min: 15@905
Breakpoint counts
Position

## Pt 28

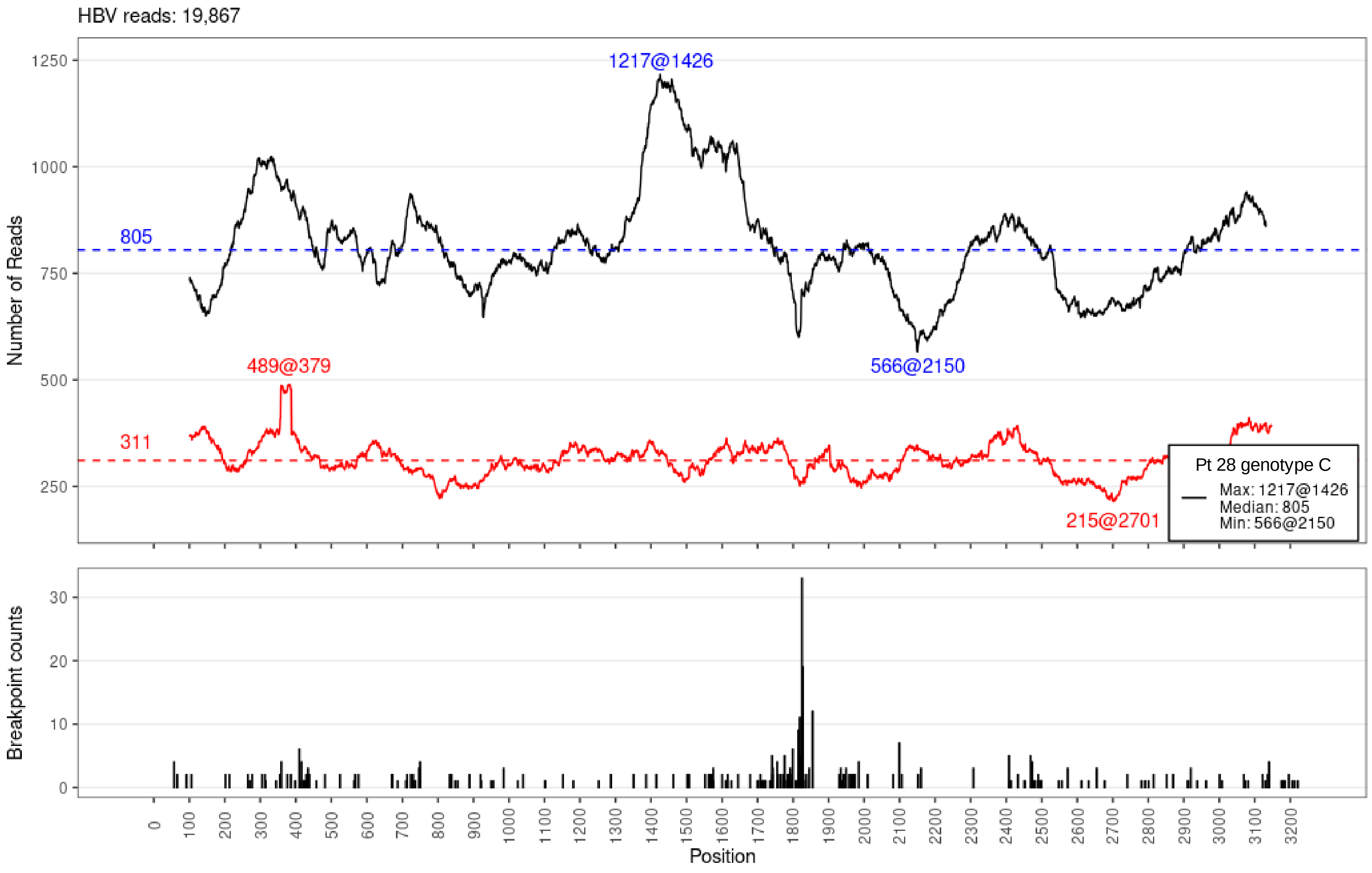
HBV reads: 19,867
Number of Reads
1250
1000
750
500
250
1217@1426
805
489@379
566@2150
311
215@2701
Pt 28 genotype C
Max: 1217@1426
Median: 805
Min: 566@2150
Breakpoint counts
30
20
10
0
0
100
200
300
400
500
600
700
800
900
1000
1100
1200
1300
1400
1500
1600
1700
1800
1900
2000
2100
2200
2300
2400
2500
2600
2700
2800
2900
3000
3100
3200
Position

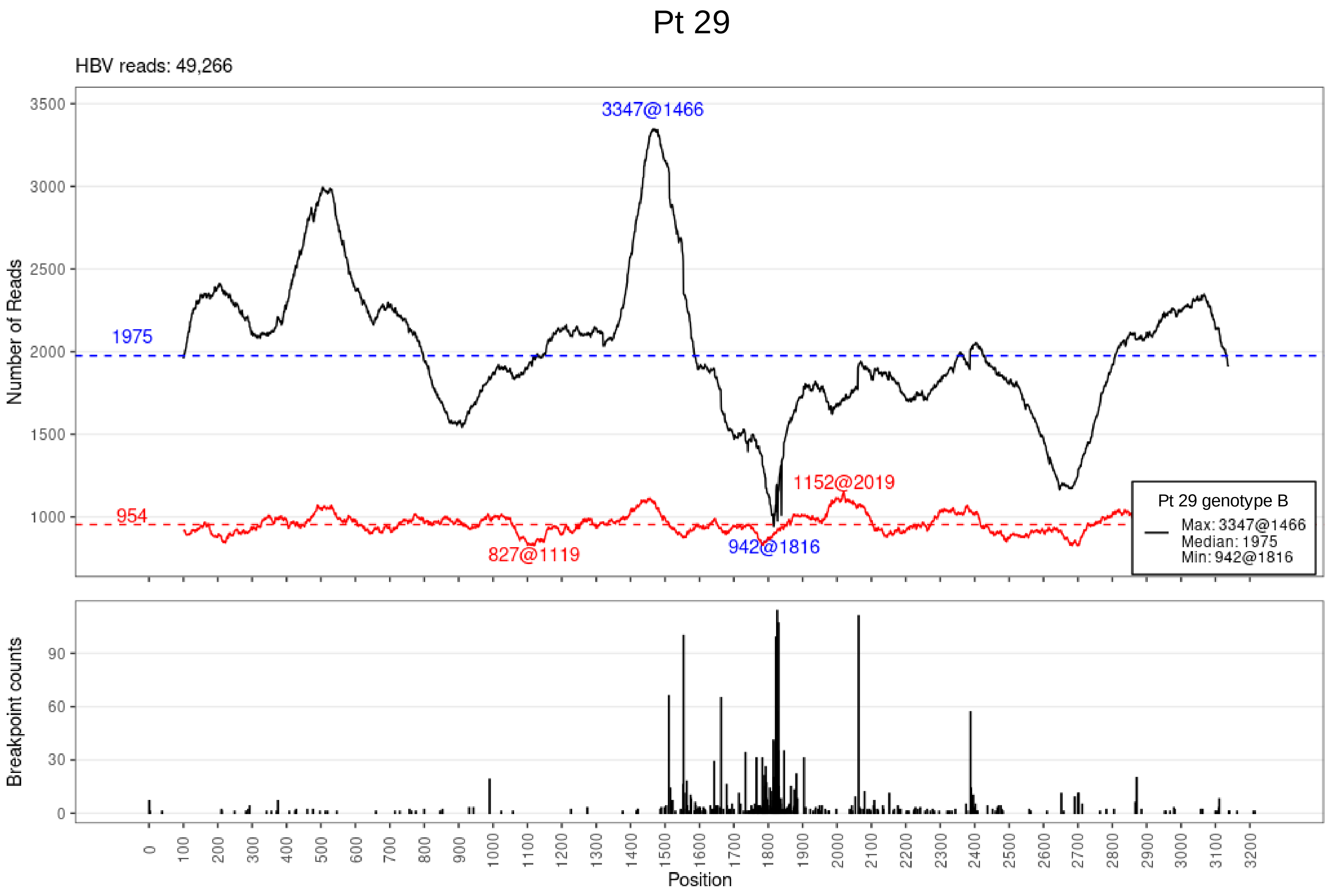

Pt 29
HBV reads: 49,266
3347@1466
1975
954
827@1119
942@1816
1152@2019
Pt 29 genotype B
Max: 3347@1466
Median: 1975
Min: 942@1816
Number of Reads
Breakpoint counts
Position

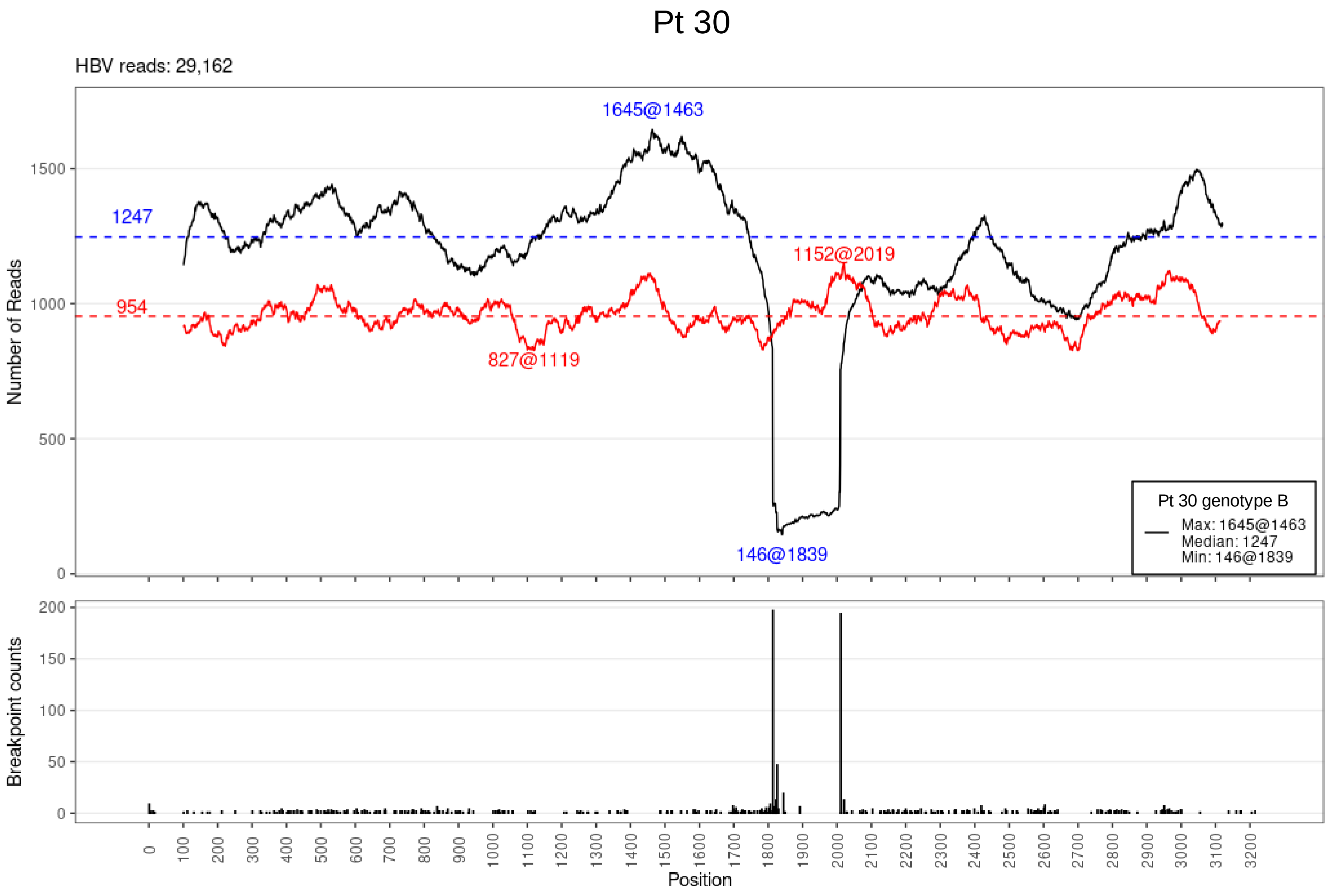

Pt 30
HBV reads: 29,162
1645@1463
1247
954
1152@2019
827@1119
146@1839
Pt 30 genotype B
Max: 1645@1463
Median: 1247
Min: 146@1839
Number of Reads
Breakpoint counts
Position

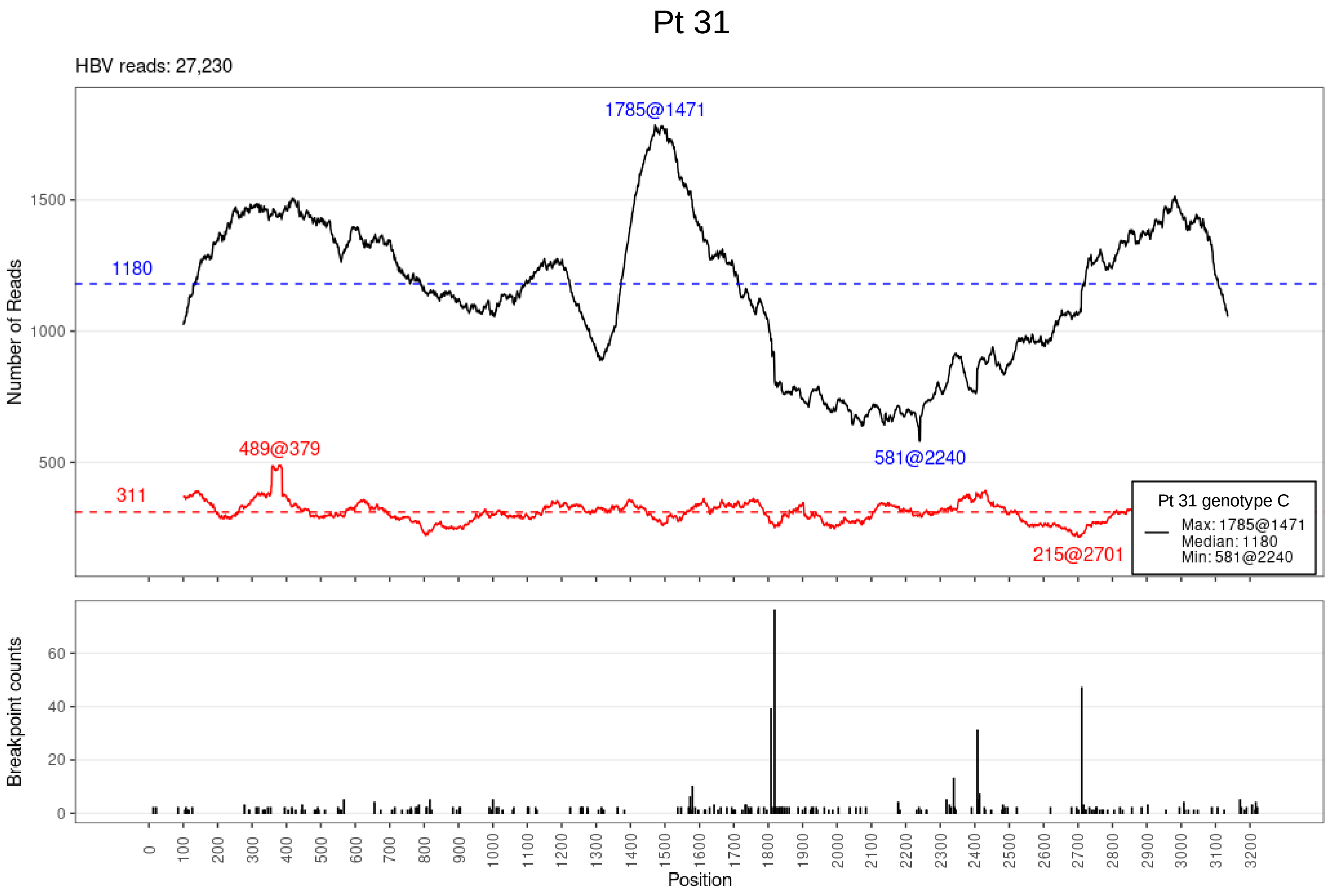
Pt 31
HBV reads: 27,230
Number of Reads
1785@1471
1180
489@379
311
581@2240
215@2701
Pt 31 genotype C
Max: 1785@1471
Median: 1180
Min: 581@2240
Breakpoint counts
Position

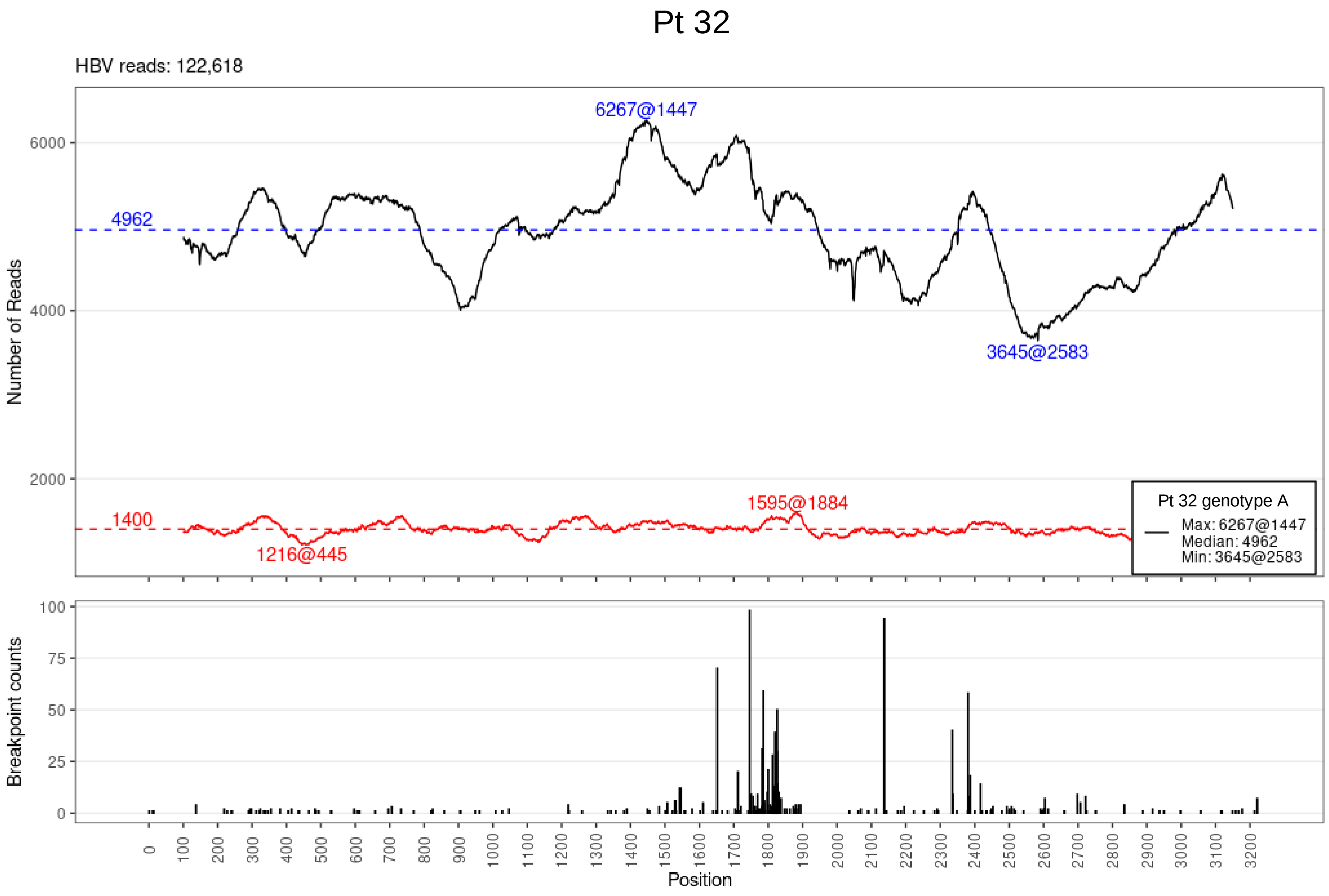
Pt 32
HBV reads: 122,618
Number of Reads
6000
4000
2000
6267@1447
4962
3645@2583
1595@1884
1400
1216@445
Pt 32 genotype A
Max: 6267@1447
Median: 4962
Min: 3645@2583
Breakpoint counts
100
75
50
25
0
Position

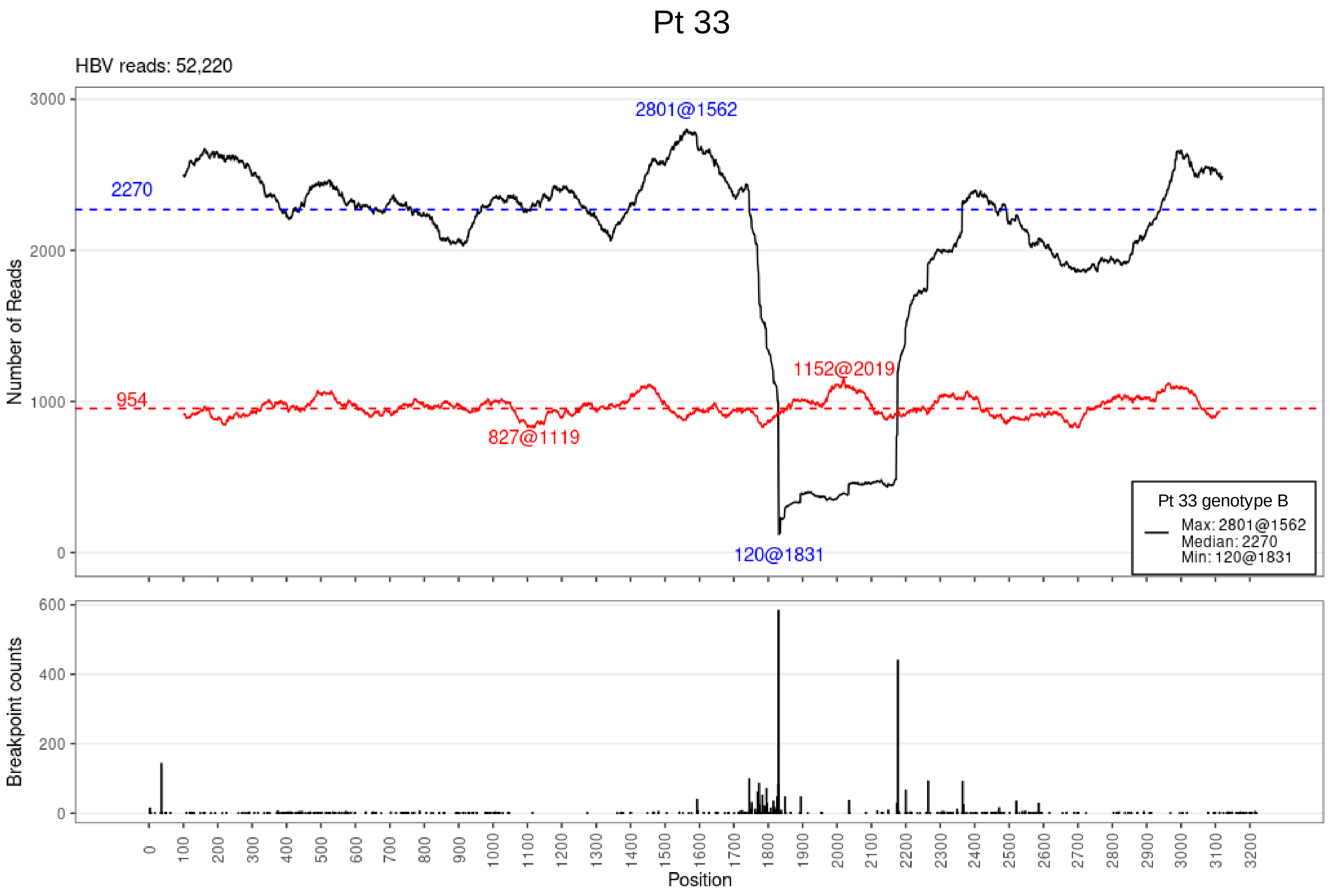

Pt 33
HBV reads: 52,220
Number of Reads
3000
2000
1000
0
2801@1562
2270
954
1152@2019
827@1119
120@1831
Pt 33 genotype B
Max: 2801@1562
Median: 2270
Min: 120@1831
Breakpoint counts
600
400
200
0
0
100
200
300
400
500
600
700
800
900
1000
1100
1200
1300
1400
1500
1600
1700
1800
1900
2000
2100
2200
2300
2400
2500
2600
2700
2800
2900
3000
3100
3200
Position

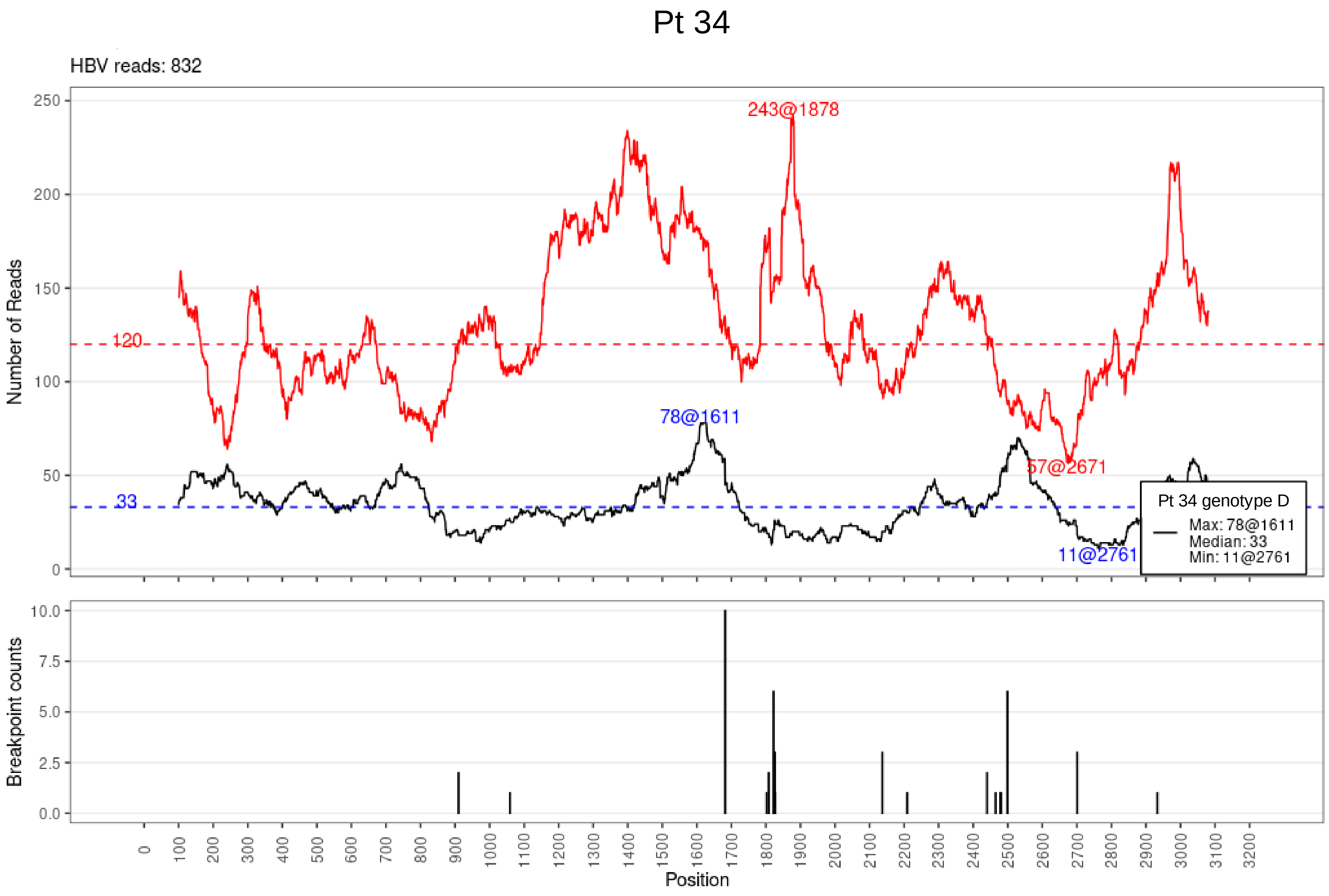
Pt 34
HBV reads: 832
Number of Reads
243@1878
78@1611
57@2671
11@2761
120
33
Pt 34 genotype D
Max: 78@1611
Median: 33
Min: 11@2761
Breakpoint counts
Position

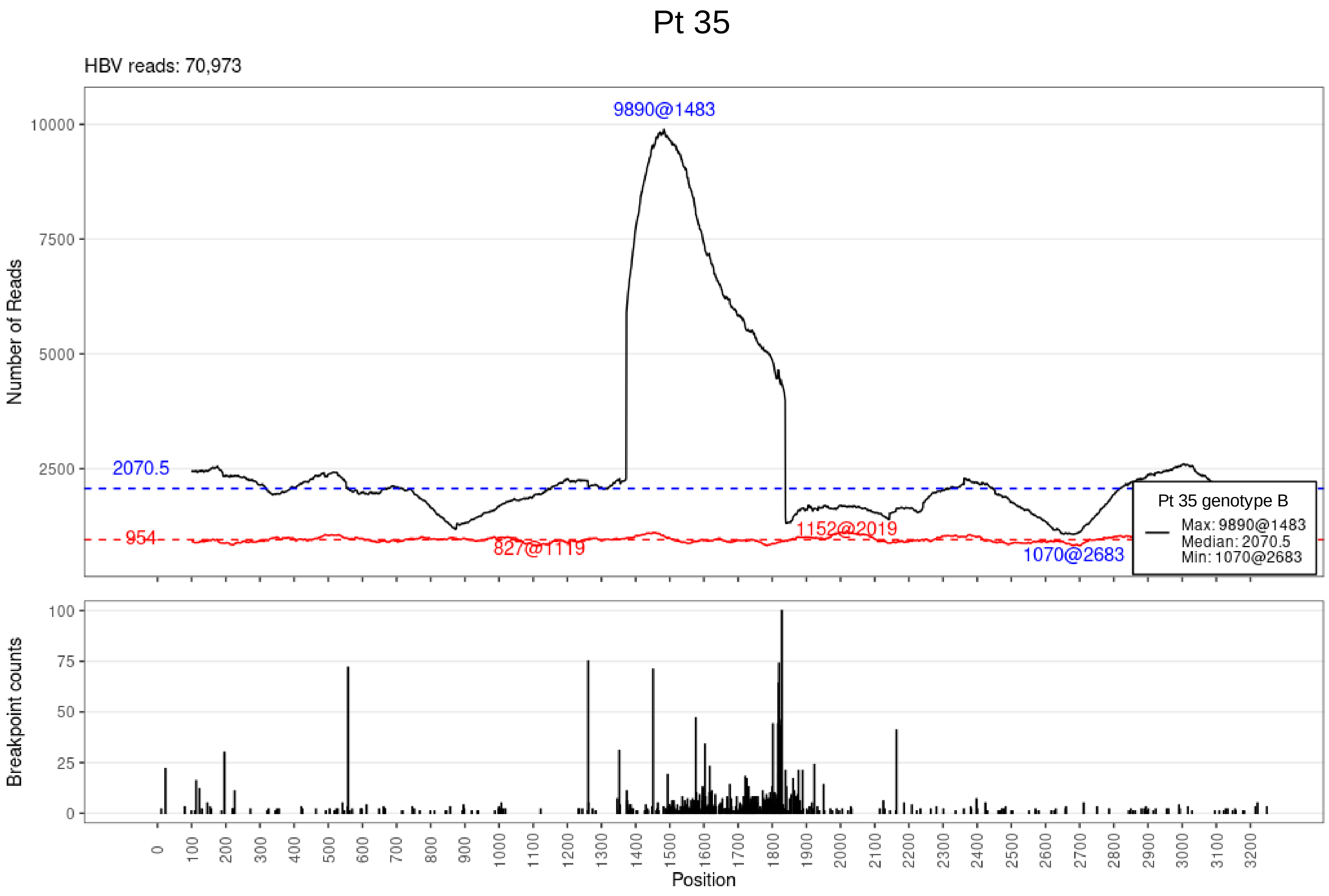
Pt 35
HBV reads: 70,973
9890@1483
2070.5
954
827@1119
1152@2019
1070@2683
Pt 35 genotype B
Max: 9890@1483
Median: 2070.5
Min: 1070@2683
Number of Reads
Breakpoint counts
Position

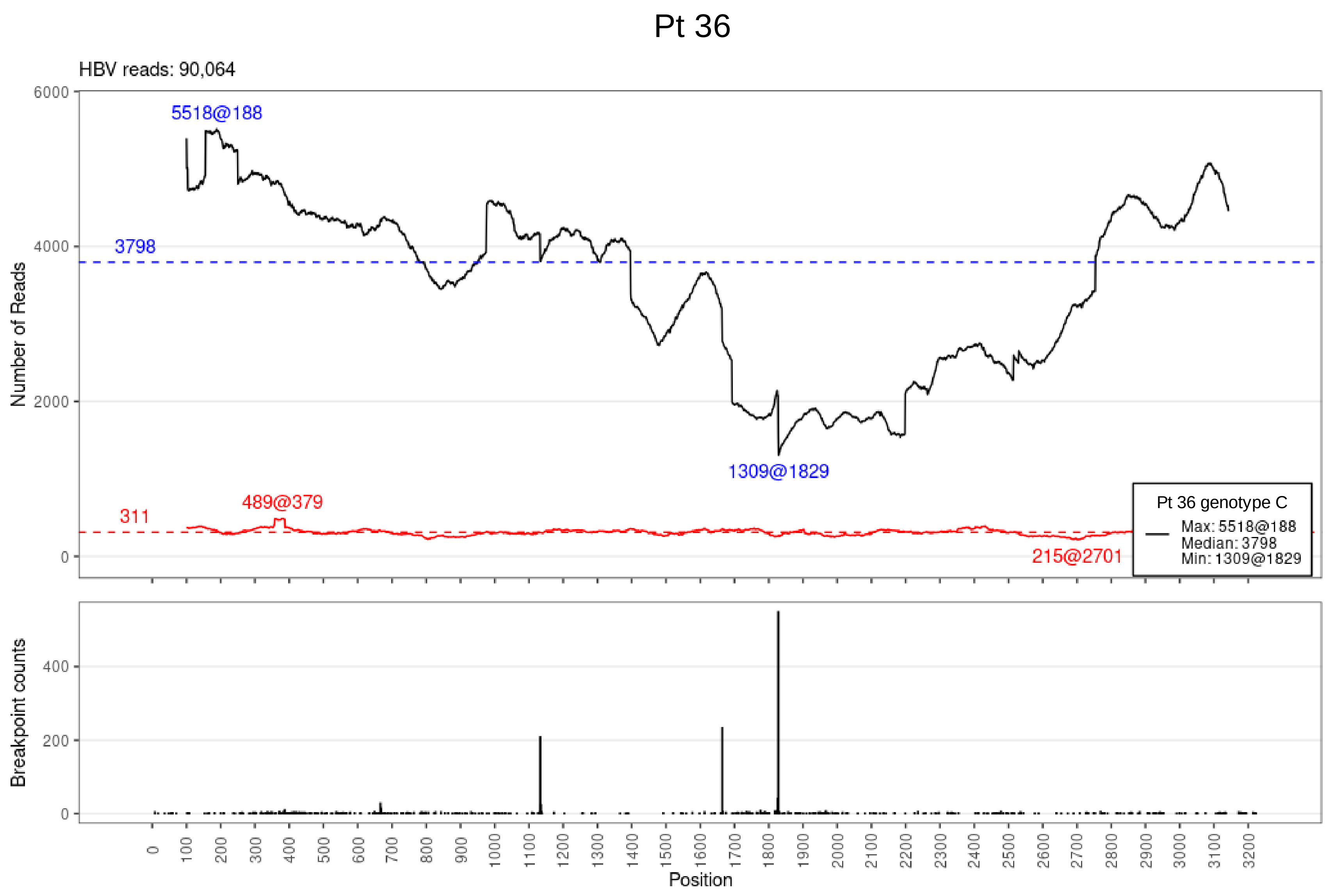

Pt 36
HBV reads: 90,064
5518@188
3798
1309@1829
489@379
311
215@2701
Pt 36 genotype C
Max: 5518@188
Median: 3798
Min: 1309@1829
Number of Reads
Breakpoint counts
Position

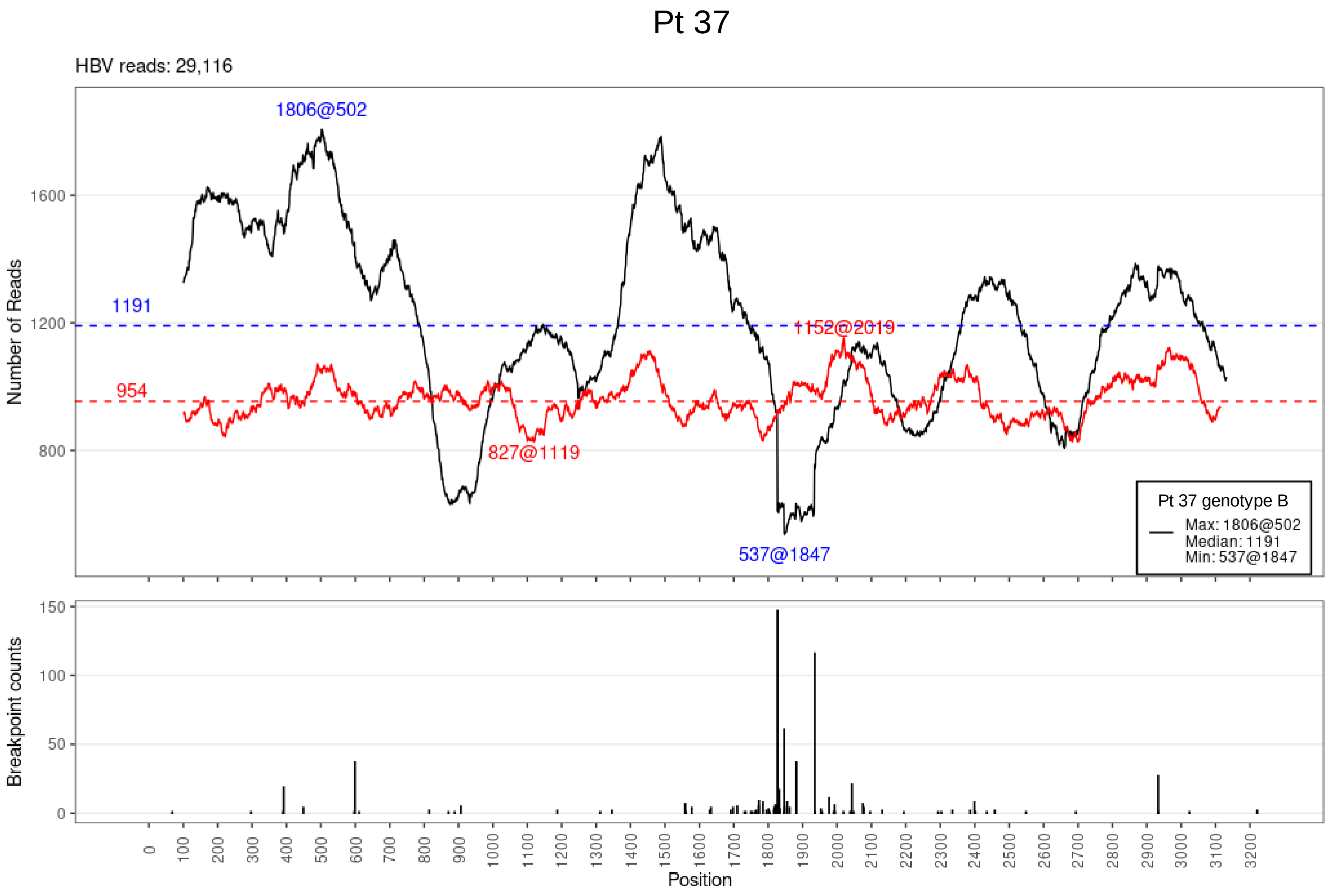
Pt 37
HBV reads: 29,116
1806@502
1191
954
1152@2019
827@1119
537@1847
Pt 37 genotype B
Max: 1806@502
Median: 1191
Min: 537@1847
Number of Reads
Breakpoint counts
Position

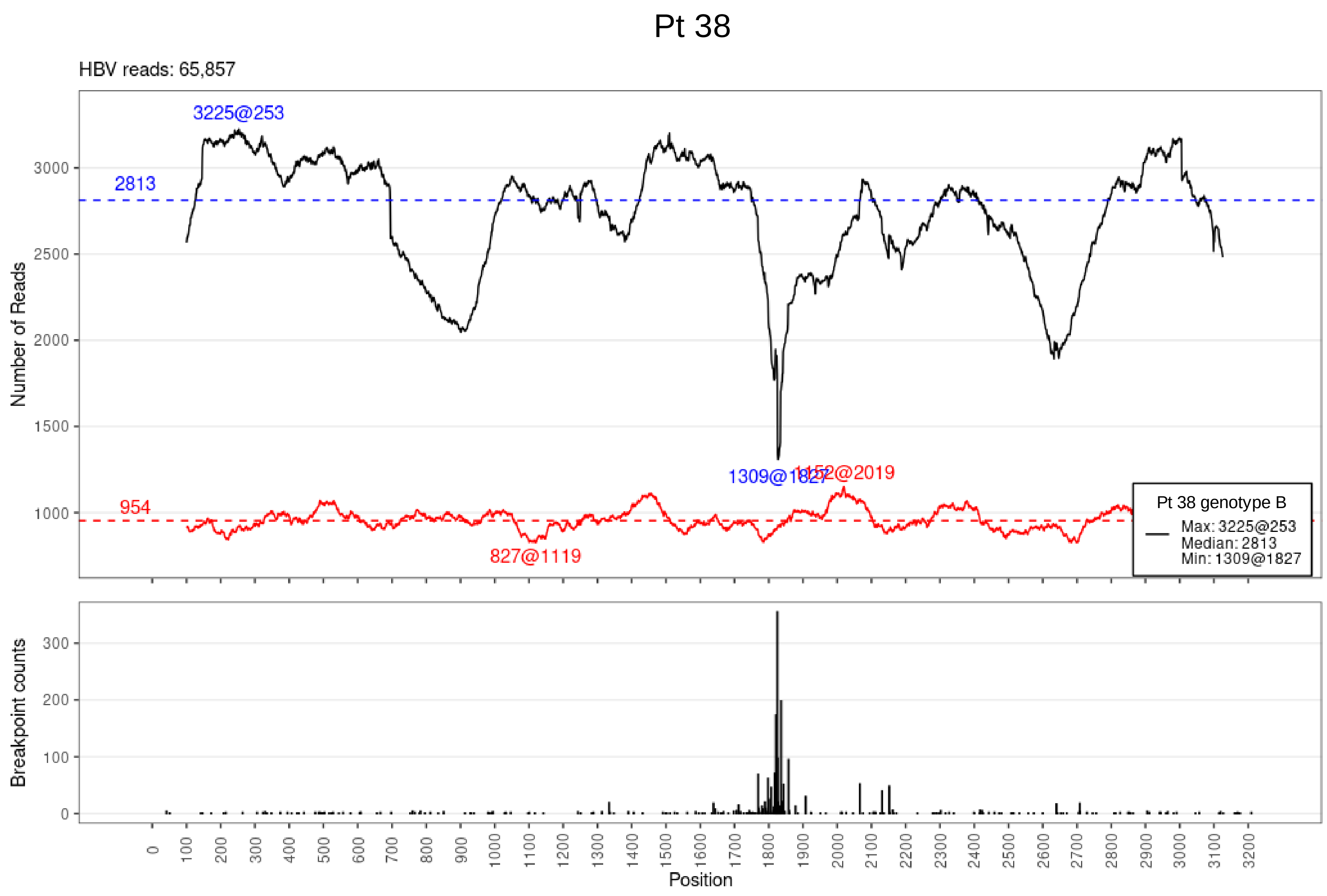

Pt 38
HBV reads: 65,857
3225@253
2813
954
827@1119
1309@1827
1152@2019
Pt 38 genotype B
Max: 3225@253
Median: 2813
Min: 1309@1827
Number of Reads
Breakpoint counts
Position

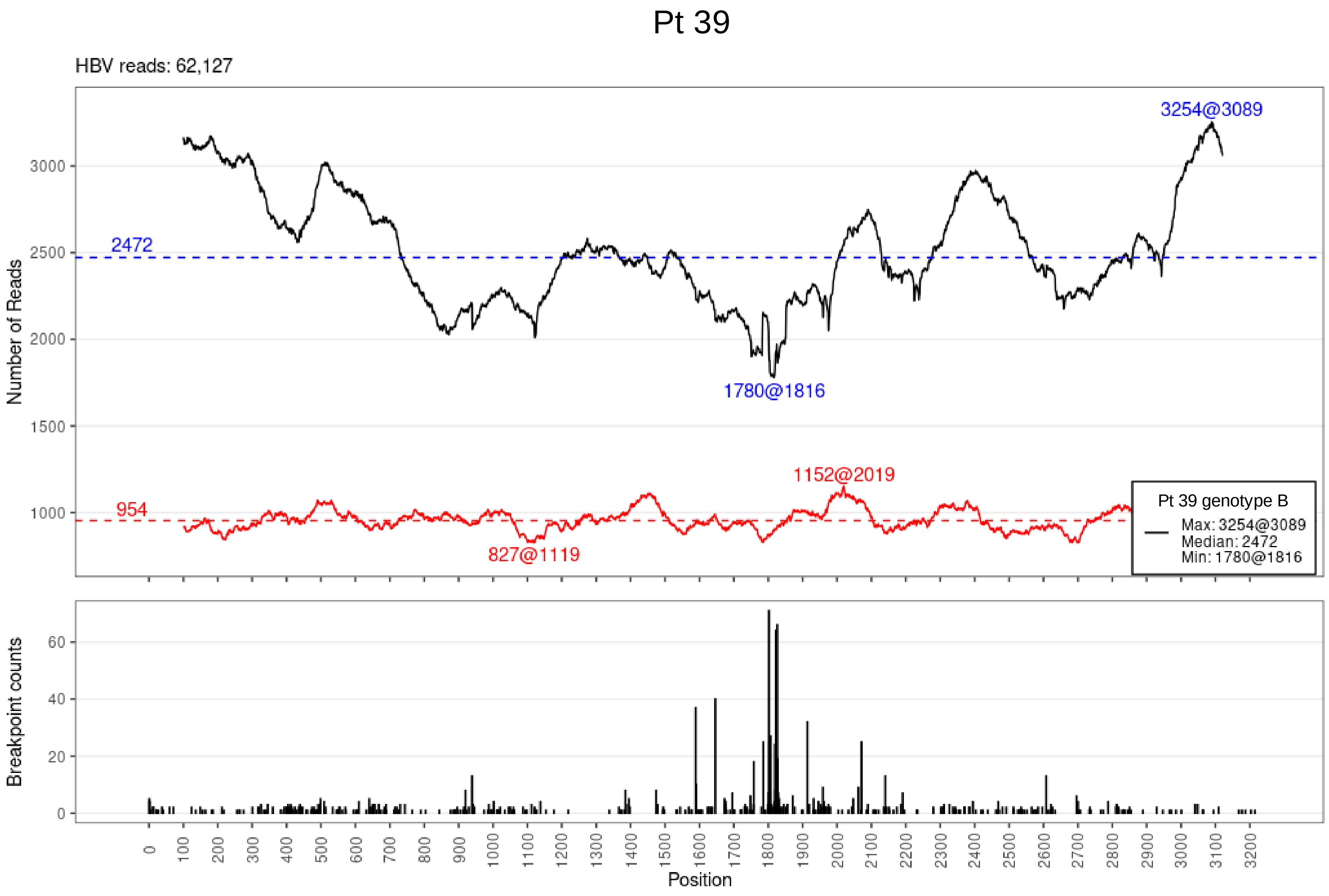
Pt 39
HBV reads: 62,127
3254@3089
2472
1780@1816
954
1152@2019
827@1119
Pt 39 genotype B
Max: 3254@3089
Median: 2472
Min: 1780@1816
Number of Reads
Breakpoint counts
Position

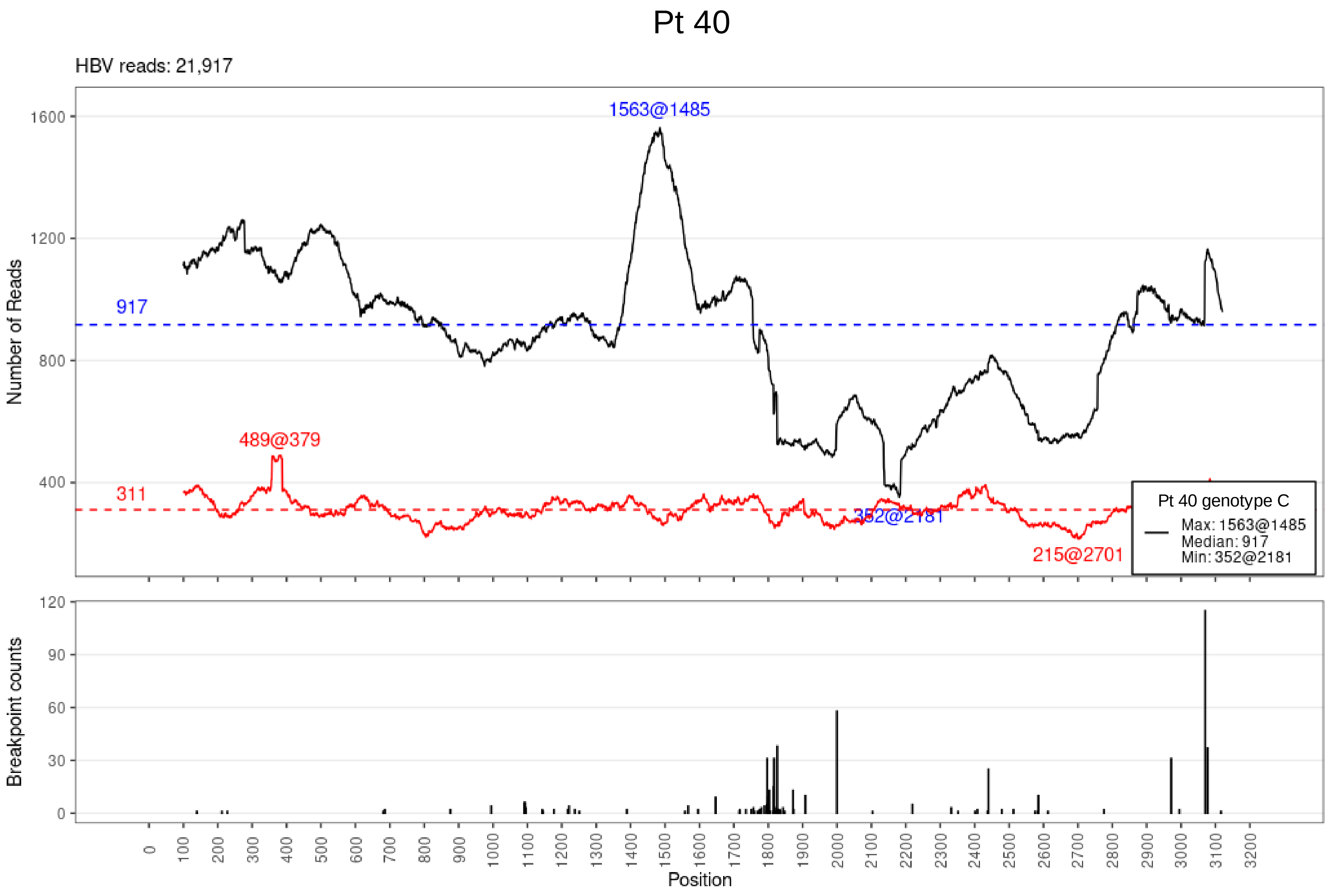

Pt 40
HBV reads: 21,917
1563@1485
917
311
489@379
352@2181
215@2701
Pt 40 genotype C
Max: 1563@1485
Median: 917
Min: 352@2181
Number of Reads
Breakpoint counts
Position

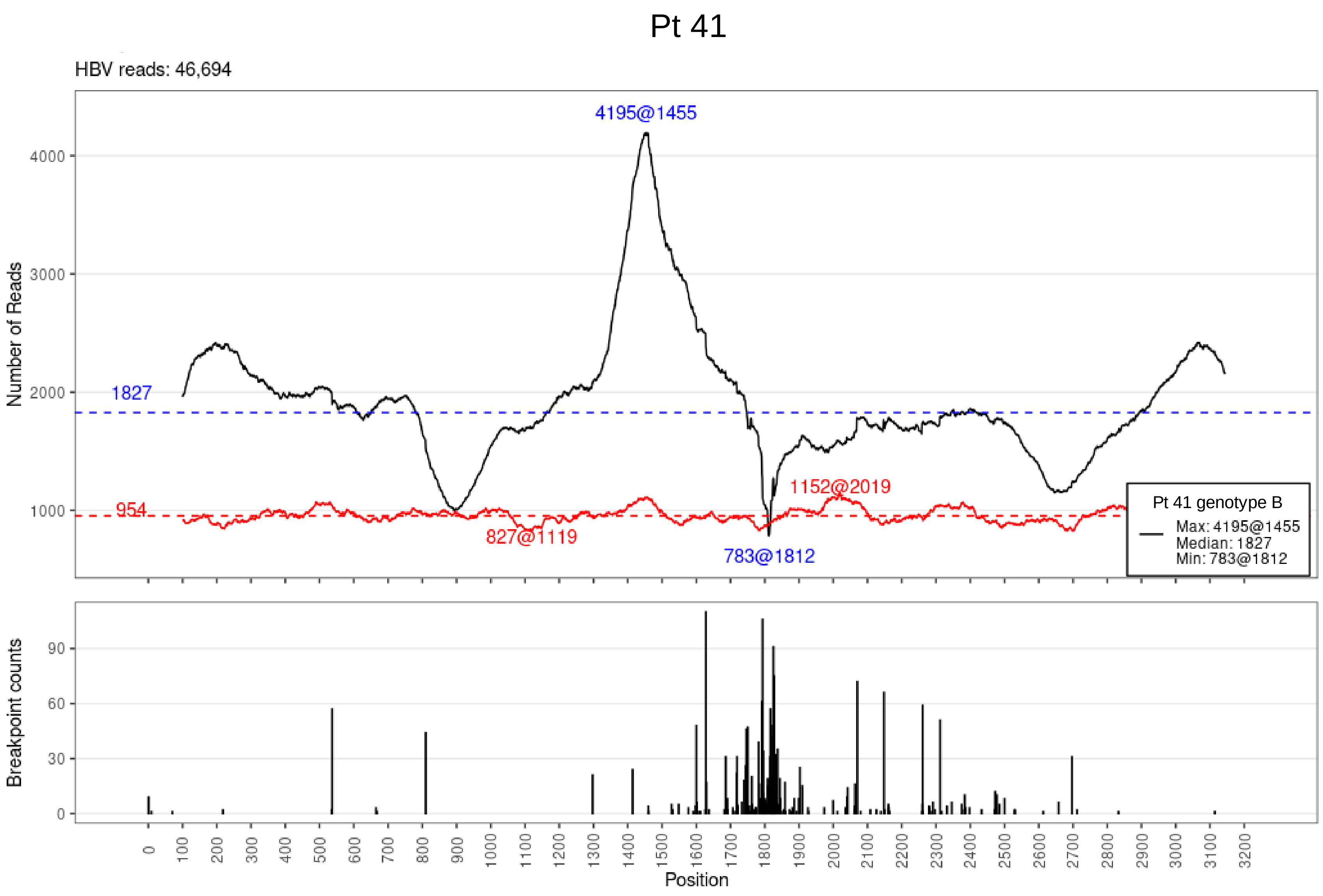

Pt 41
HBV reads: 46,694
4195@1455
1827
954
827@1119
783@1812
1152@2019
Pt 41 genotype B
Max: 4195@1455
Median: 1827
Min: 783@1812
Number of Reads
Breakpoint counts
Position

# Pt 42

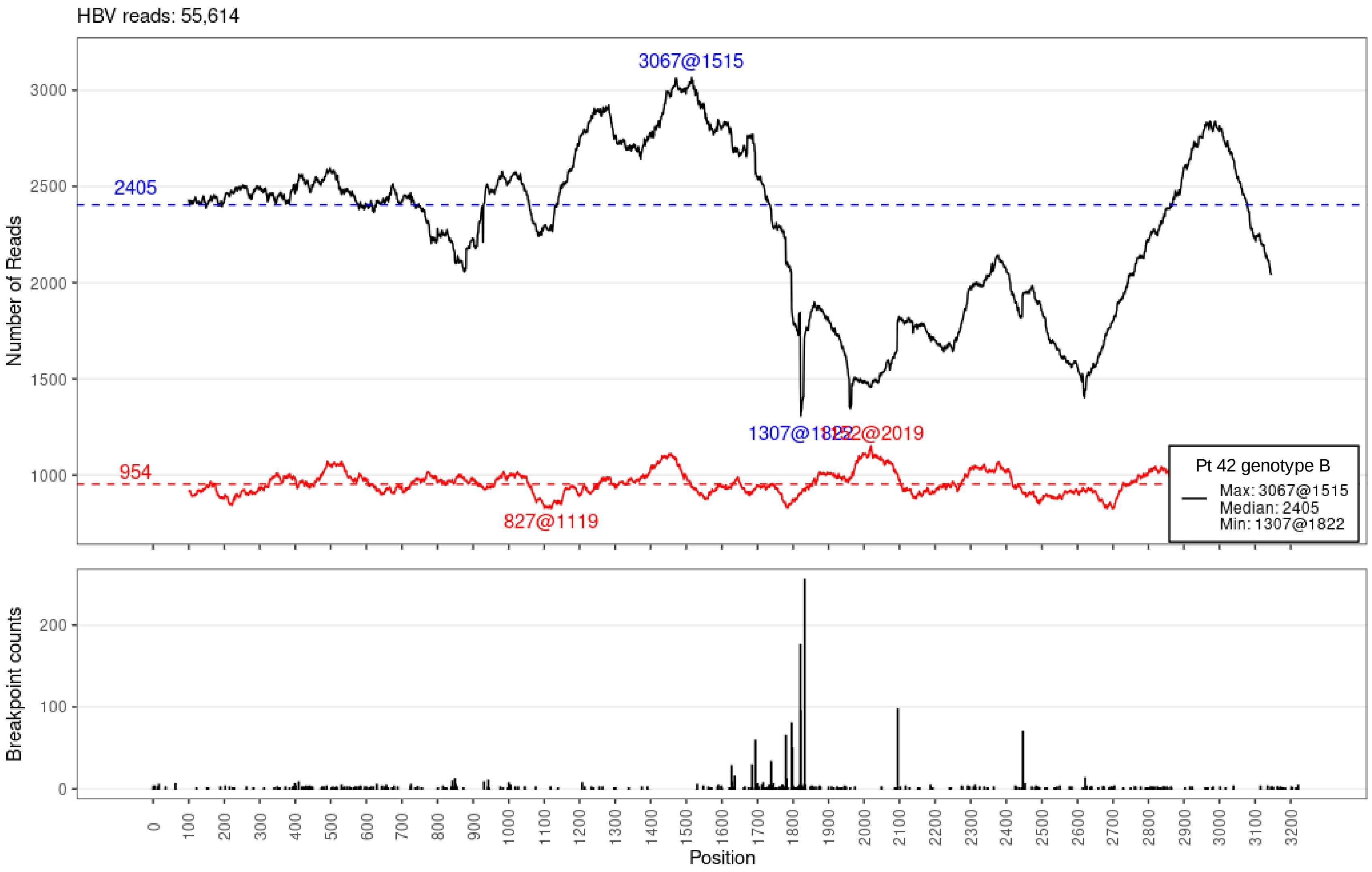

HBV reads: 55,614
Number of Reads
3067@1515
2405
954
827@1119
1307@1822
1152@2019
Pt 42 genotype B
Max: 3067@1515
Median: 2405
Min: 1307@1822
Breakpoint counts
Position

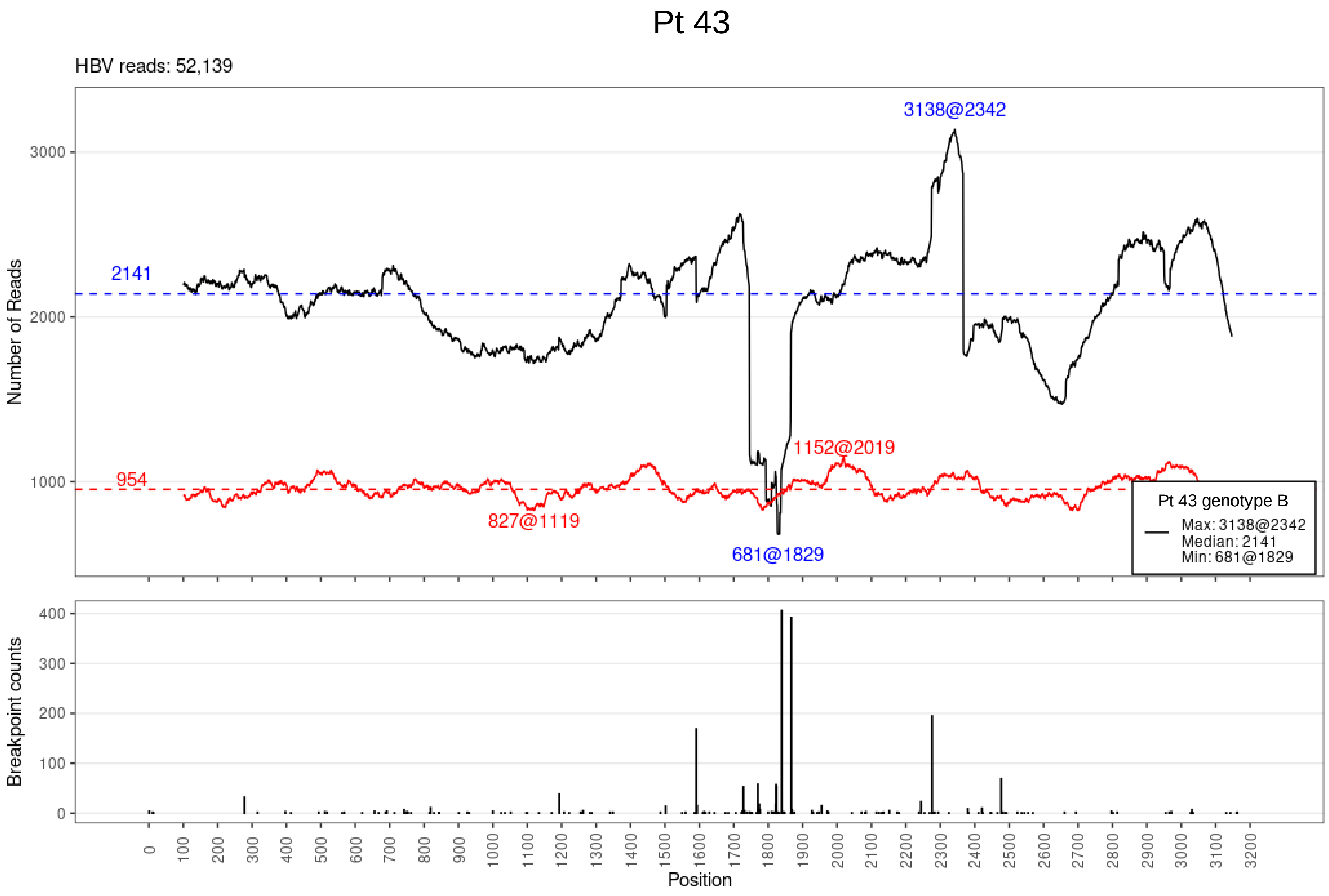
Pt 43
HBV reads: 52,139
3138@2342
2141
1152@2019
954
827@1119
681@1829
Pt 43 genotype B
Max: 3138@2342
Median: 2141
Min: 681@1829
Number of Reads
Breakpoint counts
Position

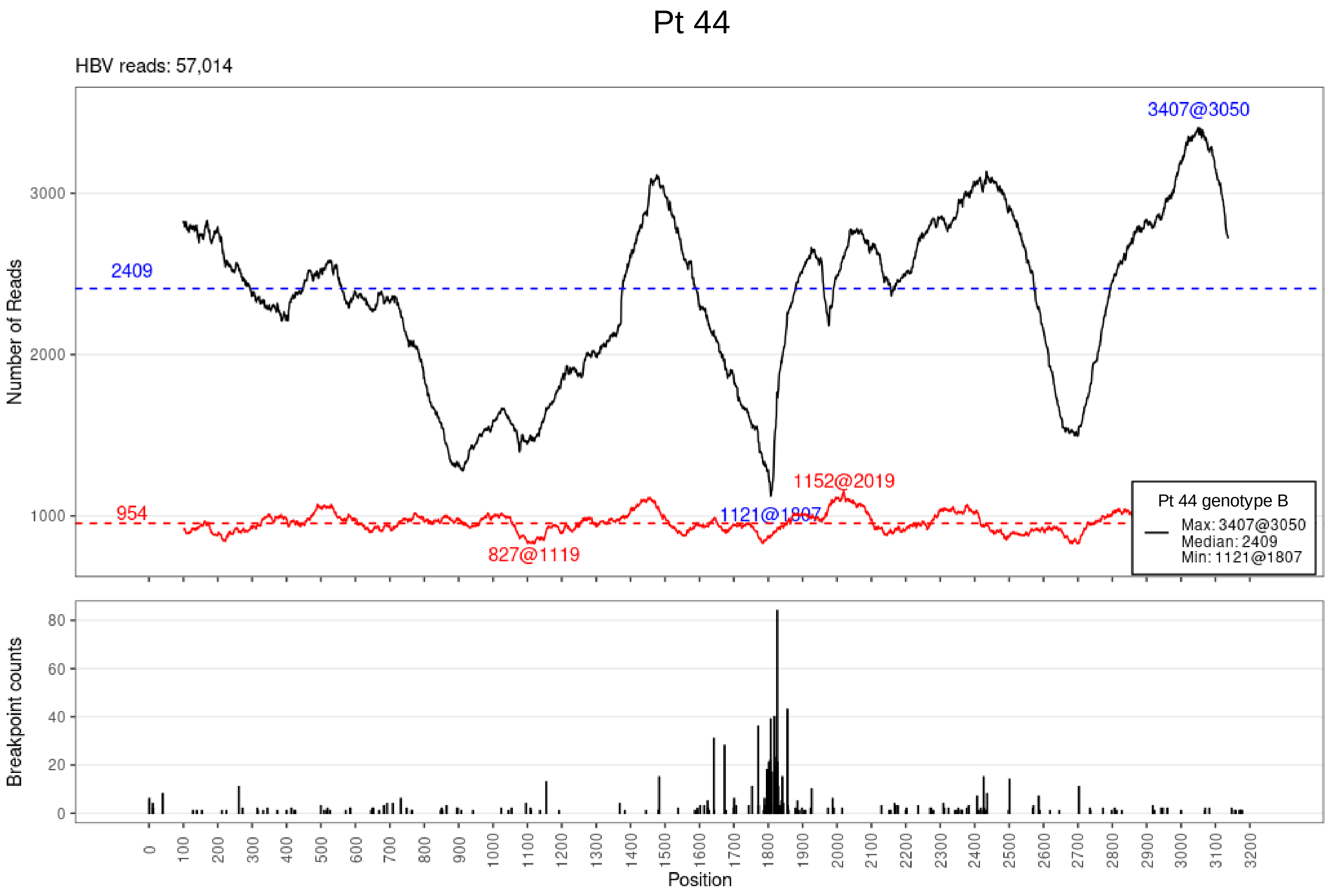

Pt 44
HBV reads: 57,014
3407@3050
2409
954
1152@2019
1121@1807
827@1119
Pt 44 genotype B
Max: 3407@3050
Median: 2409
Min: 1121@1807
Number of Reads
Breakpoint counts
Position

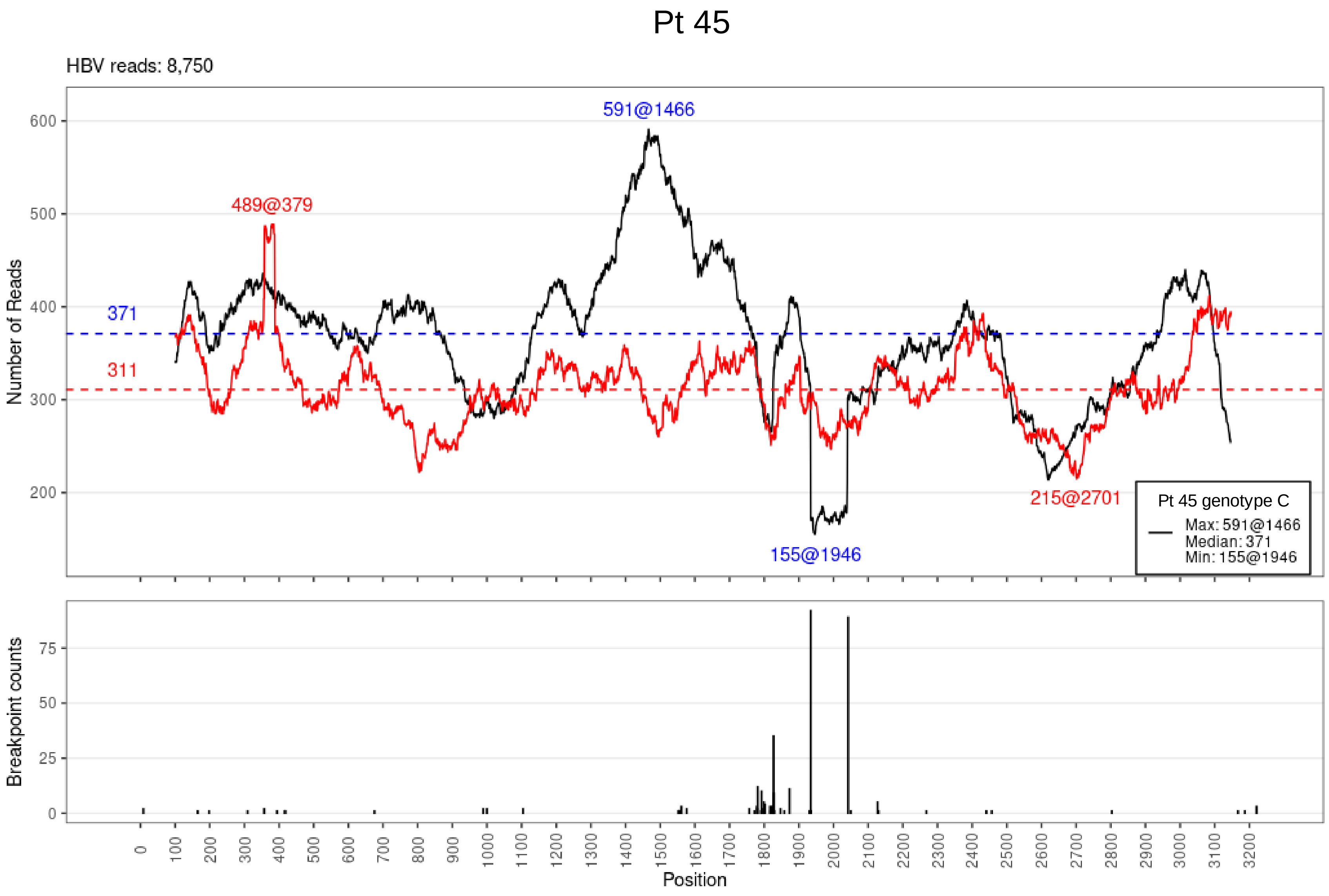

Pt 45
HBV reads: 8,750
591@1466
489@379
371
311
155@1946
215@2701
Pt 45 genotype C
Max: 591@1466
Median: 371
Min: 155@1946
Number of Reads
Breakpoint counts
Position

Pt 46

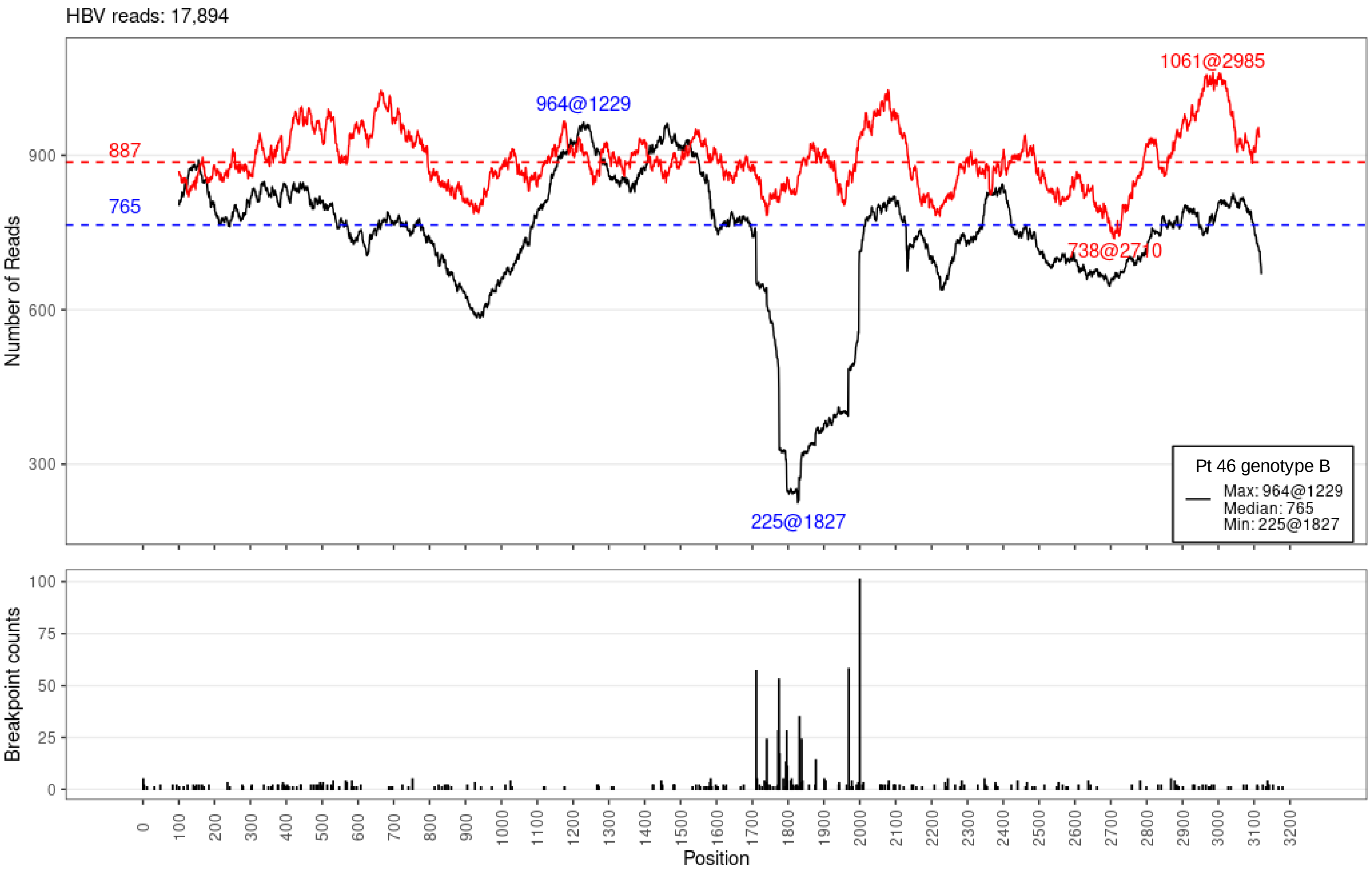

HBV reads: 17,894
Number of Reads
900
600
300
887
765
964@1229
1061@2985
738@2710
225@1827
Pt 46 genotype B
Max: 964@1229
Median: 765
Min: 225@1827
Breakpoint counts
100
75
50
25
0
0
100
200
300
400
500
600
700
800
900
1000
1100
1200
1300
1400
1500
1600
1700
1800
1900
2000
2100
2200
2300
2400
2500
2600
2700
2800
2900
3000
3100
3200
Position

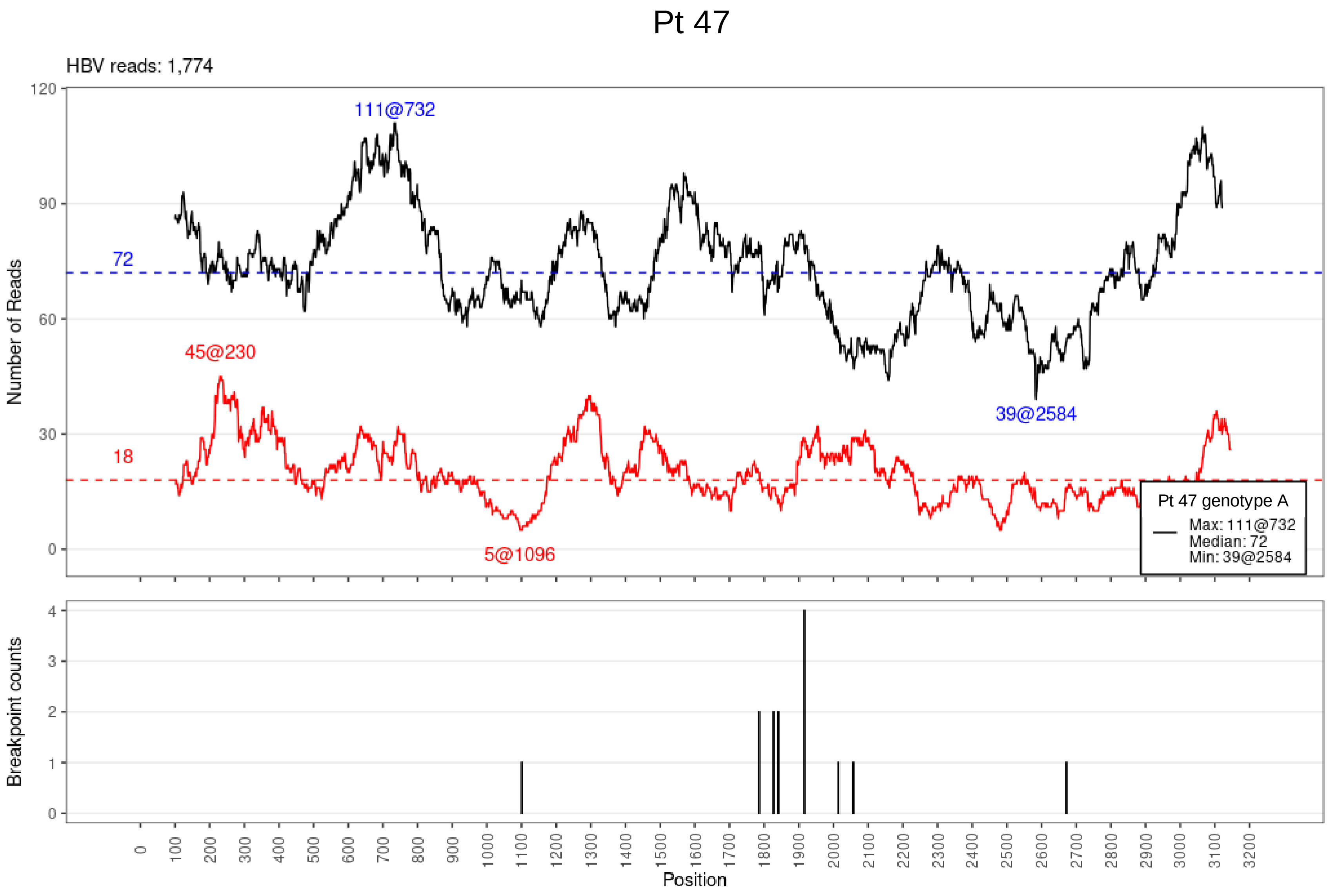

Pt 47
HBV reads: 1,774
111@732
72
45@230
18
5@1096
39@2584
Pt 47 genotype A
Max: 111@732
Median: 72
Min: 39@2584
Number of Reads
Breakpoint counts
Position

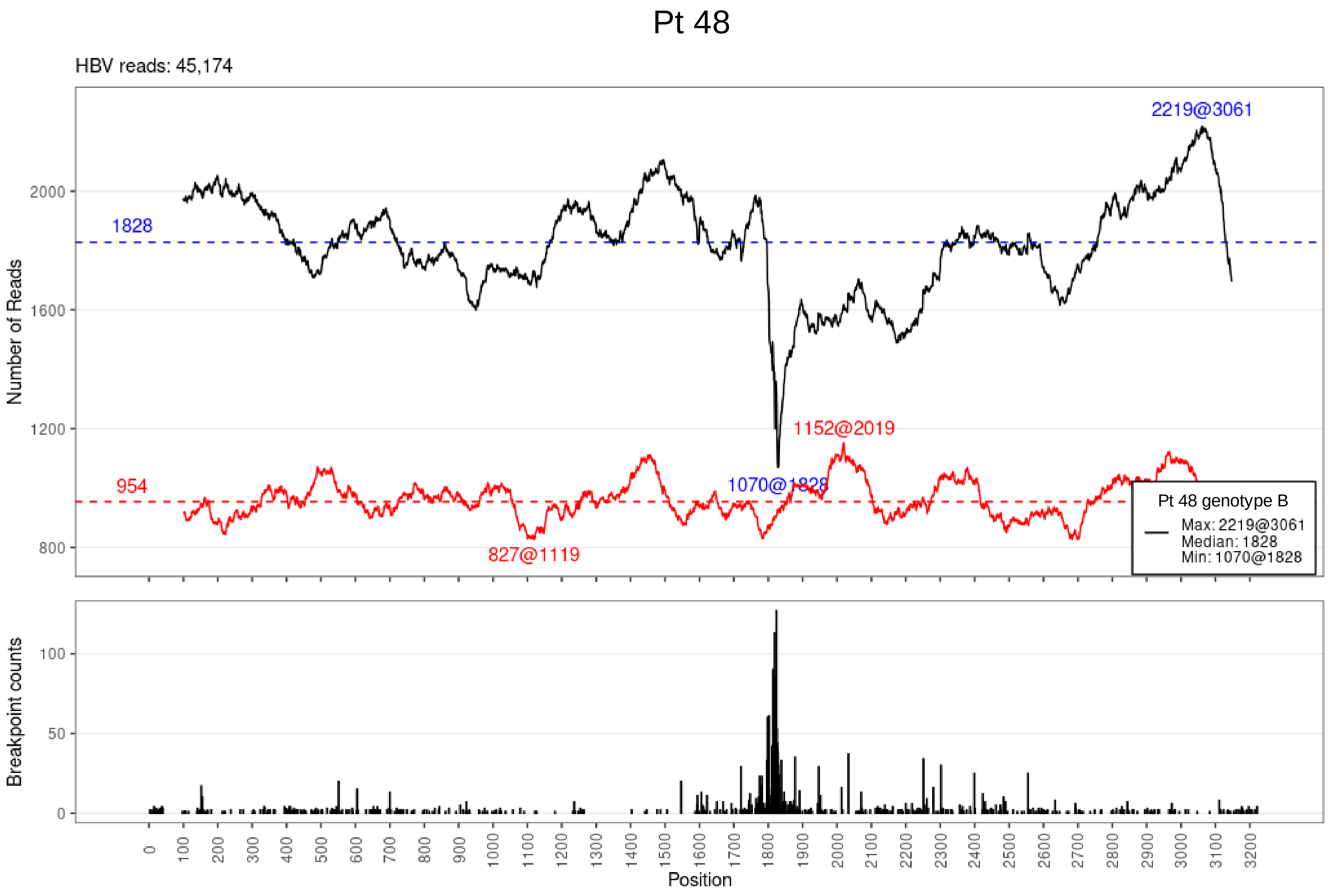

Pt 48
HBV reads: 45,174
Number of Reads
2000
1600
1200
800
1828
954
2219@3061
1152@2019
1070@1828
827@1119
Pt 48 genotype B
Max: 2219@3061
Median: 1828
Min: 1070@1828
Breakpoint counts
100
50
0
0 100 200 300 400 500 600 700 800 900 1000 1100 1200 1300 1400 1500 1600 1700 1800 1900 2000 2100 2200 2300 2400 2500 2600 2700 2800 2900 3000 3100 3200
Position

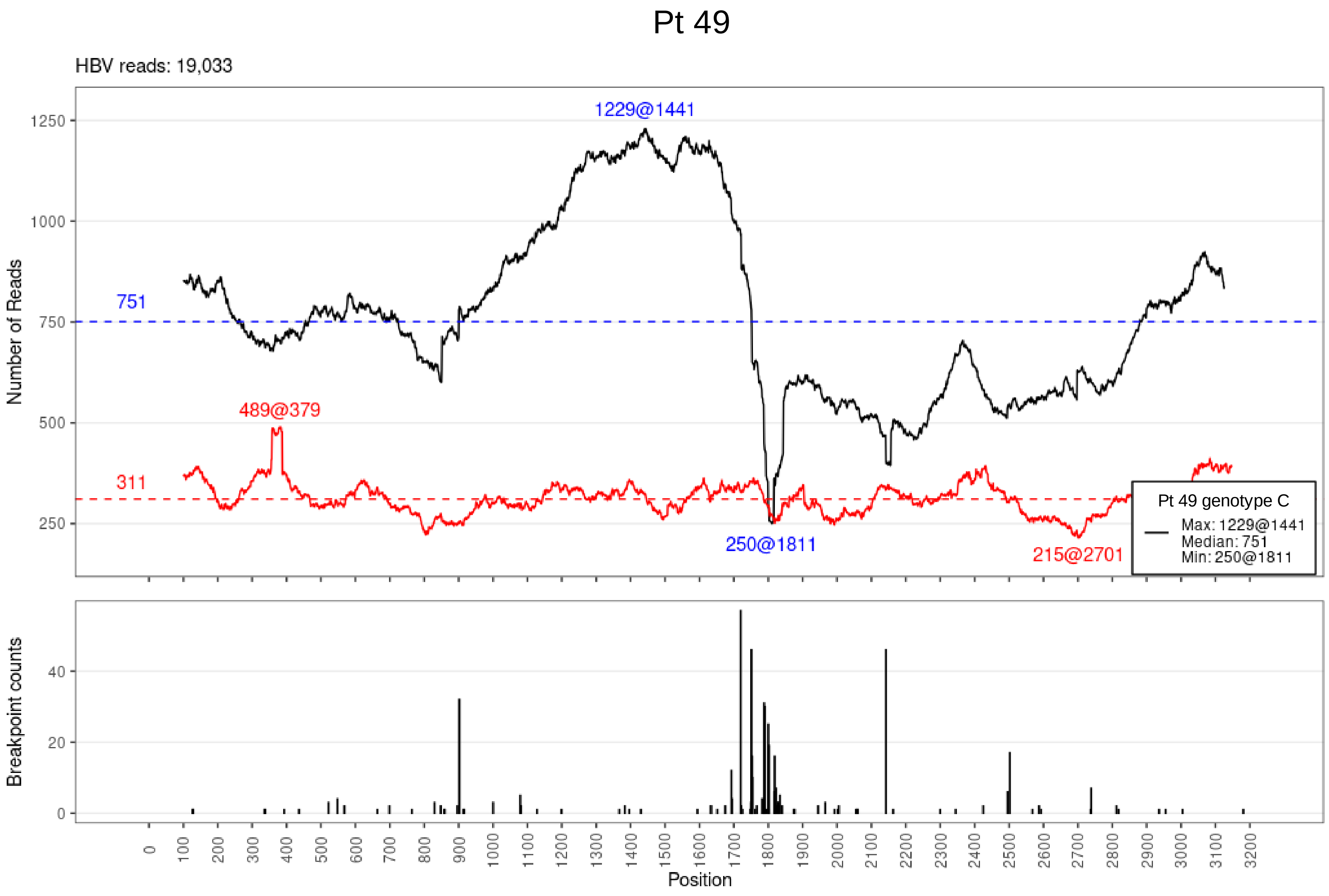
Pt 49
HBV reads: 19,033
Number of Reads
1229@1441
751
311
489@379
250@1811
215@2701
Pt 49 genotype C
Max: 1229@1441
Median: 751
Min: 250@1811
Breakpoint counts
Position

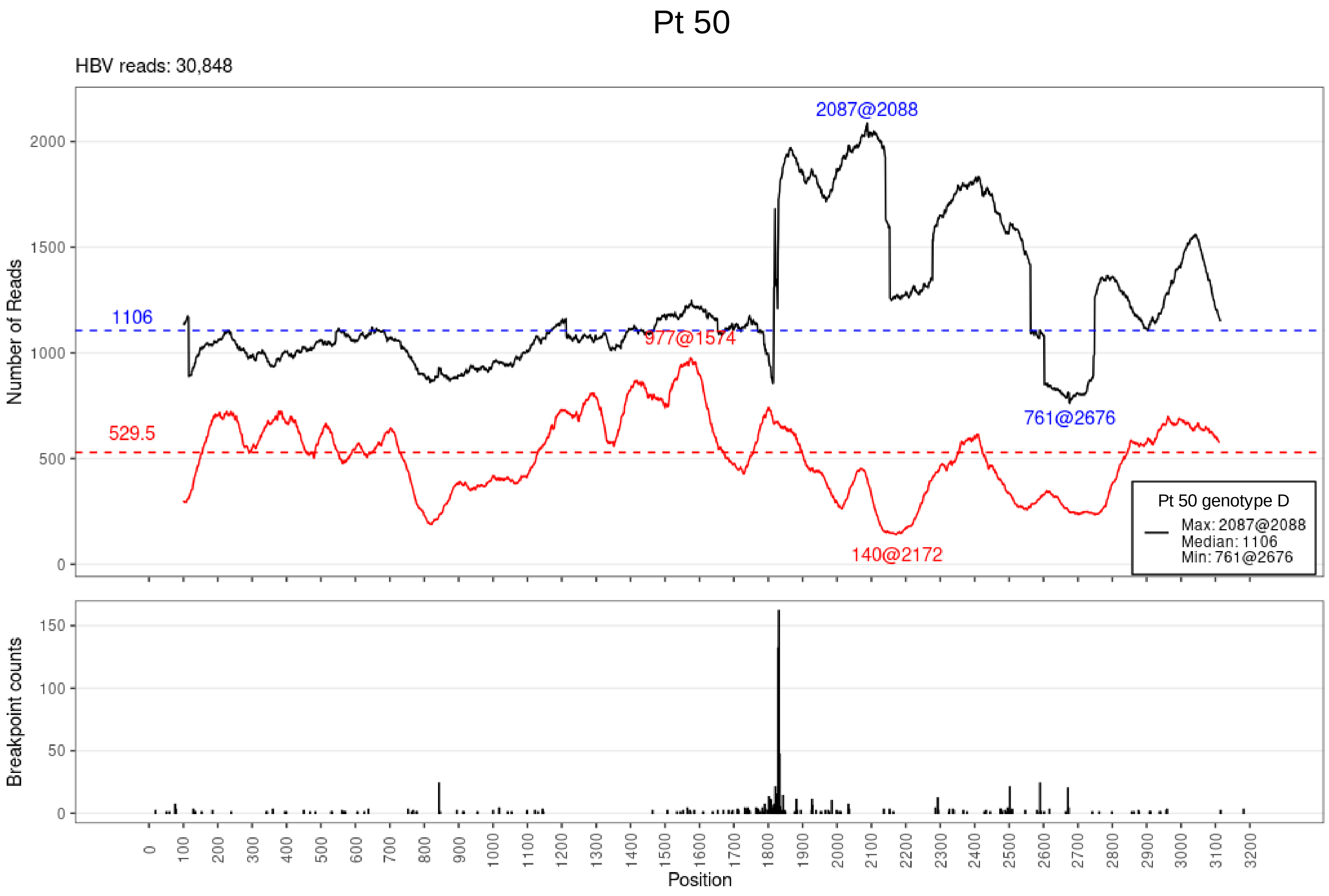

Pt 50
HBV reads: 30,848
2087@2088
1106
977@1574
529.5
761@2676
140@2172
Pt 50 genotype D
Max: 2087@2088
Median: 1106
Min: 761@2676
Number of Reads
Breakpoint counts
Position

# Pt 51

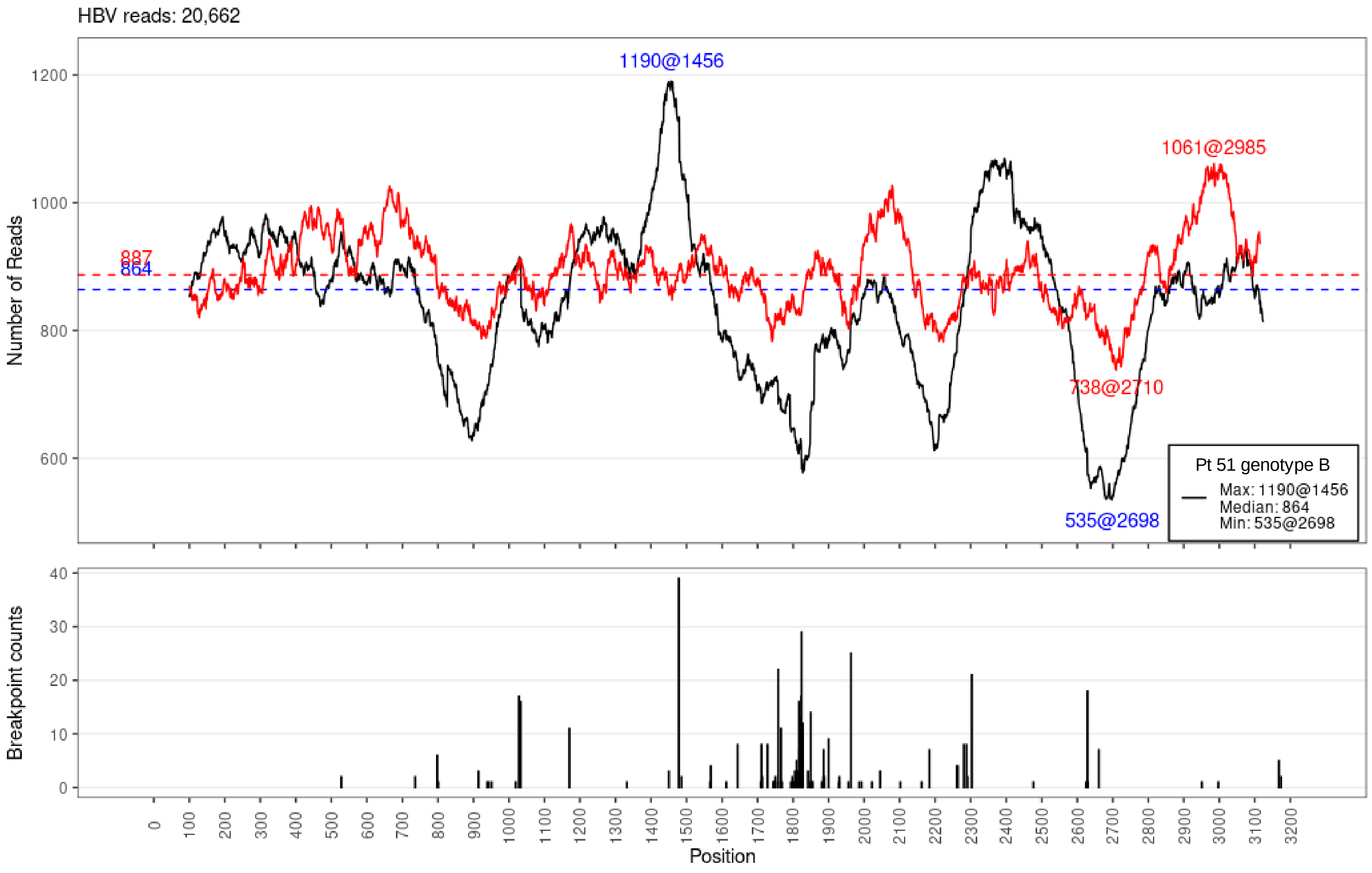

HBV reads: 20,662
1190@1456
1061@2985
887
864
738@2710
535@2698
Number of Reads
1200
1000
800
600
Pt 51 genotype B
Max: 1190@1456
Median: 864
Min: 535@2698
Breakpoint counts
40
30
20
10
0
0
100
200
300
400
500
600
700
800
900
1000
1100
1200
1300
1400
1500
1600
1700
1800
1900
2000
2100
2200
2300
2400
2500
2600
2700
2800
2900
3000
3100
3200
Position

# Pt 52

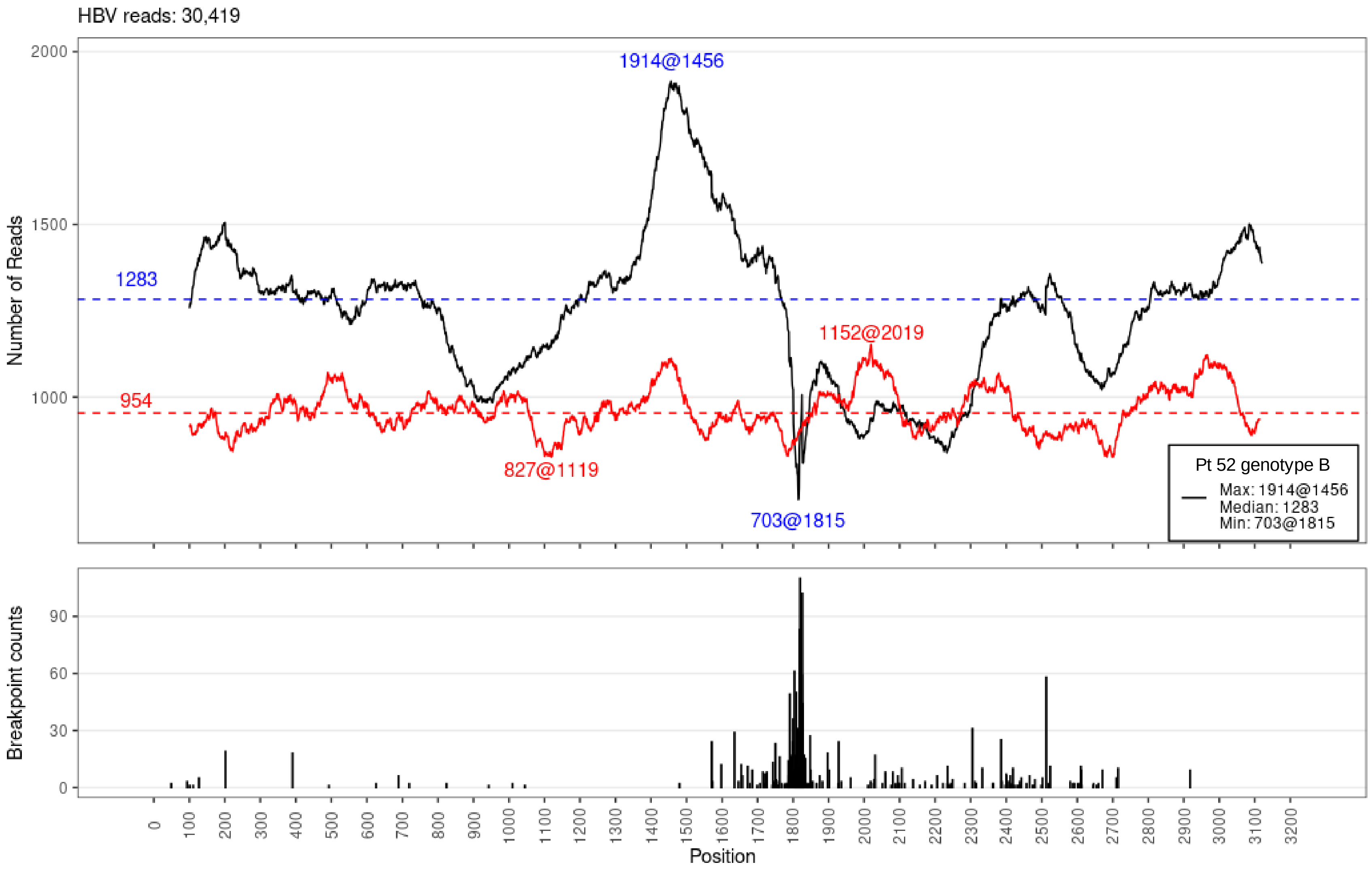

HBV reads: 30,419
1914@1456
1283
954
1152@2019
827@1119
703@1815
Pt 52 genotype B
Max: 1914@1456
Median: 1283
Min: 703@1815
Number of Reads
2000
1500
1000
Breakpoint counts
90
60
30
0
0
100
200
300
400
500
600
700
800
900
1000
1100
1200
1300
1400
1500
1600
1700
1800
1900
2000
2100
2200
2300
2400
2500
2600
2700
2800
2900
3000
3100
3200
Position

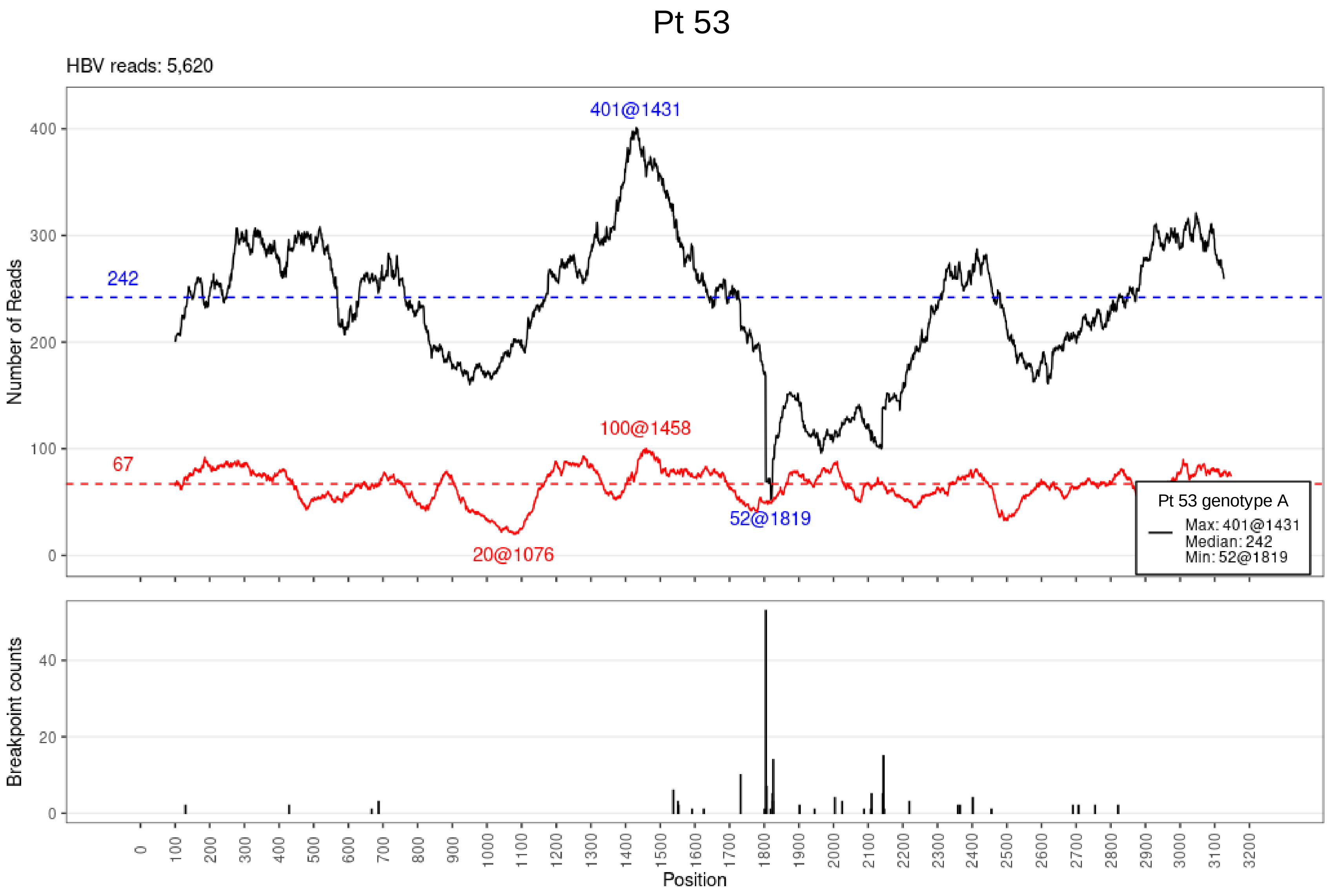

Pt 53
HBV reads: 5,620
Number of Reads
401@1431
242
100@1458
67
52@1819
20@1076
Pt 53 genotype A
Max: 401@1431
Median: 242
Min: 52@1819
Breakpoint counts
Position

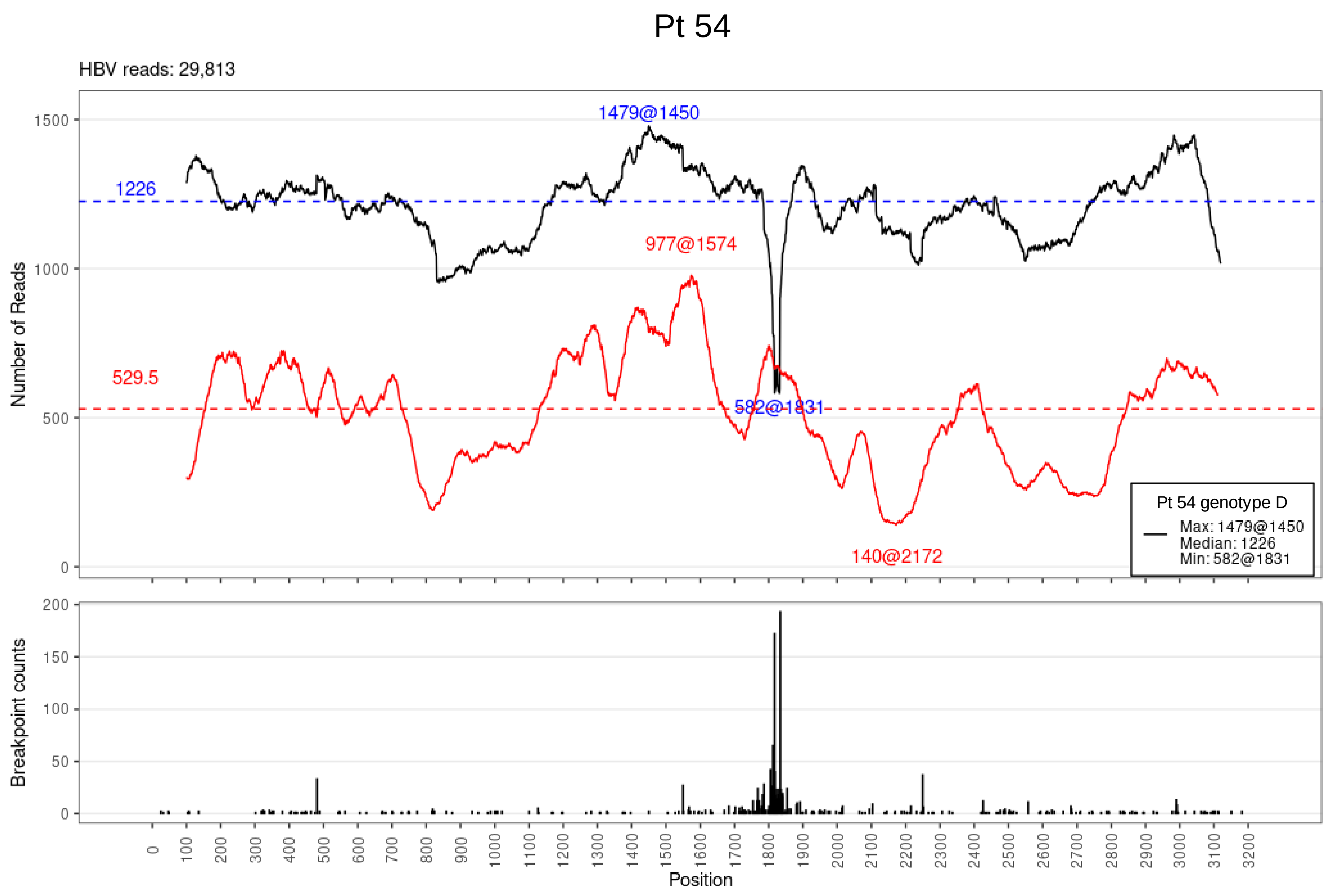

Pt 54
HBV reads: 29,813
1479@1450
1226
977@1574
529.5
582@1831
140@2172
Pt 54 genotype D
Max: 1479@1450
Median: 1226
Min: 582@1831
Number of Reads
Breakpoint counts
Position

# Pt 55

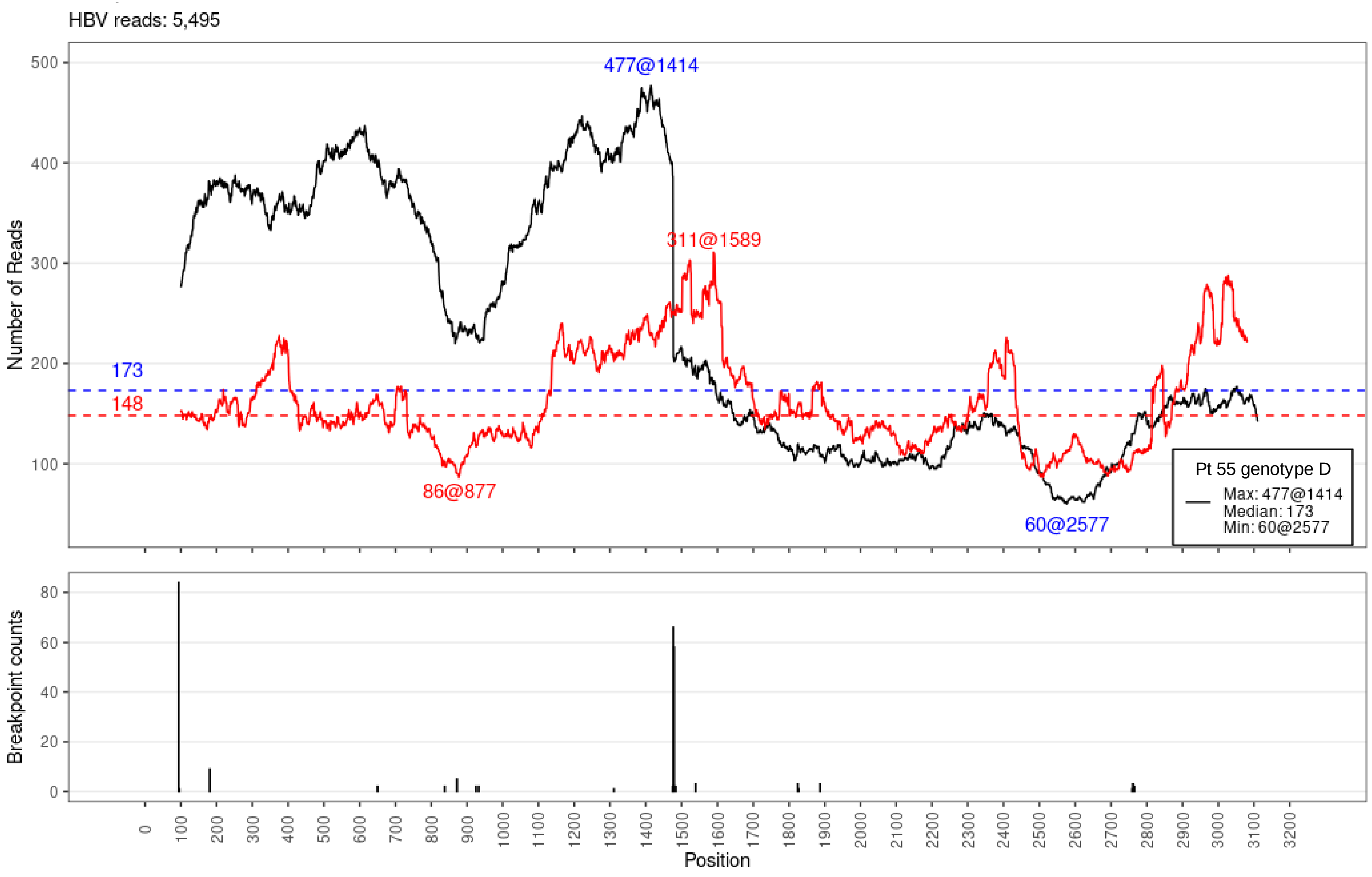

HBV reads: 5,495
477@1414
311@1589
173
148
86@877
60@2577
Pt 55 genotype D
Max: 477@1414
Median: 173
Min: 60@2577
Number of Reads
Breakpoint counts
Position

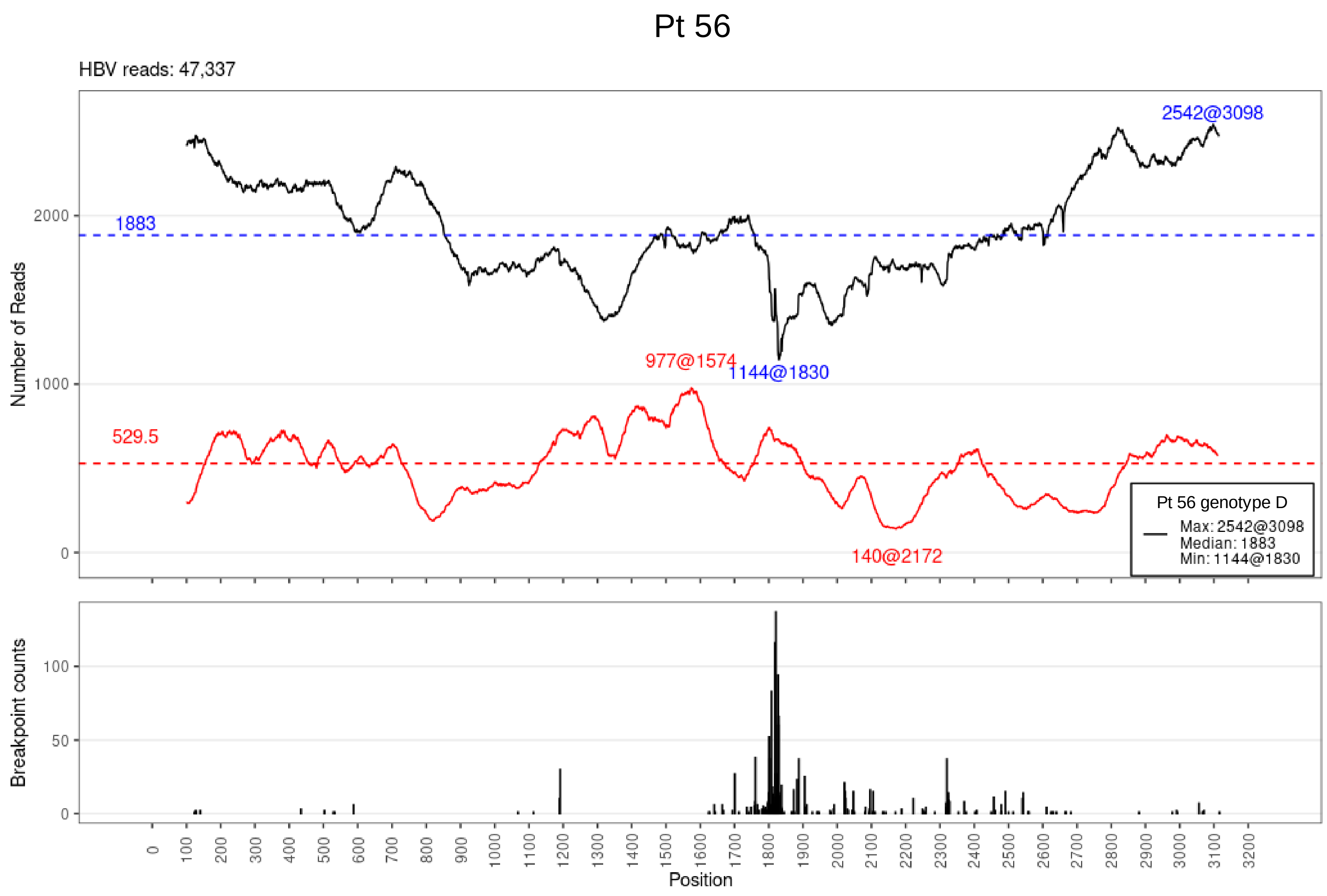

Pt 56
HBV reads: 47,337
Number of Reads
2542@3098
1883
977@1574
1144@1830
529.5
140@2172
Pt 56 genotype D
Max: 2542@3098
Median: 1883
Min: 1144@1830
Breakpoint counts
Position
